# A systematic review to critically appraise methodological rigour in research on ultra-processed food and cardiovascular disease and hypertension

**DOI:** 10.64898/2026.07.09.26357611

**Authors:** Tefera Chane Mekonnen, Zegeye Abebe, Anteneh Mengist Dessie, Teketo Tegegne, Tolassa Ushula, Kacie Dickinson, Catherine Brady, Zumin Shi, Robert Adams, Jason HY Wu, Mario Siervo, Yohannes Adama Melaku

## Abstract

Despite growing research linking ultra-processed food (UPF) consumption to risk of cardiovascular disease (CVD) and hypertension, no study has systematically evaluated the methodological rigor underlying these associations. We systematically searched major databases to identify eligible studies. Data were extracted for dietary assessment methods, UPF classification, covariate selection, confounding control, statistical modelling and effect estimates. Random-effects meta-analysis was conducted to pool effect estimates. Meta-regression and sensitivity analyses were performed to explore sources of heterogeneity. Substantial heterogeneity was observed in the application of the NOVA classification for categorising UPFs across the 46 eligible studies. Only two studies employed a directed acyclic graph to inform confounder selection; 43 used models with suboptimal adjustment, and 42 were overfitted due to adjustment for potential mediators. Pooled analyses indicated that higher consumption of UPFs was associated with a 9% higher risk of CVD and a 16% higher risk of hypertension, with stronger associations observed for coronary heart and cerebrovascular diseases. While higher UPF intake is consistently associated with increased risks of CVD and hypertension, methodological limitations may attenuate the observed associations. Strengthening methodological rigour through harmonized UPF classification and causal frameworks is essential to better elucidate the effect of UPF consumption on cardiometabolic health.

## Introduction

Cardiovascular diseases (CVDs) remain the leading cause of mortality worldwide, accounting for an estimated 17.9 million deaths annually.^1^ Sub-optimal diet is among the leading causes of the observed CVD burden. In recent years, growing attention has been directed toward exploring how overall dietary patterns and food processing levels, particularly the effects of diets high in ultra-processed foods (UPFs), rather than individual nutrients/foods alone, influence health.^2–4^ UPFs, as defined by the NOVA classification system,^5^ are industrial formulations composed largely of food-derived substances and cosmetic additives, with less or no intact whole-food components.^5^ UPFs are generally high in added chemicals, sugars, sodium, and unhealthy fats, and low in dietary fibre, micronutrients, and phytochemicals, a profile that suggest their possible contributions to adverse metabolic and cardiovascular risk.^6^ An accumulating body of evidence from observational studies suggests that higher UPF consumption has consistently been associated with an increased risk of CVD, including hypertension, coronary heart disease, and stroke.^2,7–9^ However, the methodological diversity across studies limit the ability to draw robust conclusions. These include variations in dietary assessment, differences in classification and quantification of UPFs, CVD outcome ascertainment, as well as key confounders considered in the association of UPF and CVD.^10,11^ Most existing studies assess dietary intake through self-administered tools, such as 24-hour recalls or food frequency questionnaires (FFQs), which authors classify these foods into four groups: (1) unprocessed or minimally processed foods □ NOVA group 1; (2) processed culinary ingredients □ Nova group 2, processed foods □ NOVA group 3, and UPFs □ NOVA group 4) based on the extent and purpose of processing using the NOVA food classification system. While pragmatic, this approach is prone to misclassification of foods, particularly when food composition databases are not designed for the purpose and processing-level categorisation.^11–13^ Also, these studies quantify UPF consumption using different metrics, including actual intakes (e.g., grams per day, servings per day, frequency per day) and dietary shares of UPFs, such as their proportion of total daily dietary or energy intake. Similarly, the methods used to ascertain CVD outcomes differ substantially as some rely on self-reported diagnoses, while others use objective data from medical records, hospital registries, or death certificates coded with the international classification of disease (ICD) system.^14–16^ In addition, several studies lack systematic confounders selection and fail to consider mediating pathways, with only a few applying advanced methods, such as causal inference frameworks, to draw robust conclusion on UPF-CVD associations.^17–19^

These challenges in UPF research underscore the need for a rigorous and comprehensive evaluation of the methodological foundations underpinning the evidence base. The current review therefore examines four domains central to the interpretation of UPF–CVD associations: (1) data collection on the assessment of dietary intake, CVD and hypertension assessment; (2) UPF classification and quantification; (3) covariate selection and statistical analysis; and (4) effect estimates. By appraising methodological strengths and limitations, this work aims to inform more rigorous study designs, enable greater comparability across studies, and support the development of standardized approaches for evidence synthesis. Generating accurate and reliable global estimates of the health impacts of UPF, including specific UPF subtypes, is essential to inform future guideline development and evidence-based public policies targeting UPF consumption.

## Materials and methods

### Protocol registration

This review was conducted in accordance with the Preferred Reporting Items for Systematic Reviews and Meta-Analyses (PRISMA) guidelines, which informed the development of the protocol, search strategy, study selection, data extraction, and reporting of findings (**Supplementary Figure 1**).^20^ The protocol was prospectively registered with the International Prospective Register of Systematic Reviews (PROSPERO) (registration ID: CRD42025614620).

### Eligibility criteria

A framework of inclusion and exclusion criteria was developed (**Supplementary Table 1**), drawing on empirical evidence and prior subject-matter knowledge, to evaluate methods of dietary assessment, UPF classification and quantification, and analytical approaches in studies examining the association between UPF intake and CVD outcomes.^10,11,21,22^.

Studies were eligible for inclusion if they met the following criteria:

1. **Study design**: original research articles employing randomized controlled trials, controlled before-and-after interventions, quasi-experimental studies, or prospective cohort studies.
2. **Exposure assessment**: UPF consumption assessed using the NOVA classification system, reported in grams, proportion of weight or energy intake, servings, or frequency per day.
3. **Outcomes**: at least one cardiovascular disease outcome, defined as coronary artery/heart disease, cerebrovascular disease, peripheral vascular disease, or deep vein thrombosis/pulmonary embolism, or hypertension.
4. **Population**: studies that included adults aged ≥18 years.
5. **Publication type**: full-length articles published in peer-reviewed journals.

### Search strategy

We conducted a comprehensive search across multiple electronic databases, including Medline/PubMed, Embase, Web of Science, Cochrane Library, ProQuest Central, CINAHL, Ovid Emcare, PsycINFO, Scopus, and Google Scholar. To ensure completeness, we also hand-searched the reference lists of included studies and relevant reviews and screened grey literature via ProQuest Dissertations and Theses Global. In addition, AI-powered citation mapping tools (e.g., Research Rabbit and Consensus) were used to identify emerging or potentially overlooked articles not captured by conventional databases.

The search strategy was developed by one reviewer (T.C.M.) in consultation with an information specialist (C.B.). A combination of Medical Subject Headings (MeSH) and free-text keywords was applied, including terms such as: “fast food” OR “food, processed” OR “ultra-processed foods” OR “ultraprocessed foods” OR “ultra processed foods” OR “processed meat” AND (“cardiovascular disease” OR hypertension OR “peripheral vascular disease” OR angina OR “cerebrovascular disease” OR “coronary disease” OR “myocardial infarction”). Full search strings for each database are provided in **Supplementary Table 2**. The search was conducted until March 30, 2025, with no restriction on publication date.

### Study selection and data extraction

All retrieved records were imported into the Covidence platform for removing duplication, title and abstract screening, and full-text review. Two reviewers (T.C.M. and Z.A.A.) independently screened titles and abstracts and then assessed full texts against the eligibility criteria outlined in **Supplementary Table 1**. Any discrepancies were resolved through discussion; when necessary, a third reviewer (Y.A.M.) arbitrated. For the purposes of meta-analysis, when more than one publication analysed the same dataset to investigate the association between UPF intake and CVD, we included the study with the largest sample size, greatest number of events, and/or longer follow-up duration. Studies that were purely descriptive, reported only crude effect estimates, or presented non-parametric associations were excluded from quantitative synthesis.

Data extraction was carried out independently in duplicate by two reviewers (T.C.M. and Z.A.A.) using a standardized Excel template. For each study, we collected information on general characteristics, such as the first author, year of publication, country, study design, data source, sample size (both total and analytical), follow-up duration, and health status of participants at baseline. Details of dietary assessment were extracted, including the type of dietary assessment tool employed, whether it had been validated, self-reported or not, the number and timing of dietary data collections (baseline, endpoint, or repeated), the authors involved in applying the NOVA classification, and the unit of UPF measurement (grams, proportion of weight or energy intake, servings, or frequency per day). We also recorded whether implausible dietary energy intakes had been addressed, the list and subgrouping of UPF items, mean UPF consumption, and whether exposure was modelled as a categorical or continuous variable.

Information on cardiovascular outcomes and statistical approaches was also extracted. This included the definition and method of outcome ascertainment, the statistical model applied (conventional regression versus causal inference), the covariates adjusted for, and the strategies used to manage confounding. We collated whether sensitivity analyses were performed, including tests for reverse causation, collider bias, or competing risks. From the results sections of the studies, we extracted the overall mean and variability (SD or SE) of UPF intake, mean consumption across categories, the doses used for dose–response analysis, the number of events per outcome and per UPF category, and the reported effect estimates, such as β-coefficients, odds ratios (ORs), hazard ratios (HRs), risk ratios (RRs), or incidence rate ratios (IRRs), along with their 95% confidence intervals. Finally, information on study funding sources was also recorded. Discrepancies in data extraction were resolved through discussion with the third reviewer (Y.A.M.).

### Assessment of methodological heterogeneity

We evaluated study methods across four domains: (i) data collection; (ii) UPF classification and quantification; (iii) covariate selection and statistical analysis; and (iv) effect estimation. We extracted information on the validity and reproducibility of dietary assessment tools (whether the validation designed to capture the overall dietary intakes or the processing levels of foods), and whether intake was measured once or repeatedly to reflect habitual consumption. Given that NOVA classification involves subjective judgement and risk of misclassification^11^, we also assessed the number and professional backgrounds of coders (nutrition/dietetics-trained vs. non-nutrition), procedures for resolving disagreements, and reporting of inter-rater reliability.

The accuracy of exposure estimates may be affected by measurement error, leading to differential or non-differential bias. We assessed whether studies accounted for total water intake when calculating the relative contribution of UPF (e.g., proportion of weight or servings/day). As water is not associated with adverse cardiovascular outcomes, its inclusion in the denominator can bias associations towards the null and underestimate true UPF–CVD relationships, introducing residual confounding.^23^ We also examined how studies classified alcoholic beverages, given methodological inconsistencies in prospective cohorts.^3,24^ Misclassification of alcohol as UPF may increase heterogeneity in pooled analyses. As alcohol is a well-established CVD risk factor,^25^ we evaluated whether studies excluded it from UPF estimates or applied statistical adjustment, stratification, or sensitivity analyses to distinguish its effects from those of UPF.

In nutritional epidemiology, four models are commonly considered to adjust total energy intake when estimating the causal effect of certain foods-health outcomes (the standard model-average relative causal effect, the energy partition model-total causal effect, nutrient density model-average relative causal effect, and the residual method-average relative causal effect),^26^ even though contrasting thoughts existed.^27,28^ However, the apparent heterogeneity among existing nutritional studies lies on lack of awareness of the estimand differences and accuracy of the above modelling approaches, which raises serious questions regarding the validity of meta-analyses where different estimands have been inappropriately pooled. Several sources of bias can be introduced when interpretating energy-adjusted estimates. First, because total energy intake lies on the causal pathway between food consumption and health outcomes, adjustment may introduce overadjustment bias by removing parts of the effect mediated through excess energy intake. Second, conditioning on total energy intake may induce collider bias, as it is influenced by both the exposure (UPF) and unmeasured determinants such as behavioral clustering (appetite, physical activity, and body size) and metabolic factors. Third, dietary data are subjected to substantial measurement error, and errors in total energy intake are often correlated with errors in specific food intakes. Under such conditions, energy adjustment can amplify or attenuate true associations, which is more pronounced when all-compartment/ partition model applied,^27^ and particularly when misreporting varies by participant characteristics such as body mass index (BMI) or health status. Finally, in populations with prevalent chronic disease, reduced energy intake may reflect underlying illness, raising the possibility of reverse causation when conditioning on total energy intake. Therefore, we evaluated each included studies on how total energy intake was accounted in the model and their interpretation (substitution vs total effect).

We assessed outcome ascertainment, distinguishing between objective sources (e.g., national death registries, hospital databases, validated ICD-coded records) and self-reports or physician diagnoses. To evaluate control of confounding, we extracted details on modelling approaches, covariates, and model types. A minimally sufficient confounder set for the UPF–outcomes relationship was defined (**Supplementary Figure 2**) using a directed acyclic graph (DAG), and each study was assessed against it to identify potential residual confounding, collider bias, and model overfitting (e.g., adjustment for mediators). The DAG was constructed iteratively by the research team based on a combination of: (1) existing literature on dietary determinants of cardiovascular disease; (2) subject-matter expertise in nutritional epidemiology and causal inference; and (3) previously published DAGs in the field of ultra-processed food research.^3,29,30^ Despite it involves inherent subjectivity and that alternative structures could be reasonably proposed depending on differing assumptions about causal relationships, DAG provides a structured framework to map confounding pathways, detect inappropriate adjustments, and clarify the role of confounders, mediators, or colliders.^30^

### Risk of bias assessment

Risk of bias was assessed at both the study and outcome levels. We used the Risk Of Bias In Non-randomized Studies – of Exposures (ROBINS-E) tool as the primary instrument for risk of bias assessment, given that this review focuses on observational studies of dietary *exposures (UPF)* (**Supplementary Figure 3**). ROBINS-E is specifically designed for this context and evaluates seven domains of bias relevant to exposure studies, with the overall risk judged as low risk of bias, some concerns, high risk of bias, very high risk of bias, or concerns about residual confounding.^31^ We evaluated the certainty of evidence using the Grading of Recommendations, Assessment, Development and Evaluation (GRADE) system,^32,33^ which classifies evidence as high, moderate, low, or very low for each pooled UPF–CVD and -hypertension associations. Evidence was downgraded based on limitations in risk of bias, inconsistency/heterogeneity, indirectness, imprecision, or publication bias, and upgraded where large effects or exposure–response gradients were observed. The GRADE assessment provided a transparent summary of the confidence in pooled effect estimates derived from the meta-analyses (**Supplementary Table 3**).

### Data synthesis and statistical analysis

For all included studies, qualitative summary was undertaken by demographic characteristics, dietary assessment and UPF classification methods, outcome ascertainment, and approaches to confounding mitigation. Quantitative summary was generated using meta-analysis by pooling the most adjusted effect estimates for overall UPF-CVD and -hypertension associations and specific CVD outcomes (coronary heart disease and cerebrovascular disease). We pooled studies that used the lowest UPF consumption category as the reference and compared it with the highest category, ensuring consistent units of exposure (studies with the same units of measurement). Effect estimates were expressed as risk ratios (RRs) with 95% CIs. Adjusted odds ratios (ORs) were converted to RRs using the formula: *RR* = *OR* / [(1 − *P*_0_) + (*P*_0_ × *OR*)], where OR is the odds ratio and *P*_0_ is the incidence of the outcome in the unexposed group.^34^ Adjusted hazard ratios (HRs) and incidence rate ratios (IRRs) were used as reported.^35^ For studies reporting continuous exposure, we converted estimates to a standardized increment where possible. We performed random-effects meta-analyses to synthesise the association between UPF consumption and each outcome. Effect estimates were log-transformed and pooled using the generic inverse-variance method with a random-effects model. Between-study variance was estimated using the Restricted Maximum Likelihood (REML) method.

Heterogeneity was quantified using the *I*^2^ statistic and *T*^2^. We also calculated 95% prediction intervals to provide a range within which the true effect size of a future study is expected to fall. Sources of heterogeneity were explored through pre-specified meta-regression analyses using the following covariates: sex (male/female/mixed), geographic region, dietary assessment tool (24-hour recall and food frequency questionnaire [FFQ]), follow-up duration (short: < 10 years and long: ≥10 years), energy adjustment methods (standard, residual, and nutrient density) and unit of UPF exposure (percentage of energy, grams/day, servings/day, and others).

Sensitivity analyses were conducted using the trim-and-fill method to assess the potential impact of publication bias and the leave-one-out method to evaluate the influence of individual studies on the pooled estimate.

All analyses were conducted using the *meta* and *metafor* packages in R (*version 4.3.2; R Foundation for Statistical Computing, Vienna, Austria*).

## Results

### Study characteristics

After screening of 13,055 articles, 248 full-text studies were assessed in detail for eligibility. Of these, 46 unique studies published between 2017, and January 2025 were included for evidence synthesis. These studies were comprised of a controlled quasi-experimental intervention,^36^ two modelling studies,^37,38^ and 43 prospective studies (**Supplementary Figure 1**).^3,4,24,39–54^ The studies included a total of 4,335,935 adults (aged 18-79 years at baseline). Multiple studies used data from the same data sources but with different methods, unique subsamples or outcome type (**Table 1**): eight from the UK Biobank,^4,43,45,54–58^ four from Moli-Sani,^44,47,49,59^ three from Cohort of Universities of Minas Gerais (CUME),^53,60,61^ three from Nurses’ Health Study (NHS) and the Health Professionals Follow-up Study (HPFS),^29,48,62^ two from Prostate, Lung, Colorectal, and Ovarian (PLCO) Cancer Screening Trial,^3,57^ two from National Health and Nutrition Examination Survey (NHANES),^57,63^ two from the Malmö Diet and Cancer Study (MDC),^64,65^ two from the Tehran Lipid and Glucose Study (TLGS),^66,67^ and two from the Atherosclerosis Risk in Communities (ARIC) Study.^40,68^ Although majority of studies originated from high-income regions, all continents were represented, including North America (USA,^3,29,36,40,48,52,57,62,63,68–72^ Canada,^69,73^ Mexico^39^), South America (Brazil,^37,38,60,61,73,74^ Chile,^73^ Argentina^73^,Colombia^73^), Europe (UK,^4,24,43,45,54–58^ France,^42^ Spain,^24,46,50,53,75^ Italy,^24,44,47,49,59^ Sweden,^24,64,65,73^ Denmark,^24^ Germany,^24^ the Netherlands^24,73^), Australia,^76^ Asia (China,^73,77^ Malaysia,^73^ Philippines,^73^ Korea,^41^ Turkey,^73^ Iran,^73^ Palestine,^73^ Saudi Arabia,^73^ United Arab Emirates,^73^ Bangladesh,^73^ India,^73^ Pakistan^73^) and Africa (South Africa, Tanzania, Zimbabwe).^73^ Two studies exclusively enrolled female participants^39,76^ while the rest included both sexes.

**Table 1.** Summary of characteristics of prospective studies examining the association between ultra-processed food consumption and cardiovascular outcomes (n=46)

| Author year | Country | Cohort Name | Sex | Total Sample (Female) | Population characteristics | Mean Age (SD) | Median follow-up (IQR) |
| --- | --- | --- | --- | --- | --- | --- | --- |
| Rivera 2024 | USA | ARIC (Atherosclerosis Risk in Communities) study | Both | 8923 (4,818) | 45-64 years | 53□years | 13 years |
| Wang 2024 | UK | UK Biobank | Both | 90,631 (50,831) | 40–69 years | NR | 11.6 years |
| Fang 2024 | USA | NHS and HPFS cohorts | Both | 114,064 (74,563) | Women aged 30-55 years and men aged 40-75 years | NR | 34 years |
| Torres-Collado 2024 | Valencia, Spain | VNS | Both | 1,538 (849) | ≥ 20 years | 46.2 years (18) | Range: 18 years of follow-up |
| Kermani-Alghoraishi 2024 | Central part of Iran | ICS | Both | 5432 (2,784) | ≥ 35 years | 50.7 years (11.6) | NR |
| Oladele 2024 | USA | REGARDS | Both | 5957 (3278) | ≥ 45 years | NR | Mean: 9.3 years |
| Pant 2024 | Australia | ALSWH | Female | 10,006 (10,006) | Women aged 50–55 years | 52.5 years (1.5) | NR |
| Hang 2024 | USA | NHS and HPF | Both | 2,498 (1,764) | ≥ 35 years with stage I to III CRC | 68.5 years (9.4) | 11.0 years |
| Golzarand 2024 | Iran | TLGS | Both | 2399 (NR) | ≥ 25 years | 37.6 years | Mean: 9.21 years |
| Rauber 2024 | UK | UK Biobank | Both | 126,842 (67,551) | 40-69 years | 55.9□years (7.8) | 9 years |
| Du 2024 | Sweden | MDCS | Both | 27,670 (16,787) | 45-73 years | 58.11□years (7.6) | 23.3 years |
| Zhao 2024 | USA and UK | PLCO, NHANES and UK Biobank | Both | 108,714 (NR) | 40-60 years | 56.11 years (7.94) | 16.94 years |
|  |  |  |  | 41,070 (NR) | ≥ 20 years | 47.71 years (7.94) | 12.44 years |
|  |  |  |  | 208,051 (NR) | 55-74 years | 62.51□years (5.3) | 9.33 years |
| Jalali 2024 | Iran | TLGS | Both | 2050 (1,107) | ≥ 30 years | 46.28 years (11.34) | 10.6 years |
| Mitra 2024 | USA | NR | Female | 19 (19) | Female University Students | 28□years | 8-weeks of dietary intervention |
| Li 2024 | UK | UK Biobank | Both | 5,405 (2,092) | 37-73 years with type 2 diabetes | 59.4□years (6.9) | NR |
| Mendoza 2024 | USA | NHS; NHSII and HPFS | Both | 206,957 (166,548) | 30-55 years (NHS)<br>40-75 years (HPFS) | 50.8 years (HNS)<br>36.7 years (NHSII)<br>53.4 Years (HPFS) | 31.9 years (NHS)<br>26.0 years (NHSII)<br>29.7 years (HPFS) |
| Cordova 2023 | 7 European countries | EPIC | Both | 266,666 (160,550) | 35 and 74 years | 51.6 years for women and<br>52.3 years for men | 11.2 years (IQR: 9.8–12.7) |
| Kityo 2023 | Korea | KoGES-HEXA cohort | Both | 113,576 (74,729) | ≥ 40 years | 52.0 years (0.1) | 10.6 years (IQR: 9.5–11.9) |
| Li 2023 | UK | UK Biobank | Both | 111,646 (63,777) | 40–69 years | NR | 10.0 years |
| Bonaccio 2023 | Italy | Moli-sani Study | Both | 1,065 (419) | ≥ 35 years old with type 2 diabetes | 65.2 years | 11.6 years |
| Sullivan 2023 | USA | CRIC | Both | 1,868 (NR) | 21-74 years with reduced GFR | NR | 12 years |
| Passinho 2023 | Brazil | CUME | Both | 1,286 (845) | ≥ 18 years | 31.9 years | Range: 4 years of follow-up |
| Shuai Yuan 2023 | UK | UK Biobank | Both | 186,233 (102,199) | 40-69 years | 56.15□years (7.93) | 10.5 years |
| Tu 2023 | UK | UK Biobank | Both | 121,300 (68,413) | 40-69 years | 59.4 years (7.8) | 8.8 years (IQR: 8.7–9.2) |
| Li 2023 | Sweden | MDCS | Both | 26,369 (16,428) | 45-73 years | 58□years (7.62) | 24.6 years |
| Rezende-Alves 2023 | Brazil | CUME | Both | 1,547 (1,094) | University undergraduate and/or graduate young adults | Median: 35 (IQR: 30–43) | NR |
| Dehghan 2023 | North and South America, Europe, Africa, the Middle East, and Asia | PURE | Both | 138,076 (80,775) | 35-70 years | 50.1□years (10) | 10.2 years (IQR: 8.5–12.1) |
| Silva 2023 | Brazil | ELSA-Brazil cohort | Both | 14,747 (8,025) | 35–74 years | 52□years (9.1) | NR |
| Wang 2023 | USA | SCCS | Both | 77,060 (46,175) | 40-79 years | 52.4□years (8.8) | NR |
| González-Palacios 2023 | Spain | PREDIMED-Plus Study | Both | 5,373 (2,591) | 55-75 years old | 65.1 years (4.9) | 6-12 months |
| Chen 2022 | UK | UK Biobank | Both | 60,298 (33,531) | 40 and 69 years | Median: 57 years | 10.9 years |
| Bonaccio 2022 | Italy | ✓ Moli-sani Study | Both | 22,895 (11,973) | ≥35 years | 55.4 years (11.7) | NR |
| Orlich 2022 | North America (US and Canada) | AHS-2 | Both | 77,437 (NR) | ≥ 25 years | NR | Mean: 7.46 years |
| Moreira 2022 | Brazil | Household Budget Survey | Both | 34,003 (NR) | ≥ 25 years | NR | NR |
| Bonaccio 2022 | Italy | Moli-sani Study | Both | 1171 (377) | ≥ 35 years | 67□years (10) | 10.6 years (IQR: 9.2–11.9) |
| Nilson 2022 | Brazil | ✓ Brazilian populations Estimates<br>✓ GBD Study<br>✓ Brazilian Dietary Survey (2017-2018)<br>Pagliai et al. Article | Both | NR | 30 to 69 years | NR | NR |
| Li 2022 | China | CHNS | Both | 15,054 (7,918) | ≥ 20 years | 40.2□years (14.4) | NR |
| Zhong 2021 | USA | PLCO | Both | 91,891 (49,348) | 55 and 74 years | 65.3 years | Mean: 13.5 (3.3) years |
| Monge 2021 | Mexico | MTC | Female | 64,934 (64,934) | Disease-free women aged ≥25 years at baseline | 41.7 years (7.2) | 2.2 years (IQR: 1.8-4.4) |
| Du 2021 | USA | ARIC study | Both | 13,548 (7,568) | 45–65 years | 54 years | 27 years |
| Bonaccio 2021 | Italy | Moli-sani Study |  | 22,475 (NR) | ≥35 years | 55 years (12) | 8.2 years (IQR: 7.3–9.3) |
| Rezende-Alves 2021 | Brazil | CUME | Both | 1,221 (929) | University Graduates | 35.2□years | NR |
| Juul 2021 | USA | Framingham Offspring Study | Both | 3,003 (1656) | middle-aged adults | 53.9 years (9.6) | Mean: 18 years for incidents and 20.2 for mortality |
| Srour 2019 | France | NutriNet-Santé | Both | 105,159 (83,247) | ≥ 18 years | 42.7 years (14.5) | 5.2 years |
| Hyun-JuKim 2019 | USA | NHANES III | Both | 11,898 (6,160) | ≥ 20 years | NR | 19 years |
| Mendonca 2017 | Spain | SUN Project | Both | 14,790 (9,416) | ≥ 18 years | NR | Mean: 9.1 years |

## PART I: Data collection

### Heterogeneity in dietary assessment tools

Dietary intake was mainly assessed using FFQs in 32 studies (69.6%), 24-hour dietary recalls in 13 studies (28.3%), and a combination of both methods in one study. All dietary assessments were self-reported. Although validation of the dietary assessment tools was reported in most included studies, two FFQs^40,71^ and one 24-hour recall ^36^ lacked reported validation. Importantly, none of the validations were specifically designed for UPF intake; instead, they evaluated total energy intake, nutrients, or food groups in general adult populations. Twenty-five studies^3,24,29,39,41,44,49–51,53,59,64–67,69,70,75,76^ assessed dietary intake only once at baseline using FFQs, while seven studies^40,46,48,52,60,61,68^ conducted repeated FFQs. Four studies^38,45,58,63^ assessed intake only once at baseline using 24-hour recalls, and ten studies^4,36,37,42,43,54–57,77^ collected repeated 24-hour recalls (**Table 2**).

**Table 2.**
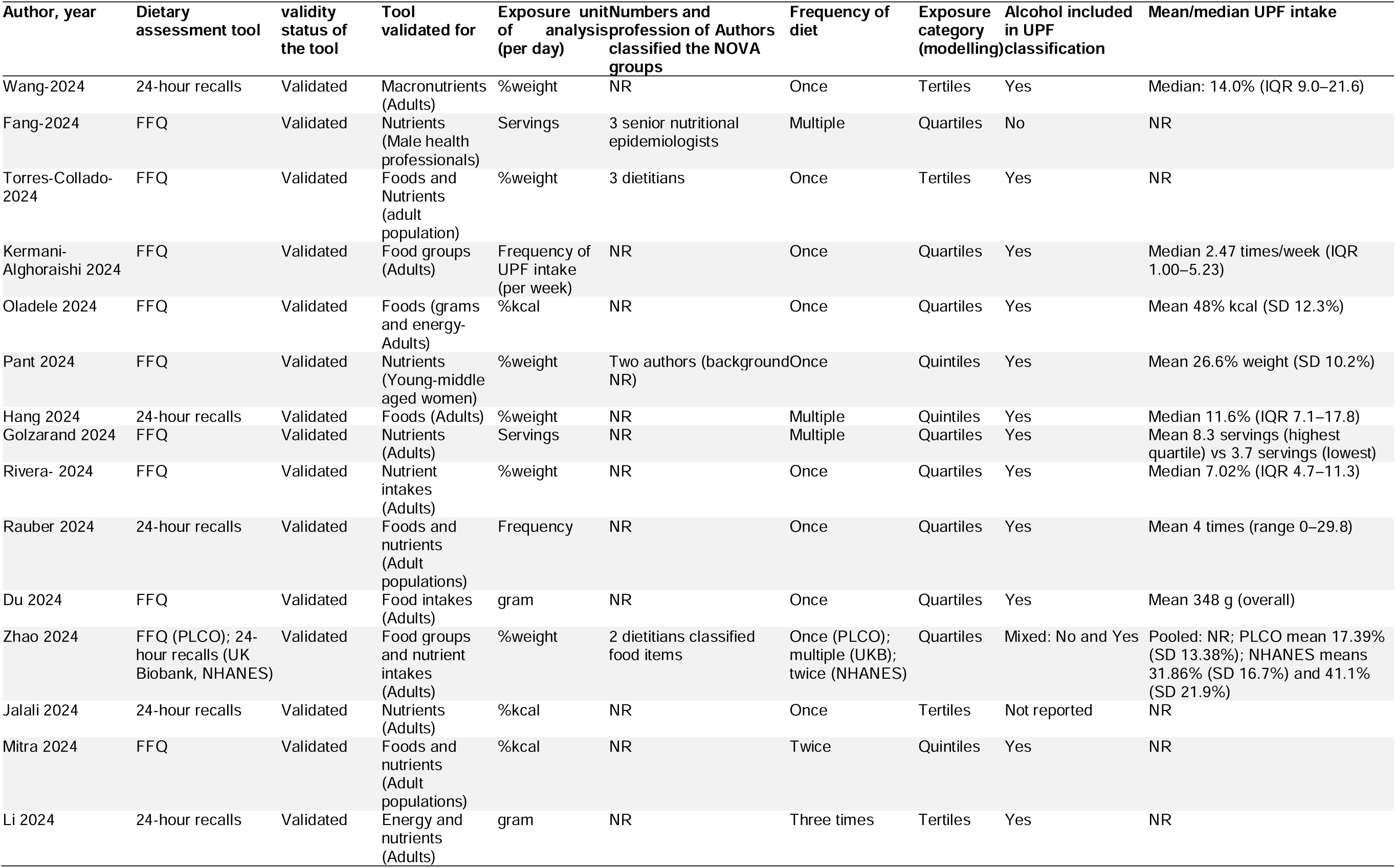
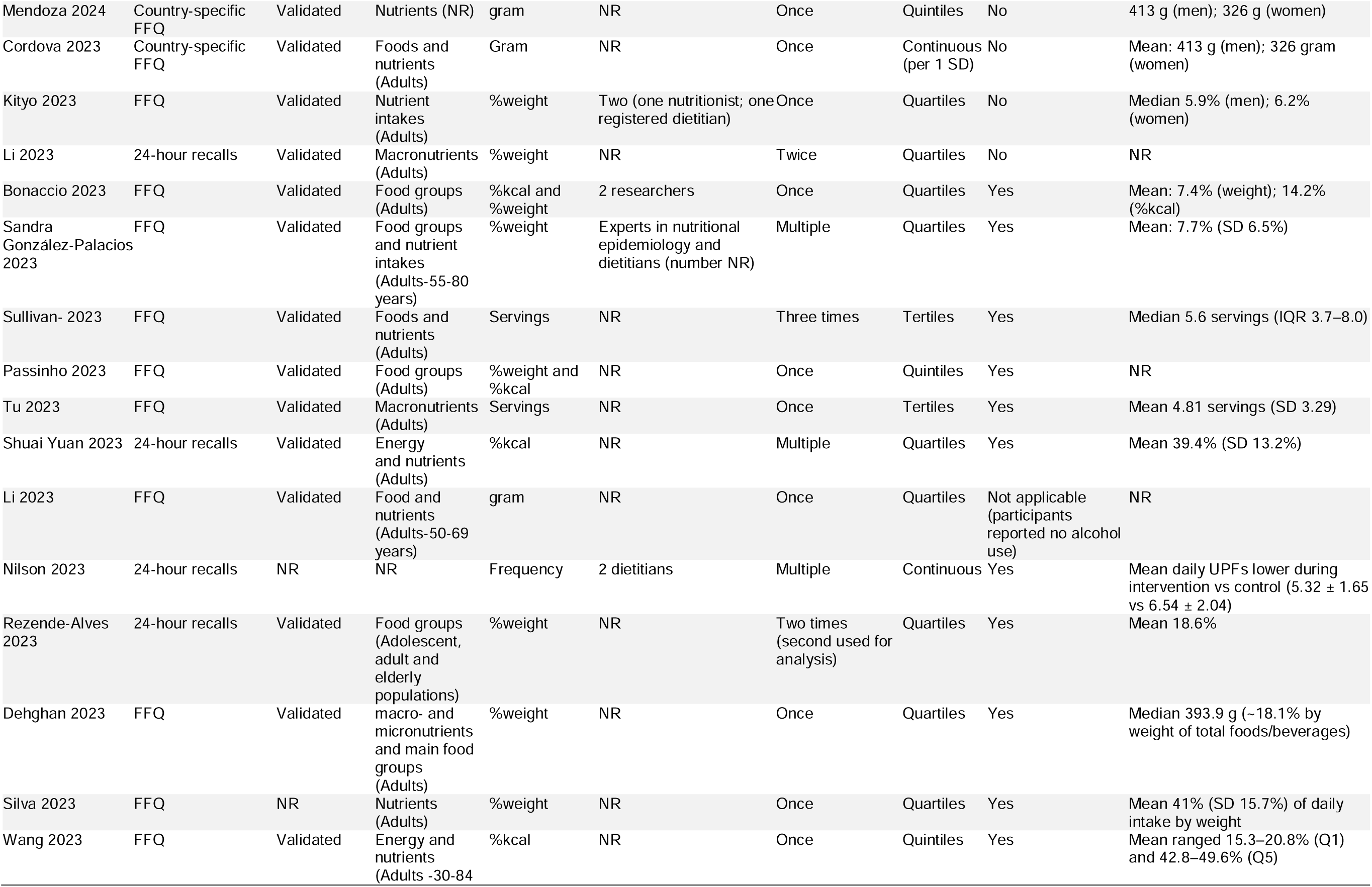
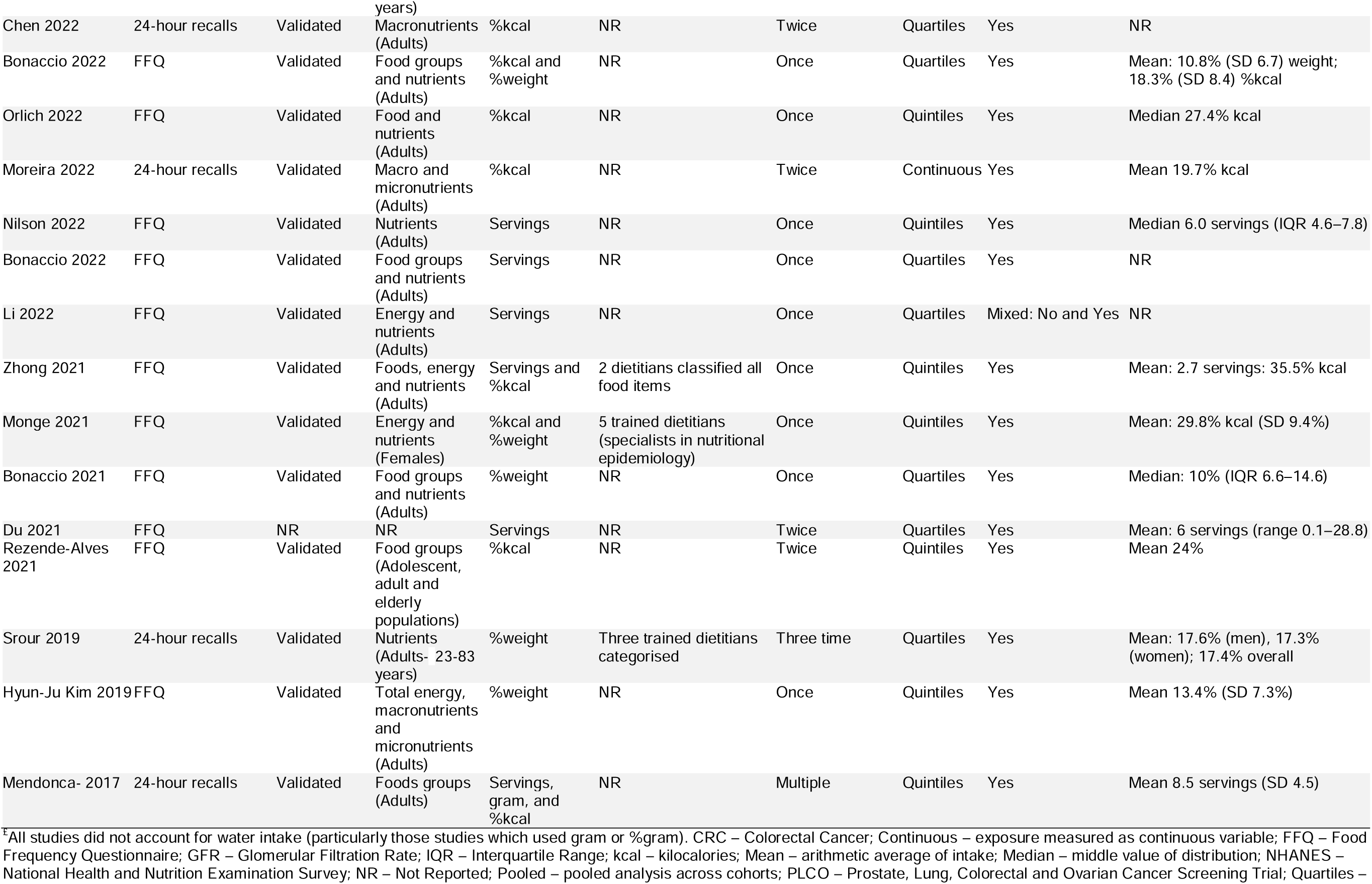
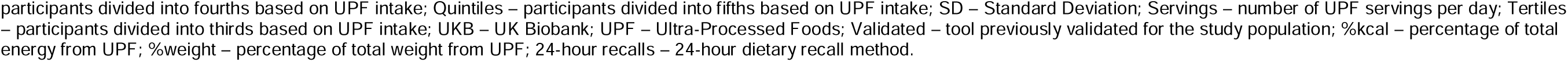
Summary of dietary intake assessment tools and approaches used to assess consumption of ultra-processed foods in prospective studies (n=46)^£^.

### Outcome assessment

Thirty-three studies investigated overall CVD outcomes,^3,4^, Cordova, Viallon, Fontvieille, Peruchet-Noray, Jansana, Wagner, Kyrø, Tjønneland, Katzke, Bajracharya, Schulze, Masala, Sieri, Panico, Ricceri, Tumino, Boer, Verschuren, van der Schouw, Jakszyn, Redondo-Sánchez, Amiano, Huerta, Guevara, Borné, Sonestedt, Tsilidis, Millett, Heath, Aglago, Aune, Gunter, Ferrari, Huybrechts and Freisling ^24,29,41–45,47–53,59,69,76^ 15 studies cerebrovascular diseases,^3,4,37,38,42–44,49,51,55,58,59,62,65,71^ 15 coronary heart disease (including myocardial infarction, ischemic heart disease, coronary artery disease, unstable angina, and atrial fibrillation),^4,37,38,40,42,43,49,54,55,58,59,62,65,71,72^ 10 hypertension,^36,39,46,60,61,66,70,75–77^ and one each venous thromboembolism^56^ and heart failure ^58^ (**Table 3**). Among the included studies, four reported both fatal and non-fatal CVD outcomes,^4,51,55,72^ 19 focused on fatal outcomes (mortality)^3,29,37,38,41,43,44,47–50,57,59,63,64,69,71,73,74^, and 22 reported only non-fatal outcomes (incidence).^24,36,39,40,42,45,52–54,56,58,60–62,65–68,70,75–77^ CVD outcomes were primarily identified through electronic health records (30 studies, **Table 3**), including national registries and death indices coded using ICD. The remaining studies relied on self-reported outcomes or author-assessed definitions. Over a mean follow-up of 12.1 years (ranging from 8 weeks to 34 years), a total of 345,079 CVD events and hypertension were recorded across all studies, corresponding to approximately 46.9 million person-years of observation (**Table 1**).

**Table 3.**
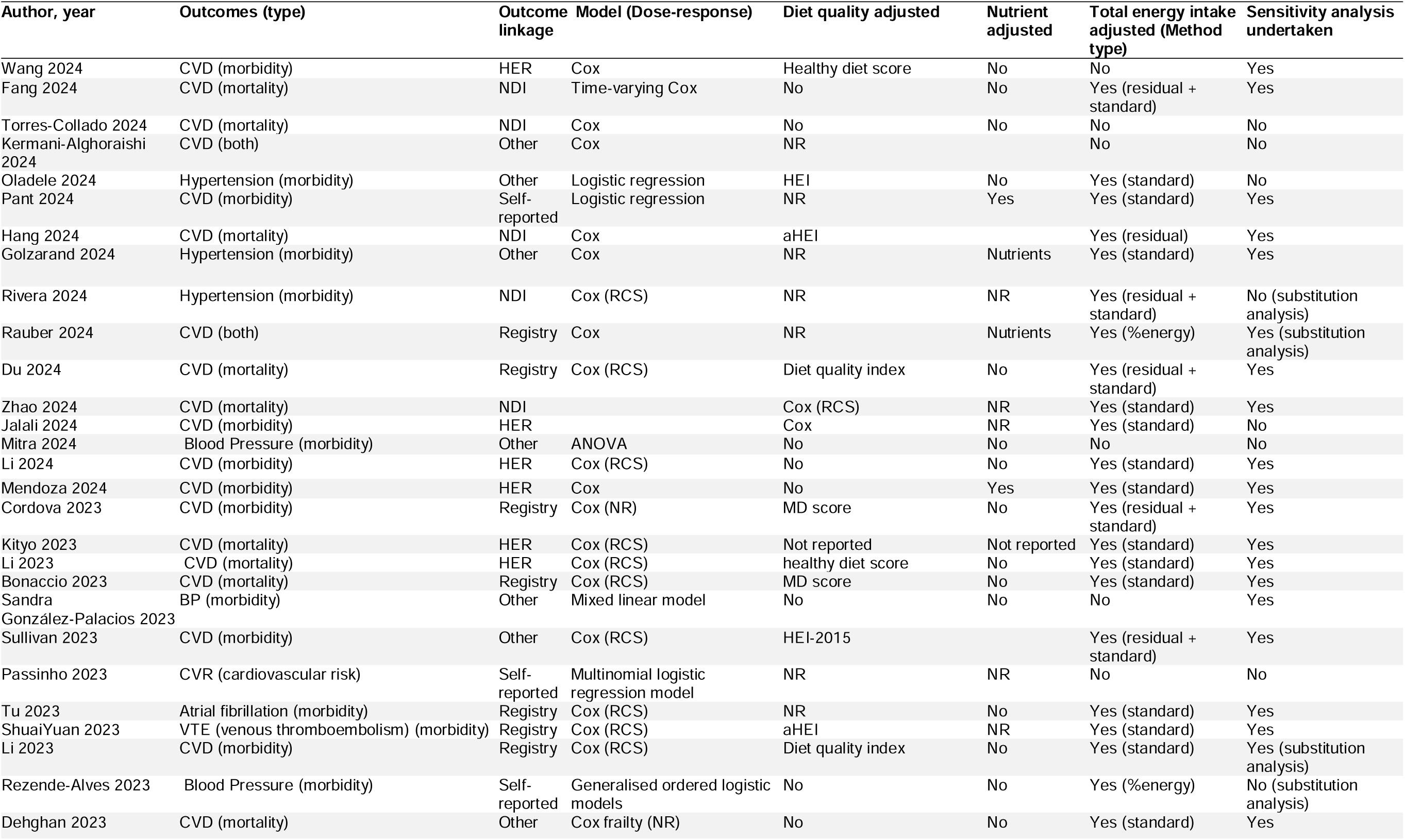
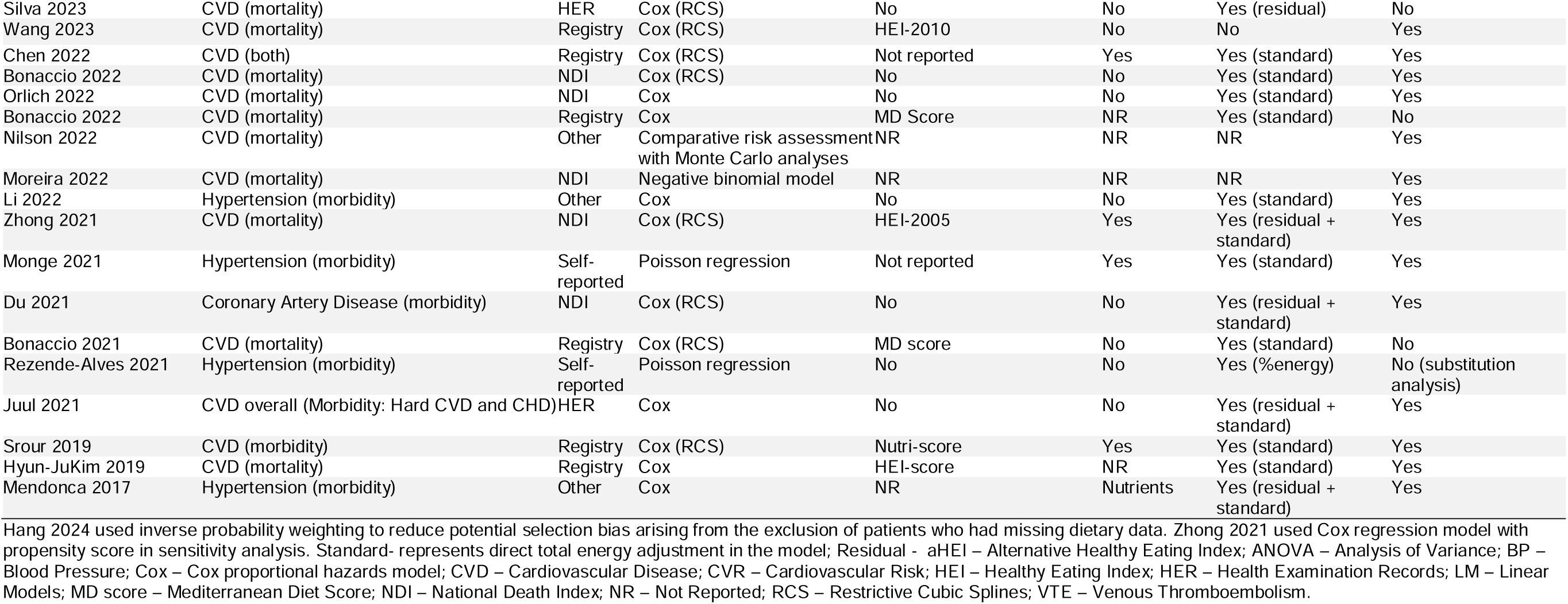
Outcome ascertainment and statistical methods used to identify the relationships between ultra-processed food consumption and cardiovascular disease outcomes in prospective studies (n=46)

## PART II: UPF classification

All included studies applied the NOVA food classification framework. However, only 11 studies (23.9%) reported the number and professional backgrounds of those who classified food items into NOVA groups.^3,36,39,41,42,44,46,48,50,57,76^ Of these, nine reported that classifications were conducted by dietitians or nutritional epidemiologists, while the remaining two did not report the professional background of the authors who classified foods.^44,76^

In all included studies, UPF intake was measured using diverse units (**Table 2**). Seventeen studies used weight ratios (g/day) ^41–46,49,54,57,58,64,65,67,71,76,77^, 11 used calorie contribution (%kcal/day),^3,4,37,38,49,50,55,60–62,69^, and six reported both weight and calorie contributions.^24,39,49,53,56,59^ In addition, UPF intake was assessed as servings/day in 11 studies^3,29,40,48,52,56,66,68,72,73,75^ and as frequency/day in three studies.^36,51,63^ Three studies modelled UPF intake as a continuous variable,^24,37,38^ while 43 studies used categorical approaches (terciles, quartiles or quintiles).^3,4,29,36,39–46,48–77^ No study reported interrater agreement in NOVA classification.

Only six studies^24,41,43,48,57,73^ explicitly excluded alcohol consumption from quantifying UPF consumption. Three Iranian studies^51,66,67^ did not report alcohol consumption, while the remaining studies included alcoholic beverages in UPF classification. None of the studies described how water intake was accounted in weight-based UPF metrics.

When examining the consistency of UPF classification across studies using the same data source, discrepancies were observed. Interrater agreement was very high (95%) in studies using UK Biobank data, high (91%) in Brazilian cohorts, but only moderate (65%) in Iranian studies (**Supplementary Figure 4**). For instance, Du et al.^40^ and Rivera et al.^68^ classified UPF subgroups differently using the same US-based ARIC cohort. Similar inconsistencies were observed among Iranian studies^66,67^, and UK Biobank data analyses.^4,45,55,58^ The most frequently reported UPF subgroups across studies are: sweets and desserts (19 studies), breads and cereals (18 studies), dairy and dairy substitutes (18 studies), meat/processed meat/fish (18 studies), added fats and oils (including Margarine) (18 studies), sugar-sweetened beverages (17 studies), and ready-to-eat/heat mixed dishes (16 studies) (**Figure 1** and **Supplementary Table 4**).

**Figure 1.**
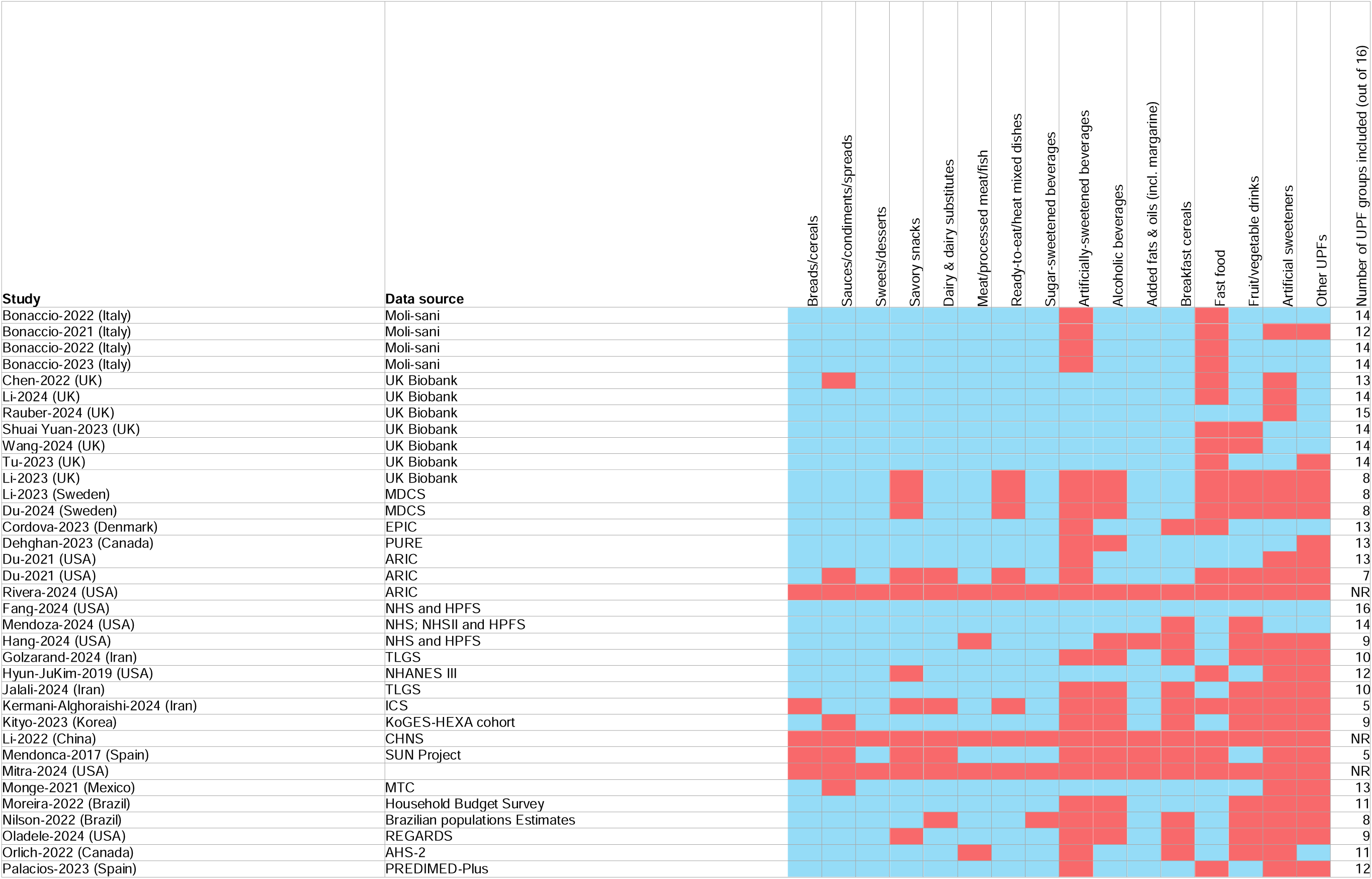
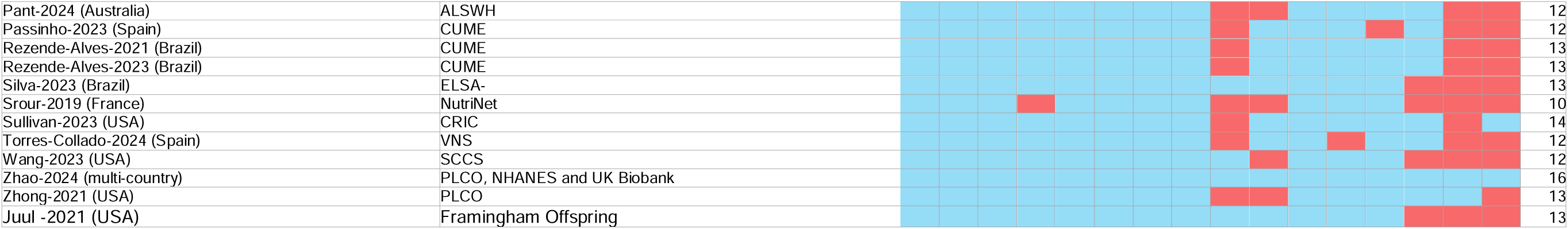
Summary of ultra-processed food groups for each study (n=46)

The average UPF contribution to energy intake (%kcal/day) was highest in the United States (48.0%)^70^ and the United Kingdom (39.4%),^55^ while it was lowest in Italy (10.0%)^49^ (**Supplementary Figure 5**). Similar trend was observed for percentage of dietary weight (% grams/day); the highest UPF contribution was reported in the United States (41.1%),^57^ followed by the UK (31.4%)^57^ and Australia (26.6%),^76^ whereas the lowest contributions were observed in Korea (5.9%)^41^ and Italy (7.0%)^49^ in Italy.

## PART III: Data analysis

### Statistical analysis

Among the conventional statistical models employed to estimate the effect of UPF consumption on CVD and hypertension outcomes, Cox proportional hazard regression model was used in 37 studies (**Table 3**).^3,4,24,29,40–45,48–52,54,59,66,68,69^ Poisson regression^37,39,61^, logistic regression^46,53,60,70,76^ and comparative risk assessment^36,38^ were used in 3, 5 and 2 studies, respectively. Out of all the studies, only one^29^ used the inverse probability treatment weighting, while another^3^ used the Cox regression model with propensity score in the sensitivity analysis. However, 34 (73.9%) of the studies portrayed the robustness of their findings through several sensitivity analyses.^29,37,38,43,54–58,62–64,66,69,71,73,75–77^ Of the 46 included studies, 19 reported using restricted cubic splines to assess dose–response relationships between UPF consumption and CVD outcomes.^40,42–46,48,50,54–56,58,64–66,68,71,72,74^ The units of UPF threshold varied across the study, some used per 10% increase in %grams/day, %kcal/day, or 1 SD increase in grams/day, or per serving/day

### Confounding variables

The covariates considered in the UPF-CVD and -hypertension associations were mainly grouped as socio-demographic, behavioural, clinical and dietary factors (**Figure 2**). Among confounders of UPF-CVD associations, age in all studies, except in five studies,^37,42,55,62,67^ sex in all studies, except in three studies,^37,42,67^ race/ethnicity in 20 studies,^3,4,39,40,43,45,48,52,54–58,60–63,68,69,71^ marital status in 13 studies,^3,40,41,48,51,53,57,60–62,69,71,76^ smoking status in all studies, except in four studies,^37,42,53,67^ physical activity level in all studies, except in three studies,^37,42,50^ BMI in 28 studies,^3,4,24,41,44,45,47–49,51,52,54,56,57,66,68–70,75,76^ and alcohol consumption in 25 studies^3,4,24,29,40,41,43,45,48,54,56,57,60,61,63–65,69,70,75,76^ were adjusted in the final models. Even though majority of (73.9%) the studies supported their main findings with sensitivity analyses, several studies accounted for mediator variables in their final models such as baseline hypertension^3,4,39,43–46,49–52,57,58,60,61,65,75,76^ and diabetes.^3,4,44–46,48–52,57,58,60–62,65,68,69,71,73,74,76^. BMI, overall diet quality^24,43–45,49,52,70,71^ and waist circumference^50,72^ were included.

**Figure 2.**
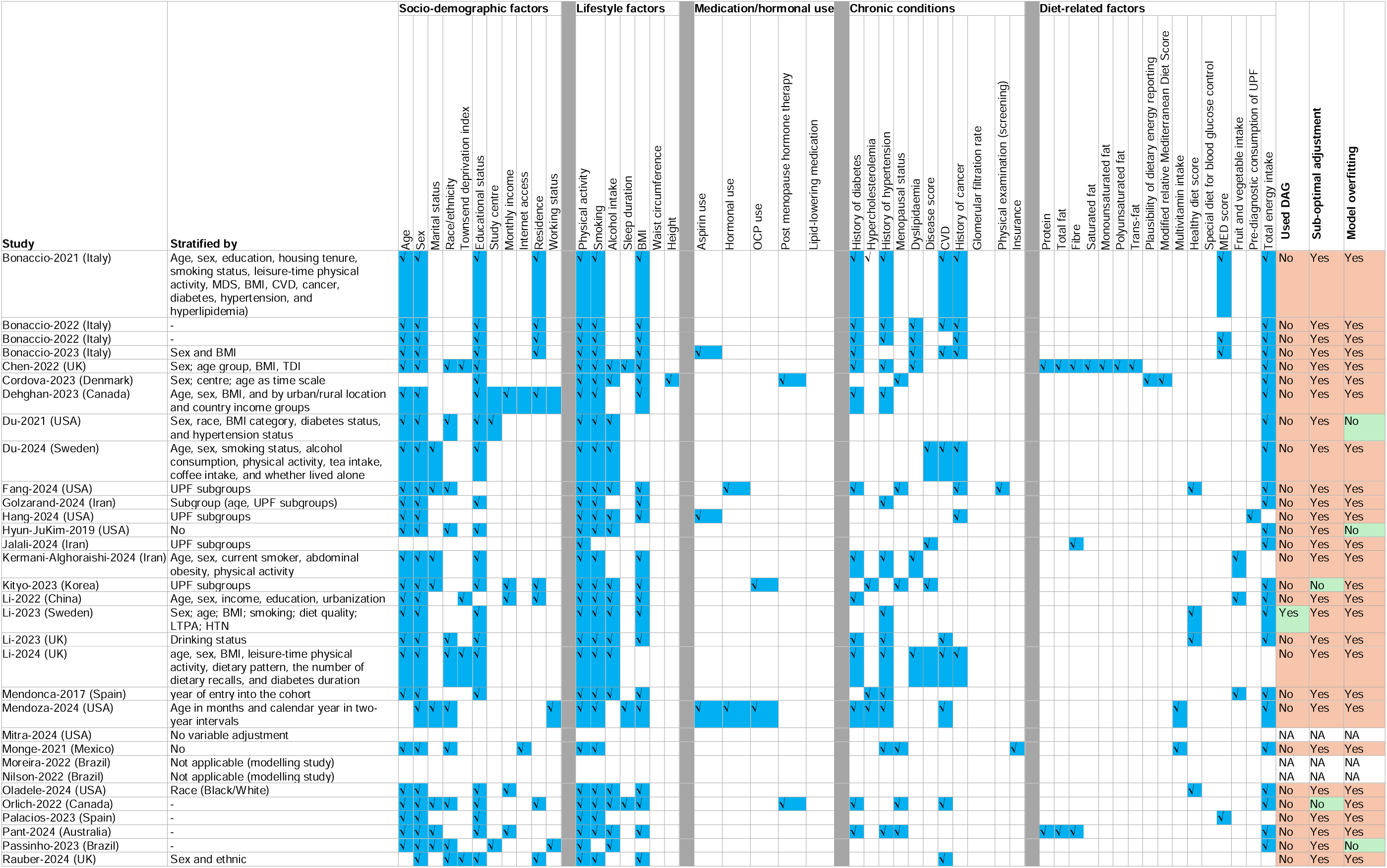
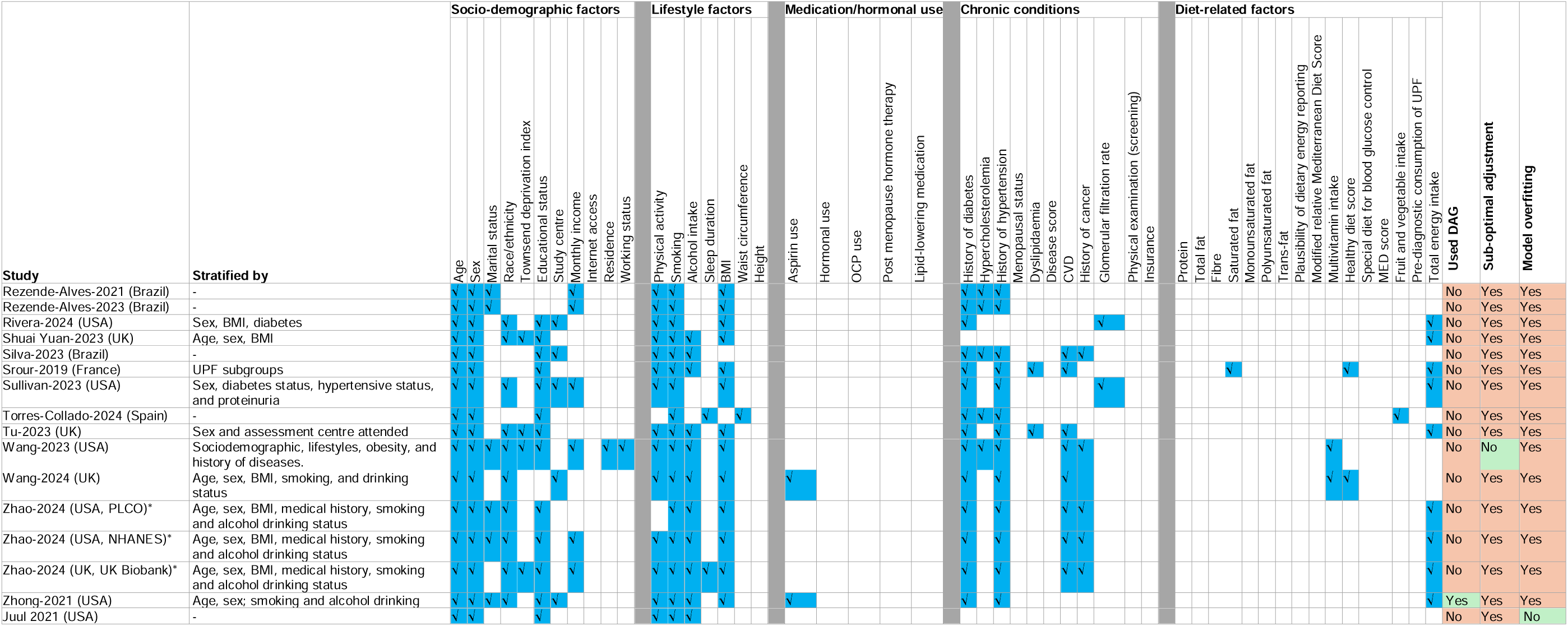
Confounders and covariates considered in association between ultra-processed food consumption and CVDs outcomes (n=46).

We assessed total dietary energy adjustment in all included studies. Dietary energy intake was accounted for in 37 studies,^3,4,24,29,39,41–44,47–49,52,54–70,72–77^ while seven studies did not adjust for it.^36,45,46,50,51,53,71^ Among the 37 studies that adjusted for energy intake, 23 used the standard model (direct adjustment for total energy),^4,39,41–44,47,49,54,56–59,62,63,65–67,69,70,73,76,77^ nine applied both the residual and standard methods (UPF intake first energy-adjusted using the residual method and *at the same time total energy also included in the final model*),^3,24,40,48,52,64,68,72,75^ three used the nutrient density approach (% of total energy intake),^55,60,61^ and two studies used the residual method only.^29,74^

Only one study used the e-value technique to account for unmeasured confounders^3^ while two studies used DAG to select confounding variables. ^3,29^ Overall, sub-optimal adjustment and model overfitting were each identified in 38 studies (82.6%) (**Figure 2**). A Cox frailty model was used in one study^73^ and time-varying Cox regression model was used in another (**Table 3**).^48^

## PART IV: Effect estimates

After excluding studies with potential sample duplication (i.e., those using the same data sources or cohorts), effect estimates (RRs) from 28 studies were pooled across studies, comprising 3,536,142 participants with a mean follow-up of 14.2 years (range: 2–29 years) (**Supplementary Table 5**). Estimates were pooled from 21, 9, 8 and 10 studies for the association between UPF consumption and overall CVD, CHD, cerebrovascular disease and hypertension outcomes, respectively. Those in the highest UPF intake category had a significantly higher risk of CVD and hypertension outcomes compared with those in the lowest category in 13 studies, with RRs ranging from 1.05 to 1.68. In the random-effects model, participants in the highest category of UPF consumption compared to the lowest had a 9% increased risk of CVD outcomes (RR = 1.09; 95% CI: 1.05–1.12 (**Figure 3**). The effect of UPF was found to be heterogenous on CHD (RR=1.19; 95% CI: 1.05, 1.34; **Figure 4 A**), cerebrovascular disease (RR=1.08; 95% CI: 1.01, 1.16; **Figure 4 B**), and hypertension (RR=1.16; 95% CI: 1.08, 1.26; **Figure 4 C**). No difference on CVD effect estimates was found when studies used proportion of weight or energy intake (**Supplementary Figure 6**). The pooled estimate was more attenuated among studies that applied both residual and standard methods compared with those using the standard model only (p = 0.009; **Supplementary Figure 7**). In contrast, no significant subgroup difference was found for the UPF–hypertension association by energy adjustment method (**Supplementary Figure 8**).

**Figure 3.**
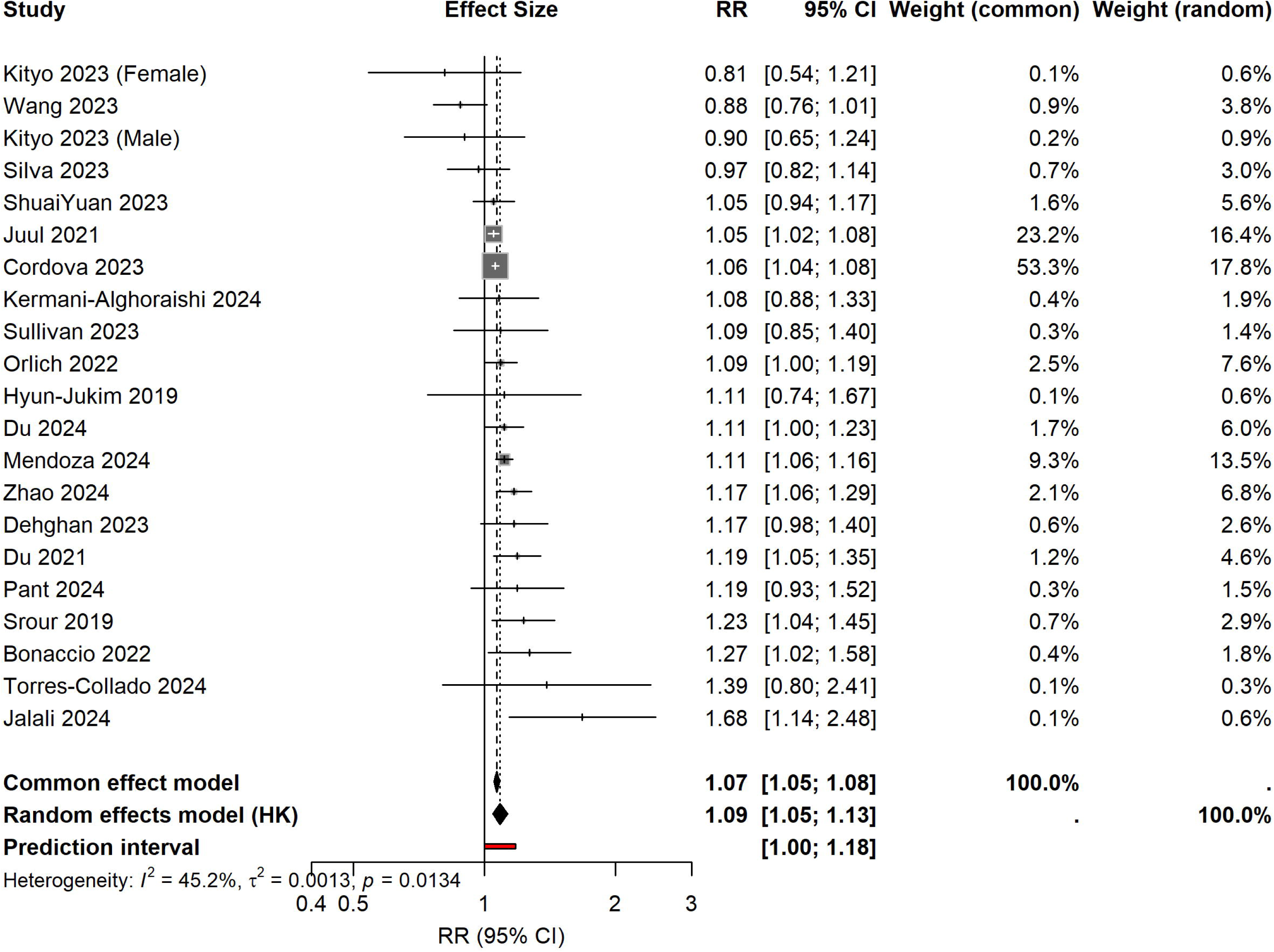
Pooled effect of ultra-processed food on cardiovascular incidence and mortality (n=21)

**Figure 4.**
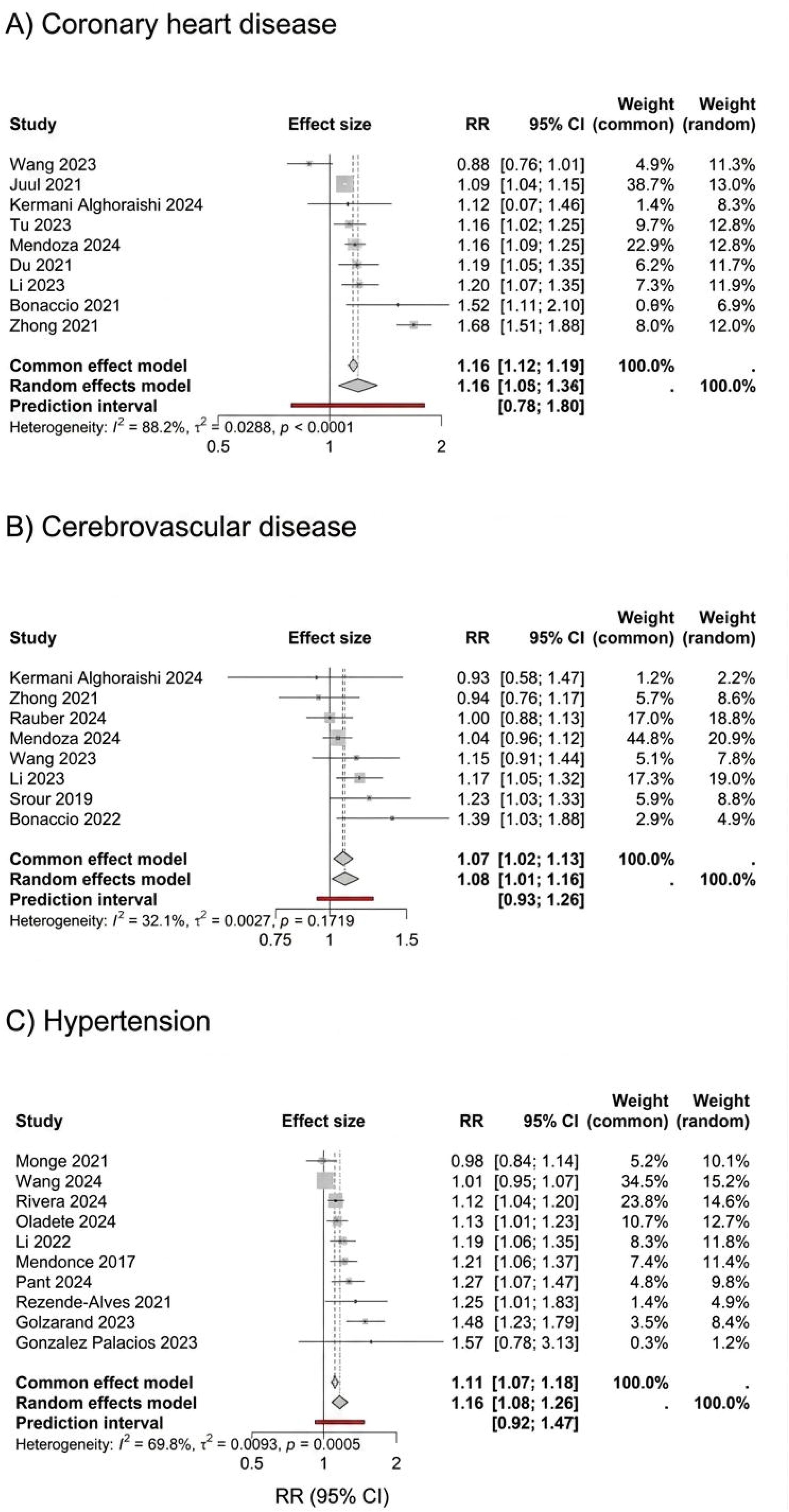
Pooled effect of ultra-processed food on coronary heart disease (n=9), cerebrovascular (n=8) and hypertension (n=10)

The dose–response relationship between UPF consumption and CVD persisted when exposure was measured either as absolute weight (g/day), relative energy contribution or both (% total energy intake) (**Supplementary Figure 9, 10, and 11**). This dose–response association was evident for coronary heart disease and hypertension (**Supplementary Figures 12 and 13**), but not for cerebrovascular disease (**Supplementary Figure 14**).

### Source of Heterogeneity

In the random-effects model, moderate between-study heterogeneity was observed (I² = 44.2% [6.6–66.6%]; H = 1.34 [1.03–1.73]; p = 0.016) (**Figure 3**). Meta-regression showed that the association between UPF consumption and CVD outcomes did not vary by sex, region, dietary assessment tool (FFQ vs. 24-hour recall), UPF unit (calorie vs. weight), CVD outcome type (overall CVD, cerebrovascular disease, CHD, hypertension), event type (morbidity vs. mortality), or follow-up duration (<10 vs. ≥10 years) (**Supplementary Tables 6**). However, energy adjustment method significantly explained heterogeneity in the association between UPF consumption and CVD risk (QM = 8.23, df = 3, p = 0.042). The included moderators accounted for nearly all between-study heterogeneity (R² = 99.98%), reducing residual heterogeneity to virtually zero (I² = 0.01%, τ² ≈ 0). Specifically, studies that used the standard energy adjustment model (direct inclusion of total energy as a covariate) reported significantly higher effect estimates compared to those using other approaches (RR = 1.04, 95% CI 1.002–1.07, p = 0.038) (**Supplementary Table 6, and Supplementary Figures 15 and 16).**

Visual inspection using funnel plot (**Supplementary Figure 17**) and statistical tests using Egger’s (p = 0.187) and Begg’s tests (p = 0.904) suggested no asymmetry. Using the trim-and-fill method, the association remained statistically significant after adjustment for potential missing studies (RR = 1.07, 95% CI: 1.04–1.11; **Supplementary Figure 18**).

Leave-one-out analyses produced consistent estimates (RRs 1.08–1.09; 95% CIs 1.05–1.14) across all models (**Supplementary Figure 19**), indicating robustness, with no single study disproportionately influencing the pooled effect. No evidence of publication bias was detected for CHD (Egger’s test p = 0.540; **Supplementary Figure 20**) or cerebrovascular disease (p = 0.464; **Supplementary Figure 21**). However, there was marginal evidence of publication bias for hypertension (p = 0.045; **Supplementary Figure 22**).

### Risk of bias and study quality

According to the ROBINS-E tool, 35 of the 43 included cohort studies were rated at high risk of bias, five at some concerns, and three at very high risk of bias (**Supplementary Figure 3**). Because most studies were judged to be at high risk of bias, we did not pool results according to the overall risk-of-bias rating. However, subgroup analyses were performed according to key ROBINS-E domains (confounding, exposure measurement, and selection of participants) (**Supplementary Figures 23, 24, 25 and 26**). Although tests for subgroup differences were not statistically significant, the pooled risk ratio for CVD events was slightly attenuated among studies with high or some concerns regarding bias in exposure measurement and participant selection, compared with those at low risk of bias (**Supplementary Figures 24 and 25**). In contrast, the association with hypertension was stronger in studies judged at high risk of bias due to selection of participants (**Supplementary Figure 26**).

According to the GRADE evaluation, the overall certainty of evidence linking UPF consumption with CVD, CHD, cerebrovascular disease, and hypertension was judged to be very low (**Supplementary Table 3**). This indicates that the observed associations are highly uncertain related to risk of bias raised from the seven domains of ROBINS-E criteria that are bias raising due to confounding, exposure measurement, selection, outcome ascertainment, missing data, post-exposure intervention and selection of reported result among the included studies.

## Discussion

This review provided a systematic and critical evaluation of the methodologies underpinning the evidence linking UPF intake with CVD by focusing on data collection, UPF classification, statistical analysis, covariate selection, and effect estimates. Methodologically, the cumulative evidence base is large and increasingly multinational, but it is characterized by significant use of one-time self-reported dietary data, diverse use of NOVA classification, predominantly conventional regression modelling with sparse causally specified models, heterogenous variable adjustment sets, and widespread adjustment for potential mediators. Nevertheless, our meta-analysis found an overall 9% and 16% higher CVD and hypertension risk for the highest versus lowest UPF intake, respectively, consistent dose–response relationships, and elevated risks for specific CVD subgroups (CHD and cerebrovascular disease). The findings appear robust across sensitivity, trim-and-fill, and leave-one-out analyses, and align with the most recent syntheses and guidance.^2,9,18,78^ However, these associations may have been attenuated due to over-adjustment in the majority of studies (n = 38, 82.6%), including adjustment for potential mediators. Although this raises the possibility that the true effect of UPF consumption on CVD and hypertension risk could be stronger than observed, the direction of bias remains uncertain given the substantial heterogeneity in measurement errors.^79^ Subgroup meta-analyses showed that studies that applied both residual and standard energy adjustment methods, as well as those judged at high risk of bias in domains related to exposure measurement and participant selection, showed more attenuated UPF-CVD associations compared with studies using the standard adjustment method alone or rated at lower risk of bias.

### Data collection

Most studies relied on self-administered FFQs, or 24-hour recalls collected at a single baseline time point. Such designs are prone to measurement error and do not capture the temporal dynamics of dietary behaviour, which tends to bias associations by providing conservative risk estimates.^80^ Established approaches, such as periodic dietary assessments and regression calibration using unbiased reference instruments (e.g., multiple 24-hour recalls), can strengthen the robustness of effect estimates,^81^ but their application was very limited across the studies included in this study. To strengthen the evidence base in UPF and cardiovascular research, repeated dietary assessments across follow-up and protocolised strategies for handling misreporting of dietary intake (e.g., sensitivity analyses) would be essential for improving the validity and generalisability of effect estimates. These practices align with contemporary dietary assessment guidance and can substantially enhance the robustness of UPF research.^80,81^

Beyond generic challenges in dietary data collection and measurement error, UPF-specific ascertainment introduces additional complexity. Accurately distinguishing “industrially manufactured” products from “culinary preparations”, as well as capturing brand- and formulation-level details, is challenging with FFQs and not assured with recalls, where brand reporting and label information are often incomplete.^11^ Older dietary databases often predate the NOVA classification and lack the metadata needed to determine industrial processing or to capture the presence and function of hallmark additives. This gap forces investigators to make assumptions that vary across studies and undermine comparability as found in the current review. Looking forward, future cohorts should incorporate NOVA-relevant prompts at data collection (e.g., brand, purchase source, packaged versus home-made flags), adopt technology-enabled tools (e.g., photo-assisted or receipt-linked recalls, barcode/app-based scanners), and maintain versioned linkages to dynamic food databases to ensure evolving formulations remain traceable over time.^82^ Furthermore, given the resource burden of high-granularity diet collection, hybrid designs can balance feasibility and validity, including sparse, routine FFQs complemented by periodic multi-pass recalls for calibration and sub-studies with intensive brand/label capture to build transferable coding dictionaries.^82^ Embedding these elements prospectively, with transparent documentation, will standardise exposure ascertainment across settings and yield more reproducible, policy-relevant estimates of UPF–CVD associations.^11,81^

### UPF classification

All studies included in the current review applied the NOVA classification; however, only 26% reported who performed the categorisation, and none reported inter-rater agreement, an omission that undermines reproducibility and complicates cross-study comparisons. Recent methodological work demonstrates that, with structured coder training and transparent decision rules, high reliability in assigning NOVA categories to 24-hour recall items is achievable.^83^ This requires explicit reporting and the use of sharable dictionaries or codebooks to standardise practice across studies. Building on published best practices for NOVA deployment, a minimum reporting set should include coder qualifications, training materials and decision trees, inter-rater reliability metrics (e.g., Cohen’s κ^84^), publicly available codebooks with exemplars, and pre-specified sensitivity analyses for items with uncertain classification.^85^ Adoption of these standards would enhance the comparability of UPF exposure across cohorts and help mitigate classification heterogeneity.

Cross-study inconsistencies we documented, such as including versus excluding alcohol in UPF classification (given the strong association between alcohol consumption and CVD^86^) and overall heterogeneous subgroup mapping of UPFs, are not trivial. These differences can bias absolute intake estimates and hinder comparability across settings and effect estimates on CVD. We recommend standardising the treatment of alcohol (exclude or model separately, with justification) and reporting a harmonized minimal set of UPF subgroups with common exemplars. These align with recent advisories and methodological commentaries that emphasise transparent, standardized application rather than abandoning processing-based information.^85,87^

### Statistical methods and covariates

Cox models were the most commonly used statistical method to determine the associations between UPF and CVD. UPF exposure, like other lifestyle factors, is dynamic and complex;^88^ it interacts with other behaviours, metabolic and health conditions, and evolves over time, creating time-varying confounding (e.g., BMI, which is itself influenced by prior UPF intake). Recent developments in causal inference for observational studies, such as target trial emulation and advanced causal statistical methods (e.g., g-methods), can appropriately address time-varying confounding and could improve the validity of findings.^89^ These causal frameworks provide clear templates for implementing advanced tools in longitudinal nutrition cohorts, and broader adoption would substantially improve the validity and interpretability of UPF–CVD associations.^89^ However, modern causal analysis approaches were rarely applied in the studies included in this systematic review, despite their increasing use in nutritional epidemiology.^90,91^

Covariate selection requires clear articulation of the intended estimand. Common covariates may lie on the causal pathway between UPF and CVD, meaning that routine adjustment can attenuate total-effect estimates through over-adjustment. In the current review, we recognized that substantial disagreement exists regarding the most appropriate adjustment sets; and only two studies^3,29^ used DAG to select confounding variables. For example, some studies considered BMI, overall diet quality, lipid profile and other metabolic markers primarily as mediators on the pathway between UPF consumption and CVD, while others viewed them as a confounder or collider. Similarly, certain modelling strategies that we classified as potentially over-adjusting (e.g., adjusting for total energy intake) could be considered appropriate under different causal estimands. We therefore caution against interpreting our appraisals as definitive judgments.

Depending on the research question (total versus direct effect), energy-partition or isocaloric substitution models, where UPF is replaced with minimally processed alternatives, are preferable to unconditional energy adjustment and should be complemented by sensitivity analyses to quantify potential over-adjustment. This perspective is supported by experimental evidence showing that UPF patterns increase ad libitum intake independent of macronutrient targets, and it aligns with current advisory recommendations to specify modelling choices a priori.^78,87,89^

Substantial methodological heterogeneity was observed in how studies adjusted for total energy intake. Meta-regression revealed that the energy adjustment method was a significant source of between-study variation, explaining nearly 100% of the heterogeneity (*R*^2^ = 99.98%). Studies using the standard model (direct inclusion of total energy as a covariate) reported stronger associations between UPF consumption and CVD risk compared with other approaches. This finding aligns with the ongoing debate in nutritional epidemiology regarding different energy adjustment strategies, which estimate distinct causal estimands: the standard and residual models assess isocaloric substitution effects, the nutrient density model evaluates diet composition, and the energy partition (all-components) model estimates total (non-isocaloric) effects.^26,28^ As highlighted by Tomova et al., this variation can lead to systematically different effect estimates.^26^ While Willett et al. argue that the partition model is often misaligned with the isocaloric substitution question most relevant in nutritional epidemiology,^27^ the ongoing debate underscores the importance of carefully considering energy adjustment methods when interpreting and synthesising evidence on diet and health outcomes. Greater transparency and consistency in energy adjustment methods are therefore needed to improve the interpretability and synthesis of evidence in this field.^28^

### Effect estimates

Our pooled estimates indicate a modest but consistent elevated CVD risk with higher UPF intake, showing graded increases across exposure categories and concordant patterns whether UPF was expressed as %kcal or %gram. These findings align with recent dose–response and multi-cohort syntheses that reported increased risks for composite CVD and CHD, with broadly similar magnitudes and directionality.^9,18^ Our meta-regression showed that associations did not differ materially by sex, region, dietary assessment instrument, exposure metric, event type, or follow-up length, suggesting that the heterogeneity observed across studies cannot be attributed to these features alone. Standardising exposure construction and applying causal, time-varying exposure modelling approaches should be developed and employed to minimise bias and strengthen future evidence. Moreover, this suggests the importance to assess other sources of heterogeneity, such as background diet.

Mechanistic plausibility further supports a causal contribution. In tightly controlled crossover RCTs, ultra-processed diets increased *ad libitum* energy intake and weight despite macronutrient-matched menus, implicating food matrix effects, palatability, and eating rate as proximal drivers.^78,92^ Large prospective cohorts have also linked specific UPF constituents and additives (e.g., emulsifiers) with higher CVD risk, consistent with inflammatory and metabolic signalling pathways.^93^ While mechanisms are multifactorial and not fully understood, triangulation across experimental, cohort, and meta-analytic evidence has strengthened the evidence that elevated UPF intake is deleterious for cardiometabolic health.^2,9,18,78,92,93^

### Potential mechanisms

UPF may increase the risk of hypertension and cardiovascular disease through multiple interconnected biological and behavioural pathways. UPF are typically high in added sugars, sodium, and saturated fats, while low in fibre and protective micronutrients, promoting insulin resistance, endothelial dysfunction, and elevated blood pressure.^62,68,94–96^ Their hyper-palatable nature strongly activates reward-related neural pathways, leading to overconsumption and positive energy balance.^97^ Ultra-processing also alters the food matrix, reducing satiety signalling and increasing eating rate.^98^ Furthermore, food additives such as emulsifiers and non-nutritive sweeteners, along with neo-formed contaminants, can disrupt gut microbiota composition, increase intestinal permeability, and induce low-grade systemic inflammation, all of which contribute to hypertension and atherosclerosis.^99,100^ These mechanisms collectively provide biological plausibility for the observed associations between high UPF consumption and adverse cardiometabolic outcomes.

### Implications

We recommend establishing a field standard for UPF epidemiology that includes: (i) repeated dietary assessments with calibration to reduce measurement error; (ii) transparent, sharable NOVA codebooks with coder training details and inter-rater reliability metrics; (iii) causal inference framework (e.g., DAGs, target-trial emulation and robust causal statistical analysis methods) with explicit estimands;^89^ and (iv) substitution models to quantify the health impacts of replacing UPF with minimally processed alternatives. These practices, grounded in established dietary assessment resources and causal inference frameworks, would substantially enhance causal interpretability and reproducibility.^81,85,89^ Additionally, a major challenge in UPF research is accurate exposure assessment. Most studies, including those in this review, rely on dietary instruments not designed to capture food processing level, often leading to potential misclassification. As highlighted by Kuhnle, FFQs and 24-hour recalls typically lack ingredient-level detail, forcing assumptions about recipes and brands.^101^ Future research should prioritise the development and validation of UPF-specific dietary assessment methods to strengthen the evidence base for policy recommendations.

Furthermore, given the uncertainty surrounding certain causal structures in this field, future studies could benefit from multiverse-style sensitivity analyses or specification curve analyses. Such approaches would systematically explore how effect estimates change across a range of plausible modelling decisions, thereby providing a more comprehensive picture of the robustness of observed associations.

Until robust methodologies specifically designed to assess UPF intake are widely adopted, caution is warranted when translating the existing evidence into policy recommendations aimed at reducing UPF consumption. In light of this, the 2025 American Heart Association Science Advisory synthesises existing science and highlights policy levers, including procurement standards, marketing and labelling regulations, and reformulation targets, while underscoring the parallel need for classification standards and mechanistic research.^87^ Our findings reinforce these recommendations and further suggest that benefits may be underestimated, given the likely attenuation from measurement error and over-adjustment in existing studies.

## Conclusion

By systematically interrogating four methodological domains, we demonstrate that UPF–CVD and -hypertension associations are consistent, dose-responsive, and robust. However, prevailing methodological challenges (e.g., single-time-point diet assessment, classification variability, and adjustment for mediators) warrant cautious interpretation and underscores the need for repeated dietary measurements, harmonized food classification systems, and advanced analytical strategies to strengthen causal inference. Moreover, the magnitude of these associations is notably influenced by the method of energy adjustment employed. As different approaches estimate distinct causal estimands, inconsistent use of these methods contributes to heterogeneity and complicates interpretation. Until greater methodological consistency is achieved in energy adjustment strategies and UPF-specific exposure assessment, the existing evidence should be interpreted with caution.

Standardizing UPF classification systems and exposure definitions, alongside the broader adoption of modern causal inference methods, will be essential to sharpen effect estimates and reduce the heterogeneity observed across existing studies. In the interim, reducing UPF consumption and encouraging substitution with minimally processed foods remain prudent evidence-based strategies for protecting cardiovascular health.^87,92^

## Data Availability

No primary data were used for analysis in this manuscript, and all associated data are available within supplementary materials.

## Author contributions

TCM: conceptualisation; methodology; formal analysis; investigation; data curation; writing – original draft; and visualisation; ZAA: conceptualisation; methodology; data curation; writing – review and editing; AMD: data curation and writing – original draft; TT, TU, KD, ZS, RA, ML, JHW, and MS: methodology; writing – review and editing; visualisation; CB: literature searching; writing – review and editing; YAM: conceptualisation; methodology; validation; writing – original draft (Discussion section); review and editing; visualisation; funding acquisition; and overall supervision of the project.

## Data availability statement

This manuscript did not involve the collection or analysis of primary data. All data underlying the findings are derived from previously published studies and are available within the supplementary materials, which include the R code used to conduct the meta-analyses.

