## Supplementary File for "A systematic review to critically appraise methodological rigour in research on ultra-processed food and cardiovascular disease and hypertension"

**Supplementary Table 1.** Description of inclusion and exclusion criteria for selecting relevant articles that assessed UPF consumption and cardiovascular outcomes

| Category | Inclusion criteria | Exclusion criteria |
| --- | --- | --- |
| Population (study participants) | Human participants:  Male and/or Female participants | Studies using animal model |
| Intervention/Exposure | Studies that examined the relationship between ultra-processed food (UPF) intake and cardiovascular diseases.   - UPF measured in absolute intake (grams or energy), proportion (%weight or caloric ratio), frequency or number of serving per day. | Studies that   - with only single food group of UPF |
| Comparator | Studies that used less processed or minimally processed food or lowest UPF intake as a reference. | - |
| Outcomes | Studies that reported incidence or mortality of Cardiovascular events during specified period (coronary artery/heart disease, cerebrovascular disease, peripheral artery disease, and a*ortic atherosclerosis)*. | Studies that did not indicate whether participants in the cohort or trial free from CVD events at baseline. |
| Study Design | - Randomized controlled trials - Quasi-experimental and controlled before and after studies - Prospective cohort studies | - Uncontrolled trials - Uncontrolled before and after studies - Case-control studies - Cross-sectional studies - Narrative reviews - Systematic reviews/meta-analysis |
| Date of publication | - | - |
| Language of publication | - | - |
| Age of participants | Age at exposure/intervention:   - Adults aged 18 years and above.   Age at outcome:   - Adults above 18 years. | Age at exposure/intervention:   - Participants age less than 18 years   Age at outcome:   - Participants age less than 18 years |
| Health status of the participants | Studies that included/enrolled participants who were healthy and or at risk for CVDs.  Studies that also included some individuals who were diagnosed with the diseases for CVD mortality outcome. | Studies that exclusively enrolled participants who were diagnosed with a disease or injured. |
| Publication status | Studies that had been peer-reviewed and published in peer-reviewed journals. | Works that had been not peer-reviewed, published in predatory journals, unpublished manuscripts, pre-prints, reports, abstracts and conference proceedings |
| Country of studies | - | - |

**Supplementary Table 2.** Search strategies for studies examined the association between ultra-processed food consumption and CVD and hypertension outcomes

| Methodological variations in ultra-processed foods assessment and its implication on effect estimates on Cardiovascular outcomes: A Systematic Review and Meta-Analysis | | | | | |
| --- | --- | --- | --- | --- | --- |
| Medline |  |  | 1 | Fast Foods/ | 3477 |
|  |  |  | 2 | Food, Processed/ | 462 |
|  |  |  | 3 | ("Ultra-processed food*" or "Ultra processed food*" or "ultraprocessed food*" or "ultra-processed product" or "fast food*" or "processed food*" or "food, processed" or "processed meat" or "artificially sweetened beverage*" or "sugary drink*" or "carbonated drink*" or "carbonated beverage*" or "food additive*" or "energy drink*" or "ready-to-eat" or chocolate or candy or cookies or "food processing" or "sugar-sweetened beverage*" or "sugar sweetened drink*" or "food additive*" or "fermented beverage*" or "frozen food*" or "meat substitute*").tw,kf. | 51933 |
|  |  |  | 4 | 1 or 2 or 3 | 52766 |
|  |  |  | 5 | cardiovascular diseases/ or heart diseases/ or atrial fibrillation/ or ventricular fibrillation/ or heart arrest/ or heart failure/ or heart failure, diastolic/ or heart failure, systolic/ or heart rupture/ or myocardial ischemia/ or angina pectoris/ or angina, unstable/ or angina, stable/ or microvascular angina/ or coronary disease/ or coronary artery disease/ or coronary occlusion/ or coronary thrombosis/ or myocardial infarction/ or anterior wall myocardial infarction/ or inferior wall myocardial infarction/ or pulmonary heart disease/ or aortic diseases/ or arterial occlusive diseases/ or cerebrovascular disorders/ or brain ischemia/ or brain infarction/ or cerebral infarction/ or infarction, anterior cerebral artery/ or infarction, middle cerebral artery/ or infarction, posterior cerebral artery/ or hypoxia-ischemia, brain/ or ischemic attack, transient/ or carotid artery diseases/ or carotid artery thrombosis/ or stroke, lacunar/ or intracranial arterial diseases/ or cerebral arterial diseases/ or intracranial aneurysm/ or "intracranial embolism and thrombosis"/ or intracranial embolism/ or intracranial thrombosis/ or sinus thrombosis, intracranial/ or intracranial hemorrhages/ or cerebral hemorrhage/ or intracranial hemorrhage, hypertensive/ or stroke/ or hemorrhagic stroke/ or ischemic stroke/ or embolic stroke/ or thrombotic stroke/ or hypertension/ or peripheral vascular diseases/ | 1494743 |
|  |  |  | 6 | Hypertension/ | 266406 |
|  |  |  | 7 | Peripheral Vascular Diseases/ | 13047 |
|  |  |  | 8 | Stroke Rehabilitation/ or "National Institute of Neurological Disorders and Stroke (U.S.)"/ or Embolic Stroke/ or Ischemic Stroke/ or Stroke Volume/ or Hemorrhagic Stroke/ or Stroke/ | 210090 |
|  |  |  | 9 | Angina, Stable/ or Angina Pectoris/ or Angina, Unstable/ | 42765 |
|  |  |  | 10 | Myocardial Infarction/ | 184060 |
|  |  |  | 11 | Coronary Disease/ or Myocardial Ischemia/ or Heart Failure/ | 320742 |
|  |  |  | 12 | ("cardiovascular disease*" or "Cardiovascular incident" or "Cardiovascular accident" or "Cardiovascular outcome*" or "cardiovascular endpoint" or "cardiovascular event" or "cardiovascular death" or "'major adverse cardiovascular " or "heart attack" or "coronary heart disease" or "cerebrovascular accident" or "ischemic heart disease" or hypertension or stroke* or "peripheral artery disease*" or angina or "myocardial infarct*" or "coronary disease*" or "myocardial ischemia*" or "heart failure").tw,kf. | 1472406 |
|  |  |  | 13 | 5 or 6 or 7 or 8 or 9 or 10 or 11 or 12 | 2147748 |
|  |  |  | 14 | 4 and 13 | 3148 |
| **Embase** 1974 to 2024 November 15 | | | | | 5326 |
|  |  |  | 1 | Fast Foods/ | 11595 |
|  |  |  | 2 | Food, Processed/ | 2006 |
|  |  |  | 3 | ("Ultra-processed food*" or "Ultra processed food*" or "ultraprocessed food*" or "ultra-processed product" or "fast food*" or "processed food*" or "food, processed" or "processed meat" or "artificially sweetened beverage*" or "sugary drink*" or "carbonated drink*" or "carbonated beverage*" or "food additive*" or "energy drink*" or "ready-to-eat" or chocolate or candy or cookies or "food processing" or "sugar-sweetened beverage*" or "sugar sweetened drink*" or "food additive*" or "fermented beverage*" or "frozen food*" or "meat substitute*").tw,kf. | 61812 |
|  |  |  | 4 | 1 or 2 or 3 | 67831 |
|  |  |  | 5 | cardiovascular diseases/ or heart diseases/ or atrial fibrillation/ or ventricular fibrillation/ or heart arrest/ or heart failure/ or heart failure, diastolic/ or heart failure, systolic/ or heart rupture/ or myocardial ischemia/ or angina pectoris/ or angina, unstable/ or angina, stable/ or microvascular angina/ or coronary disease/ or coronary artery disease/ or coronary occlusion/ or coronary thrombosis/ or myocardial infarction/ or anterior wall myocardial infarction/ or inferior wall myocardial infarction/ or pulmonary heart disease/ or aortic diseases/ or arterial occlusive diseases/ or cerebrovascular disorders/ or brain ischemia/ or brain infarction/ or cerebral infarction/ or infarction, anterior cerebral artery/ or infarction, middle cerebral artery/ or infarction, posterior cerebral artery/ or hypoxia-ischemia, brain/ or ischemic attack, transient/ or carotid artery diseases/ or carotid artery thrombosis/ or stroke, lacunar/ or intracranial arterial diseases/ or cerebral arterial diseases/ or intracranial aneurysm/ or "intracranial embolism and thrombosis"/ or intracranial embolism/ or intracranial thrombosis/ or sinus thrombosis, intracranial/ or intracranial hemorrhages/ or cerebral hemorrhage/ or intracranial hemorrhage, hypertensive/ or stroke/ or hemorrhagic stroke/ or ischemic stroke/ or embolic stroke/ or thrombotic stroke/ or hypertension/ or peripheral vascular diseases/ | 2310942 |
|  |  |  | 6 | Hypertension/ | 817463 |
|  |  |  | 7 | Peripheral Vascular Diseases/ | 17449 |
|  |  |  | 8 | Stroke Rehabilitation/ or "National Institute of Neurological Disorders and Stroke (U.S.)"/ or Embolic Stroke/ or Ischemic Stroke/ or Stroke Volume/ or Hemorrhagic Stroke/ or Stroke/ | 403769 |
|  |  |  | 9 | Angina, Stable/ or Angina Pectoris/ or Angina, Unstable/ | 90125 |
|  |  |  | 10 | Myocardial Infarction/ | 116108 |
|  |  |  | 11 | Coronary Disease/ or Myocardial Ischemia/ or Heart Failure/ | 470165 |
|  |  |  | 12 | ("cardiovascular disease*" or "Cardiovascular incident" or "Cardiovascular accident" or "Cardiovascular outcome*" or "cardiovascular endpoint" or "cardiovascular event" or "cardiovascular death" or "'major adverse cardiovascular " or "heart attack" or "coronary heart disease" or "cerebrovascular accident" or "ischemic heart disease" or hypertension or stroke* or "peripheral artery disease*" or angina or "myocardial infarct*" or "coronary disease*" or "myocardial ischemia*" or "heart failure").tw,kf. | 2210866 |
|  |  |  | 13 | 5 or 6 or 7 or 8 or 9 or 10 or 11 or 12 | 3238249 |
|  |  |  |  | 4 and 13 | 5326 |
| PsycINFO | | | | No articles for UPF -CVD outcome in PsycINFO (Only CVD without UPF) |  |
| **Ovid Emcare** 1995 to 2024 Week 45 | | | | | 2085 |
|  |  |  | 1 | Fast Foods/ | 6284 |
|  |  |  | 2 | Food, Processed/ | 1079 |
|  |  |  | 3 | ("Ultra-processed food*" or "Ultra processed food*" or "ultraprocessed food*" or "ultra-processed product" or "fast food*" or "processed food*" or "food, processed" or "processed meat" or "artificially sweetened beverage*" or "sugary drink*" or "carbonated drink*" or "carbonated beverage*" or "food additive*" or "energy drink*" or "ready-to-eat" or chocolate or candy or cookies or "food processing" or "sugar-sweetened beverage*" or "sugar sweetened drink*" or "food additive*" or "fermented beverage*" or "frozen food*" or "meat substitute*").tw,kf. | 22530 |
|  |  |  | 4 | 1 or 2 or 3 | 25532 |
|  |  |  | 5 | cardiovascular diseases/ or heart diseases/ or atrial fibrillation/ or ventricular fibrillation/ or heart arrest/ or heart failure/ or heart failure, diastolic/ or heart failure, systolic/ or heart rupture/ or myocardial ischemia/ or angina pectoris/ or angina, unstable/ or angina, stable/ or microvascular angina/ or coronary disease/ or coronary artery disease/ or coronary occlusion/ or coronary thrombosis/ or myocardial infarction/ or anterior wall myocardial infarction/ or inferior wall myocardial infarction/ or pulmonary heart disease/ or aortic diseases/ or arterial occlusive diseases/ or cerebrovascular disorders/ or brain ischemia/ or brain infarction/ or cerebral infarction/ or infarction, anterior cerebral artery/ or infarction, middle cerebral artery/ or infarction, posterior cerebral artery/ or hypoxia-ischemia, brain/ or ischemic attack, transient/ or carotid artery diseases/ or carotid artery thrombosis/ or stroke, lacunar/ or intracranial arterial diseases/ or cerebral arterial diseases/ or intracranial aneurysm/ or "intracranial embolism and thrombosis"/ or intracranial embolism/ or intracranial thrombosis/ or sinus thrombosis, intracranial/ or intracranial hemorrhages/ or cerebral hemorrhage/ or intracranial hemorrhage, hypertensive/ or stroke/ or hemorrhagic stroke/ or ischemic stroke/ or embolic stroke/ or thrombotic stroke/ or hypertension/ or peripheral vascular diseases/ | 552809 |
|  |  |  | 6 | Hypertension/ | 197157 |
|  |  |  | 7 | Peripheral Vascular Diseases/ | 6634 |
|  |  |  | 8 | Stroke Rehabilitation/ or "National Institute of Neurological Disorders and Stroke (U.S.)"/ or Embolic Stroke/ or Ischemic Stroke/ or Stroke Volume/ or Hemorrhagic Stroke/ or Stroke/ | 105679 |
|  |  |  | 9 | Angina, Stable/ or Angina Pectoris/ or Angina, Unstable/ | 21744 |
|  |  |  | 10 | Myocardial Infarction/ | 15383 |
|  |  |  | 11 | Coronary Disease/ or Myocardial Ischemia/ or Heart Failure/ | 132910 |
|  |  |  | 12 | ("cardiovascular disease*" or "Cardiovascular incident" or "Cardiovascular accident" or "Cardiovascular outcome*" or "cardiovascular endpoint" or "cardiovascular event" or "cardiovascular death" or "'major adverse cardiovascular " or "heart attack" or "coronary heart disease" or "cerebrovascular accident" or "ischemic heart disease" or hypertension or stroke* or "peripheral artery disease*" or angina or "myocardial infarct*" or "coronary disease*" or "myocardial ischemia*" or "heart failure").tw,kf. | 481835 |
|  |  |  | 13 | 5 or 6 or 7 or 8 or 9 or 10 or 11 or 12 | 761447 |
|  |  |  | 14 | 4 and 13 | 2085 |
| CINAHL |  |  |  |  | 2000 |
|  |  |  | 1 | MH “food, processed+” OR MH “ultra-processed food+” OR MH “food, commercially packaged” OR MH “ultra processed food+” OR TI “ultra-processed food*” OR AB “ultra-processed food*” OR TI “ultra-processed diet” OR AB “ultra-processed diet” OR TI “ultraprocessed food*” OR AB “ultraprocessed food*” OR TI “sugar sweetened” OR AB “sugar sweetened” OR TI “processed food*” OR AB “processed food*” OR TI “industrial food*” OR AB “industrial food*” OR TI “junk food*” OR AB “junk food*” OR TI “ready-to-eat food*” OR AB “ready-to-eat food*” OR TI “ready-to-heat food*” OR AB “ready-to-heat food*” OR TI “western diet” OR AB “western diet” OR TI “western food*” OR AB “western food*” OR TI “processed meet” OR AB “processed meat” OR TI “artificial sweetener*” OR AB “artificial sweetener*” OR TI “carbonated beverage*” OR AB “carbonated beverage *” OR TI “fast food*” OR AB “fast food*” OR TI “sugary drink*” OR AB “sugary drink*” OR TI “soda drink*” OR AB “soda drink*” OR TI “pre-prepared food*” OR AB “pre-prepared food*” OR TI “soft drink*” OR AB “soft drink*” OR TI “breakfast cereal*” OR AB “breakfast cereal*” OR TI “packed food*” OR AB “packed food*” OR TI “packaged cookies” OR AB “packaged cookies” |  |
|  |  |  | 2 | MH"cardiovascular disease+" OR MH hypertension+ OR MH stroke+ OR MH “coronary disease+” OR MH “peripheral artery disease+” OR MH angina+ OR MH “myocardial infarction+” OR MH “heart failure+” OR MH “myocardial ischemia+” OR TI “cardiovascular disease*” OR AB “cardiovascular disease*” OR TI "Cardiovascular inciden*" OR AB "Cardiovascular inciden*" OR TI "Cardiovascular accident" OR AB "Cardiovascular accident" OR TI "Cardiovascular outcome*" OR AB "Cardiovascular outcome*" OR TI "cardiovascular endpoint*" OR TI "cardiovascular endpoint*" OR TI "cardiovascular event*" OR TI "cardiovascular event*" OR TI "cardiovascular death*" OR AB "cardiovascular death*" OR TI "major adverse cardiovascular " OR AB "major adverse cardiovascular " OR TI "heart attack*" OR AB “heart attack*” OR TI "coronary heart disease" OR AB "coronary heart disease" OR TI "cerebrovascular accident*" OR AB "cerebrovascular accident*" OR TI "ischemic heart disease" OR AB "ischemic heart disease" OR TI hypertension OR AB hypertension OR TI stroke* OR AB stroke* OR TI "peripheral artery disease*" OR AB "peripheral vascular disease*" OR TI angina OR AB angina OR TI "myocardial infarct*" OR AB "myocardial infarct*" OR TI "coronary disease*" OR AB “coronary disease” OR TI "myocardial ischemia*" OR AB "myocardial ischemia*" OR TI "heart failure" OR AB “heart failure” |  |
|  |  |  | 3 | 1 AND 2 | 2000 |
| ProQuest Central | | | | | 444 |
|  |  |  | 1 | MAINSUBJECT.EXACT(“Ultra-processed foods” OR “Ultraprocessed foods” OR “Processed foods” OR “Ultra processed foods”) OR TI, AB(“Ultra-processed food*” OR “ultra processed food*” OR ("processed food") OR ("industrial food") OR ("junk food" OR "junk foods") OR “ready to eat food*” OR “ready to heat food*” OR ("western diet" OR "western diets") OR ("western food" OR "western foods") OR “processed meat” OR ("artificial sweetener" OR "artificial sweeteners") OR ("carbonated drink" OR "carbonated drinks") OR ("sugary drinks") OR ("soda drink") OR ("soft drink" OR "soft drinks") OR ("breakfast cereal" OR "breakfast cereals") OR ("packaged food" OR "packaged foods") OR “packaged cook*” OR "food, commercially packaged” OR ("carbonated beverage" OR "carbonated beverages") OR “sugar sweetened”) |  |
|  |  |  | 2 | MAINSUBJECT.EXACT("cardiovascular disease*" OR hypertension OR stroke OR “coronary disease*” OR “peripheral artery disease*” OR angina* OR “myocardial infarction*” OR “heart failure” OR “myocardial ischemia” ) OR TI,AB ( “cardiovascular disease*” OR "Cardiovascular inciden*" OR "Cardiovascular accident" OR "Cardiovascular outcome*" OR "cardiovascular endpoint*" OR "cardiovascular event*" OR "cardiovascular death*" OR "major adverse cardiovascular " OR "heart attack*" OR "coronary heart disease" OR "cerebrovascular accident*" OR "ischemic heart disease" OR hypertension OR stroke* OR "peripheral vascular disease*" OR angina OR "myocardial infarct*" OR "coronary disease*" OR "myocardial ischemia*" OR "heart failure" ) |  |
|  |  |  | 3 | 1 AND 2 | 444 |
| Scopus |  |  |  |  | 5615 |
|  |  |  | 1 | TITLE-ABS-KEY( “Food, processed*” OR “ultra-processed food*” OR “processed food” OR “processed diet*” OR “ultraprocessed food*” OR “industrial food*” OR “junk food*” OR “ready to eat food*” OR “ready to heat food*” OR “western diet*” OR “western food*” OR “processed meat” OR “artificial sweet*” OR “carbonated drink*” OR “sugary drink*” OR “soda drink*” OR “soft drink*” OR “sugar sweetened” OR “breakfast cereal*” OR “packaged food*” OR “packaged cook*” OR “food, commercially packaged” OR “carbonated beverage*” OR “sugar sweetened”) |  |
|  |  |  | 2 | TITLE-ABS-KEY ("cardiovascular disease*" OR hypertension OR stroke OR “coronary disease*” OR “peripheral artery disease*” OR angina* OR “myocardial infarction*” OR “heart failure” OR “myocardial ischemia” OR "Cardiovascular inciden*" OR "Cardiovascular accident" OR "Cardiovascular outcome*" OR "cardiovascular endpoint*" OR "cardiovascular event*" OR "cardiovascular death*" OR "major adverse cardiovascular " OR "heart attack*" OR "coronary heart disease" OR "cerebrovascular accident*" OR "ischemic heart disease" OR hypertension OR stroke* OR "peripheral artery disease*" OR angina OR "myocardial infarct*" OR "coronary disease*" OR "myocardial ischemia*" OR "heart failure" ) |  |
|  |  |  | 3 | 1 AND 2 | 5615 |
| Web of Science | | | | | 4728 |
|  |  |  | 1 | (“Food, processed” OR “Ultra-processed food*” OR “ultraprocessed food*” OR “processed food” OR “processed diet*” OR “industrial food*” OR “junk food*” OR “ready to eat food*” OR “ready to heat food*” OR “western diet*” OR “western food*” OR “processed meat” OR “artificial sweet*” OR “carbonated drink*” OR “sugary drink*” OR “soda drink*” OR “soft drink*” OR “breakfast cereal*” OR “packaged food*” OR “packaged cook*” OR “food, commercially packaged” OR “carbonated beverage*” OR “sugar sweetened”) |  |
|  |  |  | 2 | ("cardiovascular disease*" OR hypertension OR stroke OR “coronary disease*” OR “peripheral artery disease*” OR angina* OR “myocardial infarction*” OR “heart failure” OR “myocardial ischemia” OR "Cardiovascular inciden*" OR "Cardiovascular accident" OR "Cardiovascular outcome*" OR "cardiovascular endpoint*" OR "cardiovascular event*" OR "cardiovascular death*" OR "major adverse cardiovascular " OR "heart attack*" OR "coronary heart disease" OR "cerebrovascular accident*" OR "ischemic heart disease" OR hypertension OR stroke* OR "peripheral artery disease*" OR angina OR "myocardial infarct*" OR "coronary disease*" OR "myocardial ischemia*" OR "heart failure" ) |  |
|  |  |  | 3 | 1 AND 2 | 4728 |
| Cochrane library | | | | | 249 |
|  |  |  | 1 | [mh “food, processed”] OR [mh “Ultra-processed foods”[mj]] OR [mh ^“Ultraprocessed food”] OR [mh ^“Ultra processed foods”] OR  “Ultra processed food”:ti,ab,kw OR (processed NEXT food):ti,ab,kw OR “industrial food”:ti,ab,kw OR “junk food”:ti,ab,kw OR “ready to eat food”:ti,ab,kw OR “ready to heat food”:ti,ab,kw OR (western NEXT diet*):ti,ab,kw OR “western food”:ti,ab,kw OR “processed meat”:ti,ab,kw OR (artificial NEXT sweet*):ti,ab,kw OR “carbonated drink”:ti,ab,kw OR (sugary NEXT drink*):ti,ab,kw OR “soda drink”:ti,ab,kw OR “soft drink”:ti,ab,kw OR “breakfast cereal”:ti,ab,kw OR “packaged food”:ti,ab,kw OR (packaged NEXT cook*):ti,ab,kw |  |
|  |  |  | 2 | [mh "cardiovascular disease"] OR [mh ^hypertension] OR [mh ^stroke] OR [mh “coronary disease”] OR [mh “peripheral artery disease”] OR [mh ^angina] OR [mh “myocardial infarction”] OR [mh “heart failure” ] OR [mh “myocardial ischemia”] OR "cardiovascular disease":ti,ab,kw OR (Cardiovascular NEXT inciden*):ti,ab,kw OR "Cardiovascular accident":ti,ab,kw OR (Cardiovascular NEXT outcome*):ti,ab,kw OR (cardiovascular NEXT endpoint*):ti,ab,kw OR (cardiovascular NEXT event*):ti,ab,kw OR (cardiovascular NEXT death*):ti,ab,kw OR "major adverse cardiovascular ":ti,ab,kw OR (heart NEXT attack*):ti,ab,kw OR "coronary heart disease":ti,ab,kw OR (cerebrovascular NEXT accident*):ti,ab,kw OR "ischemic heart disease":ti,ab,kw OR hypertension:ti,ab,kw OR stroke*:ti,ab,kw OR "peripheral artery disease":ti,ab,kw OR angina:ti,ab,kw OR (myocardial NEXT infarct*):ti,ab,kw OR "coronary disease":ti,ab,kw OR (myocardial NEXT ischemia*):ti,ab,kw OR "heart failure":ti,ab,kw |  |
|  |  |  | 3 | #1 AND #2 | 249 |
| Google scholar | | | | | 123 |
| Research Rabbit | | | | | 28 |
| Clinical trial org | | | | | 16 |

**Supplementary Table 3.** Quality of evidence for the link between ultra-processed food consumption and cardiovascular disease based on GRADE criteria

| **Outcome** | **Number of studies** | **Pooled effect (HR) [95% CI]** | **I^2^(%)** | **Risk of bias** | **Inconsistency** | **Indirectness** | **Imprecision** | **Publication bias** | **Overall GRADE certainty** | **Importance** |
| --- | --- | --- | --- | --- | --- | --- | --- | --- | --- | --- |
| Overall CVD^a^ | 21 | 1.09 (1.05, 1.12) | 44.2 | High | Not serious | Not serious | Not serious | Not serious | Very low | Important |
| Coronary Heart Disease^b^ | 9 | 1.19 (1.05, 1.34) | 88.2 | High | Serious | Not serious | Not serious | Not serious | Very low | Important |
| Cerebrovascular disease^c^ | 8 | 1.08 (1.01, 1.16) | 32.1 | High | Not serious | Not serious | Not serious | Not serious | Very low | Important |
| Hypertension^d^ | 10 | 1.16 (1.08, 1.26) | 69.8 | High | Serious | Not serious | Not serious | Serious | Very low | Important |

^a^ Downgraded due to high risk of bias.

^b^ Downgraded due to high risk of bias and serious inconsistency (high heterogeneity).

^c^ Downgraded due to high risk of bias.

^d^ Downgraded due to high risk of bias, serious inconsistency (high heterogeneity) and serious publication bias.

**Supplementary Table 4.** Summary of food items and food subgroups categorised as ultra-processed food by authors and countries for prospective cohort studies examined ultra-processed food intake and cardiovascular diseases outcomes among adults

| Author-Year | **Country** | **Number and Professional background of authors classified NOVA** | **Food subgroups of UPF** | **Food items classified as UPF** | **Funding sources** |
| --- | --- | --- | --- | --- | --- |
| Cordova- 2023 | 7 EU countries (Denmark, Germany, Italy, Spain, Sweden, the Netherlands, and the United Kingdom) | NR | - Ultra-processed breads and cereals - Sauces, spreads, and condiments - Sweets and desserts - Savory snacks - Dairy substitute products, Meat alternatives - Animal-based products: Processed meat - Ready-to-eat/heat mixed dishes - Artificially and sugar-sweetened - Other ultra-processed food | Breads, Biscuits, Breakfast cereals;  Sauces, dressing and gravies - also in powder/dehydrated form/condensed form, Vegetable spread and products, Margarine;  Pastries, buns, and cakes, Ice cream, ice pops and frozen yogurts, Sweet snacks, Industrial desserts, Dairy desserts and drinks (ultra processed versions);  Packaged salty snacks; Dairy substitute products, Meat alternatives; Processed meat (beef, pork, and fish), Processed cheese;  Potato products, Pizza and focaccia (dough),Pasta (filled), Instant and canned soups, Ready meals, Vegetables and legumes in ultra-processed medium, Rice-based dishes; Soft drinks, Fruit drinks, iced tea and other sweetened beverages;  Artificial sweeteners  , Alcohol-free versions of alcoholic beverages, Nutrition powders and drinks | The Fondation de France, Cancer Research UK and the World Cancer Research Fund International |
| Zhong -2021 | USA | 2 Dietitians | - Soft Drinks - Cereals - Ultra-processed fruits and vegetables - Ultra-processed dairy products - Meat and fish - Sauces and dressings - Salty snacks - Sugary products - Margarine | From 156 questionnaire items, 65 food items of UPF:  Soft drinks, diet/caffeinated  Soft drinks, regular/caffeinated  Soft drinks, diet/decaffeinated  Soft drinks, regular/decaffeinated  Hot dogs, turkey/low fat  Hot dogs, regular  English muffins and bagels  White bread/rolls  Corn bread/muffins  Biscuits  Quick breads  Donuts, sweet rolls, Danishes, and pop tarts  Pancakes, waffles, and French toast  Ready-to-eat cereal, highly fortified  Ready-to-eat cereal, other  Cookies and brownies  Cakes, low fat  Cakes, regular  Pies, cream/custard/other  Pies, fruit  Pies, pecan  Pies, pumpkin/sweet potato  Lasagna, ravioli, shells  Macaroni and cheese  Pizza without meat  Fried potatoes  Potato salad  Fruit drinks, diet  Fruit drinks, regular  Sour cream, low fat  Sour cream, regular  Cream cheese, low fat  Cream cheese, regular  Ice cream/ice milk, low fat  Regular ice cream  Frozen yogurt, ices, and sorbet  Creamed soups  Restaurant or industrial hamburgers  Fried chicken, dark meat/no skin  Fried chicken, dark meat/with skin  Fried chicken, white meat/no skin  Fried chicken, white meat/with skin  Fried fish, fat added  Pizza with meat  Gravy  Salad dressing, low fat on salad and vegetables  Salad dressing, non-fat on salad and vegetables  Salad dressing, regular on salad and vegetables  Mayonnaise, diet on salad  Mayonnaise, fat free on salad  Mayonnaise, regular on salad  Mayonnaise, diet on sandwich  Mayonnaise, fat free on sandwich  Mayonnaise, regular on sandwich  Crackers  Potato/corn/other chips  Potato/corn/other chips-low fat  Popcorn  Candy, chocolate  Candy, not chocolate  Saccharine in coffee and tea  Margarine, diet  Margarine, fat free  Margarine, regular  Margarine, diet on bread) | no funding specified |
| Chen- 2022 | UK | NR | - Drinks - Cereals - Sweets - Snacks | From 204 common food and drink items, 57 items of UPF:  Low calorie drink, fizzy drink, squash, orange juice, grapefruit juice, instant coffee, other coffeea, flavored milk, low calorie hot chocolate, hot chocolate, spirits, other alcohola;  Porridgea, oat crunch, sweetened cereal, other cereala, sliced breada, bapa, crispbread, snackpot;  Yogurta, ice-cream, milk-based puddinga, other milk-based pudding, soya dessert, fruitcake, cake, doughnut, sponge pudding, cheesecake, other desserta, chocolate bar, white chocolate, milk chocolate, dark chocolate, chocolate-covered raisin, chocolate sweet, diet sweet, sweets, chocolate-covered biscuits, chocolate biscuits, sweet biscuits, cereal bar, other sweetsa;  Crisp, savoury biscuits, cheesy biscuits, other savoury snack, powdered/instant soup, canned soup, sausage, crumbed or deep-fried poultry, buttera, margarinea. | National Natural Science  Foundation of China |
| Monge -2021 | Mexico | Five trained dietitians | - Dairy - Cereal and breakfast cereals - Added Fats - Sugary products - Salty snacks - Processed meats - Beverages (sugar-sweetened) - Beverages (distilled alcohol) - Fast food | From 140-items of FFQ, 44 food items of UPF:  flavored yoghurt, fermented dairy (yakult), petite-suisse (danonino), ice-cream;  oats, ready-to-eat (RTE) cereal (high and low fiber), cereal bar, RTE loaf of bread (high and low fiber), wheat tortilla;  cream, cream cheese, margarine; jello or flan, cake, cookies, candies, chocolate, jelly/ate/honey, pan dulce, cakes and donuts;  chips and saltines;  sausages, pork and turkey ham, bacon, chorizo/longaniza, other deli meats;  regular and diet cola and soda, flavored water (artificial)*, orange juice, soy milk;  brandy, whisky, tequila, mezcal, rum, aguardiente;  hamburgers, hot dogs, sandwich (processed meat torta), pizza | Supported by the American Institute for Cancer Research and the Consejo Nacional de Ciencia y Tecnología. |
| Du -2021 | USA | NR | - Dairy products - fats and oils - meats - sugary products - bakery goods - cereals - fried foods - beverages - liquor | From 66-item FFQ, 35 items of UPF  Ice cream; margarine and spreads; hamburgers, processed meats (sausage, salami, bologna), meats prepared with added sauce; catsup, hot sauce and soy or steak sauces; hot dogs, chocolate bars or pieces (Hershey’s, Plain M&M’s, Snickers, Reese’s), candy without chocolate, ready-made pie, donut, biscuits or cornbread; Danish pastry, sweet roll, coffee cake and croissant; cookies, cold breakfast cereal, potato chips or corn chips, French fried potatoes, food fried away from home, low calorie and regular soft drinks, orange and grapefruit juice, fruit-flavored punch or non-carbonated beverages (lemonade, Kool-Aid, Hawaiian Punch), hard liquor. | supported by National Heart, Lung, and Blood Institute |
| Kityo-2023 | Korea | Two (  A nutritionist independently assigned each of the 106 items to a NOVA group and then consulted a registered dietitian who also participated in the design of the HEXA SQFFQ). | - Instant noodles (ramen) - Cornflakes - Loaf bread (‘sikppang’); bread with red bean; other bread - Jam/honey/butter/margarine (when eaten on bread); - Cake/chocopie cookie/cracker/snack; candy/chocolate - Pizza/hamburger - Processed meat (ham, sausage) - Fish cake/Crab meat - Milk - yogurt/Yoplait - Ice-cream - Soymilk drink - Soft drinks (cola/cider)/ fruit beverages - Other drinks | From 106 items of FFQ, food items of UPF:  Instant noodles;  cereal, rice crisps; cereal cornflakes; cereal, brown rice flakes;  Bread, red bean bread, buns, red bean filling, gombo bread; doughnut; cream bread; castella;  strawberry jam*, apple jam*, apricot jam*, grape jam*, butter*, margarine*;  Choco Pie, Roll Cake, Whipped Cream, Blueberry Cake, Sponge Cake, Pound Cake, Hot Cake/Pancake, Montshell /Fresh cream; Dry Bread, Biscuit, Soft, Sen Bay, Rice Cake, Potato chips/crisps, Shrimp Snack, Corn snacks, Crackers; Drops, Candy;  Pizza, hamburger;  Raw bacon, raw Sausage, dry Sausage, Frankfurt Sausage, Hot Dog, Ham, Slice, Luncheon Meat;  Imitation crab*, crab sausage*;  Coffee milk*, chocolate milk*, fruit flavored milk*;  Liquid yoghurt, strawberry-flavored yoghurt;  Ice cream, 12% fat; ice cream, 8% fat; ice cream, strawberry-flavored; ice cream, vanilla flavored; ice cream, chocolate-flavored; sherbet; selection; world corn; Joanna;  Soymilk drink (vegemil);  fruit soda; lemon soda; orange soda; cream soda; grape soda; cider; coke;  Sweet rice punch (Sikhye) * |  |
| Srour-2019 | France | Three trained dieticians categorised the food and beverage items  into one of the four food groups in NOVA and five researchers—then reviewed the classification. | - Beverages - Sugary products - Fruits and vegetables - Starchy foods and breakfast cereals - Meat, fish and eggs - Dairy products - Fats and sauces - Salty snacks | Carbonated drinks; sweet or savoury packaged snacks; ice-cream, chocolate, candies (confectionery); mass-produced packaged breads and buns; margarines and spreads; industrial cookies (biscuits), pastries, cakes, and cake mixes; breakfast ‘cereals’, ‘cereal’ and ‘energy’ bars; ‘energy’ drinks; flavoured milk drinks; cocoa drinks; sweet desserts made from fruit with added sugars, artificial flavours and texturizing agents; cooked seasoned vegetables with ready-made sauces; meat and chicken extracts and ‘instant’ sauces; ‘health’ and ‘slimming’ products such as powdered or ‘fortified’ meal and dish substitutes; ready to heat products including pre-prepared pies, pasta and pizza dishes; poultry and fish ‘nuggets’ and ‘sticks’, sausages, burgers, hot dogs, and other reconstituted meat products, and powdered and packaged ‘instant’ soups, noodles and desserts | Supported by the Ministère de la Santé, Santé Publique France, Institut National de la Santé et de la Recherche Médicale, Institut National de la Recherche Agronomique, Conservatoire National des Arts et Métiers, and Université Paris |
| Li 2023 | UK | NR | Food groups of UPF are not reported | From 168-item FFQ, food items of UPF:  Soft drinks, sweets/candies, chocolate, snacks, cookies, cakes, ice cream, margarine, mayonnaise,  wafers, wheat crusts, low-fiber bread, marmalade, jam and honey, gruel, breakfast cereals,  tomato ketchup, nutritional powder, industrial soups, fish sticks, sausage, bacon and ham, blood  pudding | funded by Crafoord Foundation and Albert Påhlsson Foundation |
| Bonaccio-2023 | Italy | 2 researchers (background unspecified) | - Processed meat - Cakes, pies, pastries, puddings (non-milk based) - Fruit drinks - Pizza (not homemade) - Ice-cream - Crispbread/rusks - Fruit yoghurts - Carbonated/soft/isotonic drinks, diluted syrups - Dry cakes, biscuits - Savoury snacks - Stock cube - Chocolate - Breakfast cereals - Spirits, brandy - Spreadable cheese - Nut spread - Confectionery non chocolate - Sliced cheese - Margarine - Salty biscuits, aperitif biscuits, crackers - Artificial sweeteners - Mayonnaise and similar | From 188 food items, 22 food groups categorised as UPF:  Processed meat;  Cakes, pies, pastries, puddings(non-milk based);  Fruit drinks;  Pizza (not homemade);  Ice-cream;  Crispbread/rusks;  Fruit yoghurts;  Carbonated/soft/isotonic, drinks, diluted syrups;  Dry, cakes, biscuits;  Savoury snacks;  Stock cube; Chocolate;  Breakfast cereals;  Spirits, brandy;  Spreadable cheese;  Nut spread;  Confectionery non chocolate;  Sliced cheese;  Margarine;  Salty biscuits, aperitif biscuits, crackers; Artificial sweeteners;  Mayonnaise and similar | supported by research grants from Pfizer Foundation (Rome, Italy), the Italian Ministry of University and Research (MIUR, Rome, Italy)–Programma Triennale di Ricerca, Decreto no. 1588 and Instrumentation Laboratory, Milan, Italy |
| Wang- 2024 | UK | NR | - Ultra-processed breads - Soft drinks - Milk-based drinks - Pastries, buns, and cakes - Sausages or other reconstituted meat products - Industrial-manufactured desserts - Biscuits,cereal bar,other - Other beverages or coffee drinks - Butter/Margarine - Packaged salty snacks - Alcoholic drinks - Other ultra-processed foods | Bap intake, Bread roll intake,Naan bread intake, Crispbread intake, Oatcakes intake, Other bread intake;  Low calorie drink intake, Fizzy drink intake carbonated (fizzy) drinks,  Squash intake; Flavoured milk intake,  Milk-based pudding intake, Other milk-based pudding intake;  Double crust pastry intake, Danish pastry intake,Scone intake, Ice-cream intake, Soya dessert intake, Fruitcake intake, Cake intake, Doughnut intake, Sponge pudding intake, Cheesecake intake, Other dessert intake, Single crust pastry intake, Crumble intake, Pancake intake, Scotch pancake intake, Yorkshire pudding intake, Indian snacks intake, Croissant intake; Sausage intake,Crumbed or deep-fried poultry intake, breadcrumbs or deep fried  Bacon intake,Ham intake, Vegetarian sausages/burgers intake,Quorn intake,  Other vegetarian alternative intake;  Chocolate bar intake, White chocolate intake,Milk chocolate intake, Dark chocolate intake, Chocolate-covered raisin intake,Chocolate sweet intake, Diet sweets intake, Sweets intake;  Chocolate biscuits intake,Sweet biscuits intake,Savoury biscuits intake, Cheesy biscuits intake,Cereal bar intake,Other sweets intake sweet snacks,Chocolate-covered biscuits intake; Instant coffee intake, Intake of artificial sweetener added to coffee, Intake of artificial sweetener added to tea; bread slices with butter/margarine, baguettes with butter/margarine, baps with butter/margarine, bread rolls with butter/margarine, crackers/crispbreads with butter/margarine, oatcakes with butter/margarine, other bread types with butter/margarine; Crisp intake, Other savoury snack intake; Fortified wine intake, Spirits intake; and  Pizza intake, Couscous intake, Baked bean intake, Fried potatoes intake,  Powdered/instant soup intake,  Snackpot intake (snack pot, noodles/rice),Low fat cheese spread intake, and Cheese spread intake | Not mentioned. |
| Sandra González‐Palacios-2023 | Spain | Experts on nutritional epidemiology and dietitians classified (number not specified) | - *Dairy products* - *Processed meats* - *Sweets* - *Fast-foods* - *Beverages* - *Alcoholic beverages* | From 143 food items, 36 foods of UPF:  Flavored milk drinks; Petit-suise; Cheese portions or cream cheese; Custard, pudding or similar; Ice cream; Ham; Processed meats such as dried sausage, chorizo or similar; Patés and foie gras; Burgers and, meatballs; Crisps; Breakfast cereals; Pizza; Margarine; Plain biscuits; Whole grain biscuits; Chocolate biscuits; Croissants, pastries or similar; Doughnuts; Muffins; Spanish churros or similar; Pies; Chocolates and chocolate; Cocoa powder; Spanish nougat; Marzipan or similar; Croquetas, fishcakes, pasties, or similar; Packet or canned soups; Mustard; Commercial mayonnaise; Fried tomato sauce or ketchup; Snacks other than crisps; sugar-sweetened soft drinks; artificially- sweetened soft drinks; commercial fruit juices; Liquors; Spirits: whisky, vodka, gin and cognac. | supported by the official Spanish Institutions for funding scientific biomedical research, CIBER Fisiopatología de la Obesidad y Nutrición and Instituto de Salud Carlos III, through the Fondo de Investigación para la Salud, which is co-funded by the European Regional Development Fund |
| Bonaccio-2022 | Italy | NR | NR | From 188 foods, a total of 22 foods and beverages grouped as UPF:  Processed meat, Cakes, pies, pastries, puddings, (non-milk based), Fruit drinks, Pizza (not homemade), ice-cream,Crispbread/rusks,Fruit yoghurts,Carbonated/soft/isotonic drinks, diluted syrups, Dry cakes, biscuits, Snacks,Stock cube,Chocolate , Breakfast cereals,Spirits, brandy, Spreadable cheese, Nut spread, Confectionery non chocolate,Sliced cheese, Margarine,Salty biscuits, aperitif biscuits, crackers, Mayonnaise and similar, Artificial sweeteners | supported by research grants from Pfizer Foundation (Rome, Italy), the Italian Ministry of University and Research (MIUR, Rome, Italy) and Instrumentation Laboratory, Milan, Italy. |
| Fang -2024 | USA | Three senior nutrition epidemiologists | - ultra-processed breads and breakfast foods; - fats, condiments, and sauces; - packaged sweet snacks and desserts; - sugar sweetened and artificially sweetened beverages; - ready-to-eat/heat mixed dishes; - meat/poultry/seafood based ready-to-eat products (for example, processed meat) - packaged savory snacks; dairy based desserts; and - other. | Whole grain (cold breakfast cereal; rye, pumpernickel bread; whole grain bread);  English muffins, bagels, rolls; White bread; Cream cheese, Ketchup, Margarine, Mayonnaise (regular and low fat), Non-dairy coffee  whitener, Red chili sauce, Salad dressings, Salsa, Soy sauce, Spread butter; Apple sauce; Candy bar with chocolate; Candy bar without chocolate; Chocolate bars; Dark  chocolate bars; Breakfast bar; Energy bar; High protein, low carb candy bar; Brownies;  Cookies; Fat-free/reduced fat doughnuts; Muffins or biscuits; Ready-made cake; Readymade sweet rolls and coffee cakes; Ready-made pie; Jams, jellies, preserves, hone; Fat-free popcorn; Fat-free, light crackers; Regular crackers; Regular carbonated beverage with caffeine & sugar; Carbonated, low-cal ‘diet’ beverage  with caffeine; Carbonated beverage without caffeine but with sugar; Fruit drinks/pouch,  lemonade, Sunny D, Koolaid, sugared ice tea or other non-carbonated fruit drink – NOT juice; Bacon; Beef hotdogs or pork hotdogs; Breaded fish cakes, pieces, sticks; Chicken hotdogs  or turkey hotdogs; Processed meats, sausages; Salami, bologna, processed meat  sandwiches; Chowder or cream soup, French fries potatoes, Pizza, Ready-made soup from cans, Soup  made with bouillon; Artificially sweetened yogurt; Flavored yogurt without Nutrasweet; Frozen yogurt, ice cream,  sherbet; Nutrasweet or equal, Other artificial sweeteners, Splenda, Distilled alcohol | supported by the US National Institutes of Health grants |
| Bonaccio- 2021 | Italy | NR | From 188 food items, 18 food groups of UPF:   - Processed meat, - Cakes, pies, pastries, puddings, (non-milk based), - Fruit drinks, - Pizza (not homemade), - ice-cream, - Crispbread/rusks, - Fruit yoghurts, - Carbonated/soft/isotonic drinks, diluted syrups, - Dry cakes, biscuits, - Snacks, - Stock cube, - Chocolate , - Breakfast cereals, - Spirits, brandy, - Spreadable cheese, - Nut spread, - Confectionery non chocolate, - Sliced cheese, - Margarine, - Salty biscuits, aperitif biscuits, crackers, Mayonnaise and similar, Artificial sweeteners | NR | Supported in part by Italian Ministry of Health grant |
| Torres-Collado-2024 | Valencia, Spain | Three  dietitians  classified. | - *Dairy* - *Eggs, meats, fish, seafood* - *Bread, grains or others* - *Fat* - *Sweets and pastries* - *Drinks* - *Pre-cooked, convenient-food, others* | From 93 food items, 39 foods as UPF:  Dairy Condensed milk; cheese portions or cream cheese; custard, pudding or similars; ice cream; Eggs, meats, fish, seafood Cured ham and processed meats (sausage, salami, bologna); sausages or similar; patés, foie-gras; hamburgers Bread, grains or others Pretzel; fries; potato chips. Fat Margarine Sweets and pastries Biscuits; biscuits with chocolate; croissants, donuts; cupcake, sponge cake; pies, cakes; Spanish churros; chocolates, praline; chocolate powder or similar Drinks Brandy, gin, rum, whisky, vodka, spirits; soft drinks with gas (cola, orange soda, fanta or similar); commercial fruit juices Pre-cooked, convenient-food, others Croquetas; commercial mayonnaise; packed or canned soups; commercial tomato sauce; fish “sticks” and breadsticks | supported by a grant from the Direccion General de Salud Pública, Generalitat Valenciana 1994 and the Fondo Investigacion Sanitaria. |
| Kermani-Alghoraishi - 2024 | Iran | NR | NR | From 48 food items, 9 foods categorised as UPF:  margarine, sausage, artificial soft drinks, sugar-sweetened beverages, cookies, biscuits, confectionaries, chocolate, canned foods, and mayonnaise sauce | The Isfahan Cardiovascular Research Center, Cardiovascular Research Institute, affiliated with the Isfahan University of Medical Sciences, funded |
| Sullivan-2023 | USA | Two researchers independently categorized all items, with substantial agreement (Cohen’s κ=0.73). Discordantly classified items were conservatively assigned to the less processed group. | - Beverages - Snacks and sweets - Grains - Fats and oils - Protein foods - Mixed dishes - Vegetables - Condiments and sauces - Alcoholic beverages - Sugars | fruit drinks, meal-replacement beverages, soft drinks; crackers, potato chips, corn chips, pretzels, energy bars, frozen yogurt, ice cream, cake, cookies, brownies, doughnuts, sweet rolls, Danish, fruit crisp/cobbler, pies, chocolate candy, other candy; ready-to-eat breakfast cereals, bagels, English muffins, bread, rolls, cornbread, biscuits, sweet muffins, dessert breads; salad dressing, margarine, cream cheese, mayonnaise, nondairy creamer; roast beef, poultry cold cuts, deli-style ham, other cold cuts, hot dogs, bacon, sausage, fish sticks, fried fish, tofu∗ and soy meat products, egg substitute; stuffing, dumplings, chili, Mexican foods, pizza; french fries, home fries, hash-browned potatoes, tater tots; cheese sauce, ketchup, gravy; liquor, mixed drinks; jams, jellies, honey. | no relevant financial interests. |
| Passinho-2023 | Spain | NR | - dairy products, - processed meats, - sweets, - fast-foods, - beverages and - alcoholic beverages | From 143 foods, 36 items classified in the UPF:  Flavored milk drinks; Petit-suise; Cheese portions or cream cheese; Custard, pudding or similar; Ice cream; Ham; Processed meats such as dried sausage, chorizo or similar; Patés and foie gras; Burgers and, meatballs; Crisps; Breakfast cereals; Pizza; Margarine; Plain biscuits; Whole grain biscuits; Chocolate biscuits; Croissants, pastries or similar; Doughnuts; Muffins; Spanish churros or similar; Pies; Chocolates and chocolate; Cocoa powder; Spanish nougat; Marzipan or similar; Croquetas, fishcakes, pasties, or similar; Packet or canned soups; Mustard; Commercial mayonnaise; Fried tomato sauce or ketchup; Snacks other than crisps; sugar-sweetened soft drinks; artificially- sweetened soft drinks; commercial fruit juices; Liquors; Spirits: whisky, vodka, gin and cognac. | supported by the official Spanish Institutions for funding scientific biomedical research, CIBER Fisiopatología de la Obesidad y Nutrición (CIBEROBN) and Instituto de Salud Carlos III |
| Orlich-2022 | North America (US and Canada) | NR | - Fruits - Vegetables - Legumes - Nuts & seeds - Grains & cooked cereals; breads - Store-bought cold cereals - Oils and fats - Snack foods - Sweets & desserts - Meal replacements - Meat analogues - Dairy products - Meats - Beverages - Mixed foods | fruit jams, preserves, fruit pie fillings, French fries, hash browns, fried potatoes, catsup, chili with beans, nut candies, white breads,  other breads (bagels, biscuits, corn bread—write in), gluten steaks, Special K (Kelloggs), Frosted Flakes (Kelloggs), Frosted Mini Wheats (Kellogs), Honey Bunches of Oats (Post), Cinnamon Toast Crunch (General Mills),100% Natural Oats, or Oats & Honey (Quaker), salad dressings, mayo or miracle whip  gravies, margarines, potato chips, doughnuts, cookies (store-bought),  cake, ice cream, milk shakes, ice milk, frozen yogurt, other sweets, meal replacement drinks, meat analogues, cheese (American processed and cheddar), cream cheese/spreads,  other (whipping cream/sour cream),  evaporated/condensed milk,  soy/imitation cheese,  soy/rice drinks, processed red meat  processed white meat, soft drinks/soda, other (hot chocolate),  spirits/liqueurs, macaroni and cheese pizza | the Ardmore Institute of Health and Loma Linda University Health |
| Oladele-2024 | USA | NR | NR | bacon, bagel , baked beans, bars power, bean refried, biscuit , bread dark, bread white, Cake low fat, Cake regular, Candy not chocolate, Cereal bran, Cereal fiber, Cereal high fiber, Cereal not fiber, Cereal raisin bran, Cereal total, Cheese low fat, Chili beans, Chinese dish, chocolate, coleslaw, Cookie low fat, Cookie regular, cornbread, cracker ,Creamer nondairy, Crisco, French fry, gravy, Half margarine butter, ham hamburger, hotdog, Hot dog low fat, ice cream, Ice cream low fat, jelly, Kool-Aid, Lunch meat low fat, Mac n cheese, Margarine diet , Margarine soft , Margarine stick, mayo, Meat, Milk rice substitute, Milk soy, pancake, pastry, Pie cobbler, Pie pumpkin, pizza, Salad dressing, Salad dressing diet, Sauce mustard BBQ, Sausage breakfast, Shakes diet ,Snack salty , Snack salty, low fat, Soft drinks not diet, soup lentil , Soup other, Soup vegetable, Sunny-d, taco, tortilla | Supported by the National Heart, Lung, and Blood Institute Career Development Award and Programs to Increase Diversity Among Individuals Engaged in Cardiovascular Health-Related Research. |
| Pant-2024 | Australia | 2 authors (unspecified background) | - Milk-based drinks - Mass-produced packaged breads - Margarine and other spreads - Breakfast cereals - Packaged salty snacks - Packaged ready meals - Confectionary - Milk-based drinks - Ice cream, ice pops and frozen yogurts - Reconstituted meat products - Industrial potato chips - Sauces, dressing and gravies | Soya milk, Bread White bread Hi-fibre, Bread White bread, Bread Wholemeal bread, Bread rye bread, Multigrain bread, Margarine, Polyunsaturated margarine, Monounsaturated margarine, Butter and margarine blend, All Bran, Sultana Bran™, FibrePlus™, Branflakes™, Weet Bix™, Vita Brits™, Weeties™, Cornflakes, Nutrigrain™, Special K™, Crackers, crispbreads, dry biscuits, Sweet biscuits, Cakes, sweet pies, tarts and other sweet pastries, Meat pies, pastries, quiche and other savoury pastries, Hamburger with a bun, Chocolate, Flavoured milk drink (coca, Milo (TM) etc), Corn chips, potato crisps, Twisties(TM) etc, Vegemite(TM), Marmite(TM) or Promite™, Ice-cream, Yoghurt, Bacon, Ham , Corned beef, luncheon meats or salami, Sausages or frankfurters, Fruit juice, Potatoes roasted or fried (include hot chips), Tomato sauce, tomato paste, Baked beans | supported by a Heart Foundation Fellowship |
| Moreira-2022 | Brazil | NR | NR | Sweet and savoury biscuits; ice cream, candies, chocolate and treats generally; breakfast cereals and cereal bars; cakes and cake mixes; instant soups, noodles and seasonings; ready-made sauces; margarine; packet snacks; non-carbonated sweetened beverages (soft drinks) and carbonated sweetened beverages (soft drinks); yogurts and other dairy drinks with added colouring and/or flavouring; frozen and ready-to-heat products such as pasta dishes, pizzas, hamburgers and breaded chicken or fish meat extracts such as nuggets, sausages and other sausages; loaves of  bread, buns for hamburgers or hot dogs. | National Council for Scientific and Technological Development |
| Hang- 2024 | USA | NR | - ultra-processed bread and breakfast foods - fats/condiments/sauces - packaged sweet snacks/desserts - sugar or artificially sweetened beverages - animal protein-based ready-to-eat foods - flavoured yogurt/dairy-based desserts - packaged savory snacks - ready-to-eat/ready-to-heat mixed dishes - others | 130 food items, UPF food items not mentioned. | supported by grant MRSG-17-220-01-NEC from the American Cancer Society |
| Tu- 2023 | UK | NR | NR | Items considered ultra-processed: low calorie drink, fizzy drink, squash drink, orange drink,  grapefruit juice, pure fruit and vegetable juice, fruit smoothie, dairy smoothie, flavoured  milk, oat crunch, sweetened cereal, plain cereal, bran cereal, wholewheat cereal, other cereal,  cheese spread, low fat cheese spread, bap, bread roll, crisp bread, other bread, double-crust  pastry, single-crust pastry, pizza, Indian snacks, croissant, danish pastry, ice-cream,  doughnut, chocolate bar, white chocolate, milk chocolate, chocolate-covered raisin, chocolate  sweet, diet sweets, sweets, chocolate-covered biscuits, chocolate biscuits, sweet biscuits,  cereal bar, other sweets, crisps, savoury biscuits, cheesy biscuits, canned soup, sausage,  crumbed deep-fried poultry, breaded fish, Quorn, vegetarian sausages and burgers, powdered  instant soup, sugar added to coffee, sweetener added to coffee, sugar added to tea, sweetener  added to tea, sugar added to cereal, and sweetener added to cereal. | SJT is supported by a Postgraduate Scholarship from the National Health and Medical Research Council of Australia |
| Golzarand-2024 | Iran | NR | - industrial breads - processed meat and dairy - salty snacks - sweets and pastries - industrial fat products - soft drinks | mass-produced breads, pizza, sausages, hot dogs, burgers, lunch meat, chocolate milk, cream cheese, ice cream, mayonnaise, hydrogenated fat, margarine, cakes, biscuits, crackers, candies, chocolates, carbonated beverages, confectionaries, chips, cheese puffs, and toasted bread | Not specified |
| Rivera- 2024 | USA | NR | Not reported but referenced Du 2021   - Sugar‐sweetened beverages - Red and processed meat - Cold breakfast cereal - Fried foods - Sugary snacks - Baked goods - Dairy products - Alcohol - Margarine | Not reported but referenced Du 2021 | funded in whole or in part with federal funds from the National Heart, Lung, and Blood Institute, National Institutes of Health, US Department of Health and Human Services |
| Bonaccio- 2021 | Italy | NR | - Processed meat - Pizza - Cakes, pies, pastries, puddings (non-milk based) - Ice-cream - Crispbread/rusks - Fruit yoghurts - Dry cakes, biscuits - Snacks - Chocolate - Breakfast cereals - Candy bars - Confectionery non chocolate - Margarine - Salty biscuits, aperitif biscuits, crackers - Mayonnaise and similar - Fruit drinks - Carbonated/soft/isotonic drinks, diluted syrups - Spirits, brandy | Processed meat, Cakes, pies, pastries, puddings(non-milk based),Fruit drinks, Pizza (not homemade),Ice-cream, Crispbread/rusks, Fruit yoghurts, Carbonated/soft/isotonic, drinks, diluted syrups, Dry, cakes, biscuits, Savoury snacks, Stock cube, Chocolate, Breakfast cereals, Spirits, brandy Spreadable cheese, Nut spread, Confectionery non chocolate , Sliced cheese, Margarine , Salty biscuits, aperitif biscuits, crackers, Artificial sweeteners, Mayonnaise and similar | Supported by the Associazione Cuore Sano Onlus |
| Mendonca- 2017 | Spain | NR | - carbonated drinks, - processed meat, - biscuits [cookies], - candy [confectionery], - ‘instant’ packaged soups and noodles, - sweet or savory packaged snacks, and - sugared milk and fruit drinks | NR | Received  funding from the Spanish Government-Instituto de Salud  Carlos II and European Regional Development Fund; the Navarra Regional Government; and the University of Navarra. |
| Shuai Yuan- 2023 | UK | NR | NR | Low calorie drink intake, Fizzy drink intake carbonated (fizzy) drinks, Squash intake, Instant coffee intake, Intake of artificial sweetener added to coffee, Intake of artificial sweetener added to tea , Flavoured milk intake, Fortified wine intake , Spirits intake , Porridge intake, muesli intake, Oat crunch intake, Sweetened cereal intake , Plain cereal intake, Bran cereal intake , Whole-wheat cereal intake , Other cereal intake, Bap intake , Bread roll intake , Naan bread intake, Crispbread intake, Oatcakes intake, Other bread intake, Number of bread slices with butter/margarine, Number of baguettes with butter/margarine, Number of baps with butter/margarine, Number of bread rolls with butter/margarine, Number of crackers/crispbreads with butter/margarine, Number of oatcakes with butter/margarine, Number of other bread types with butter/margarine, Double crust pastry intake, Single crust pastry intake, Crumble intake, Pizza intake, Pancake intake, Scotch pancake intake, Yorkshire pudding intake, Indian snacks intake, Croissant intake, Danish pastry intake, Scone intake, Ice-cream intake , Milk-based pudding intake, Other milk-based pudding intake, Soya dessert intake, Fruitcake intake, Cake intake, Doughnut intake, Sponge pudding intake, Cheesecake intake, Other dessert intake, Chocolate bar intake , White chocolate intake, Milk chocolate intake, Dark chocolate intake, Chocolate-covered raisin intake, Chocolate sweet intake, Diet sweets intake, Sweets intake, Chocolate-covered biscuits intake, Chocolate biscuits intake, Sweet biscuits intake ,Cereal bar intake, Other sweets intake sweet snacks, Crisp intake, Savoury biscuits intake, Cheesy biscuits intake, Other savoury snack intake, Powdered/instant soup intake, Snackpot intake (snack pot, noodles/rice, Couscous intake, Low fat cheese spread intake, Cheese spread intake , Sausage intake, Crumbed or deep-fried poultry intake (chicken or turkey in breadcrumbs or deep fried , Bacon intake, Ham intake, Vegetarian sausages/burgers intake, Quorn intake, Other vegetarian alternative intake, Baked bean intake, Fried potatoes intake, Mashed potato intake | Funded by the Karolinska Institutet’s Research Foundation Grants, the Swedish Research Council, the Swedish Research Council for Health, Working Life and Welfare and the Swedish Heart-Lung Foundation. |
| Rauber- 2024 | UK | NR | - Sausage and other reconstituted red meat products - Nuggets and other reconstituted meat products - Milk based desserts - Mayonnaise and spreadable cheese | Slice bread, Bread roll, bap, burger bun, hotdog roll, bagel; Crackers, crispbread, rice cakes, corn cakes, Double or single crust pie/flan; Pancake, crêpe; Yorkshire pudding; Croissant; Scone (plain, fruit, cheese); Fruit cake, Cake, muffin, flapjack, brownie; Doughnuts; Sponge pudding; Cheesecake, Chocolate covered biscuits; Chocolate biscuits; Sweet biscuits; Cereal bars manufactured, Olive based spread; Margarine; Chocolate/nut spread, Potatoes (fried, chips, wedges, roast), Chocolate bars; Chocolate sweets; Low sugar / sugar free sweets (hard and soft); Sweets (hard and soft, e.g. peppermints, toffees, fudge, fruit flavoured sweets), Sweetened oat crunch type cereal; extruded plain cereals with sugars, Fruit drinks and fruit juices, soft drinks, other beverages, Low calories drinks (soft or fruit drinks), Crisps; Savoury crispbread/corn cake snacks; Cheesy biscuits, Industrial pizza, Dried/powdered soup; Carton/pouch/canned soup (pea, bean, lentil, vegetables, pasta); Snack pot, noodles/rice; baked beans, Spirits (e.g. vodka, whisky, gin, rum); other alcoholic drinks (e.g. Punch), Yeast extract; Tomato ketchup; Brown sauce/BBQ sauce; Salad dressing; Tomato-based sauce (e.g. pasta sauce); Gravy, Vegetarian sausage/burger; Tofu/tempeh/TVP/soya mince; Quorn, Sausage; Ham/Parma ham/salami/pastrami/cured meats, Chicken or turkey in breadcrumbs or deep fried; Liver or liver pâté; Breaded fish (e.g. fish fingers) or fish cakes; Battered fish, Ice-cream; Custard, rice pudding; Other milk-based desserts (e.g. mousse, tiramisu, crème caramel), Mayonnaise/salad cream (including low fat); Cheese sauce (e.g. cauliflower cheese); White sauce/cream sauce (e.g. bechamel) | Funding obtained from World Cancer Research Fund |
| Hyun-JuKim- 2019 | USA | NR | NR | Chocolate milk, ice cream, ice milk, milkshakes, bacon, sausage, processed meats, sweetened cereals, spaghetti/pasta with tomato sauce, cheese dishes, pizza, calzone, lasagna, salted snacks, cakes, cookies, brownies, fruit juices, sugar-sweetened and artificially sweetened beverages (Hi-C, Tang, Koolaid, diet colas, diet sodas, regular colas and sodas), hard liquor, margarine | supported by a Mentored Research Scientist Development Award from the National Institute of Diabetes and Digestive and Kidney Diseases |
| Du- 2024 | Sweden | NR | - starchy foods and breakfast cereals, - beverages (i.e., soft drinks), - sugary products, - fats and sauces, - meat and fish, - dairy products, and - salty snacks | Soft drinks, sweets/candies, chocolate, snacks, cookies, cakes, ice cream, margarine, mayonnaise, wafers, wheat crusts, low-fiber bread, marmalade, jam, gruel, breakfast cereals, tomato ketchup, nutritional powder, industrial soups, fish sticks, sausage, bacon and ham, blood pudding | funded by Crafoord Foundation, Magnus Bergvall Foundation, Albert Påhlsson Foundation, Swedish Research Council and Heart and Lung Foundation |
| Li- 2023 | Sweden | NR | NR | Soft drinks, sweets/candies, chocolate, snacks, cookies, cakes, ice cream, margarine, mayonnaise, wafers, wheat crusts, low-fiber bread, marmalade, jam, gruel, breakfast cereals, tomato ketchup, nutritional powder, industrial soups, fish sticks, sausage, bacon and ham, blood pudding | funded by Crafoord Foundation and Albert Påhlsson Foundation |
| Zhao- 2024 | USA and UK | 2 dieticians divided | NR | USA-PLCO: Soft drinks, diet/caffeinated; Soft drinks, regular/caffeinated; Soft drinks, diet/decaffeinated; Soft drinks, regular/decaffeinated; Hot dogs, turkey/low fat; Hot dogs, regular; English muffins and bagels; White bread/rolls; Biscuits; Donuts, sweet rolls, Danishes, and pop tarts; Pancakes, waffles, and French toast ; Ready-to-eat cereal, highly fortified; Ready-to-eat cereal, other; Cookies and brownies; Cakes, low fat; Cakes, regular; Lasagna, ravioli, shells; Pies, cream/custard/other; Pies, fruit a ; Pies, pecan; Pies, pumpkin/sweet potato; Granola bars; Pizza without meat; Meal replacement bar; Meal replacement, liquid; Puddings/custards; Croissants; Muffins/dessert breads, reg; Muffins/dessert breads, low-fat; Cheesecake; Fried potatoes; Fruit drinks, diet; Fruit drinks, regular; Whipped Cream, substitute; Whipped Cream, regular; Cream cheese, low fat; Cream cheese, regular; Ice cream/ice milk, low fat; Regular ice cream; Frozen yogurt, ices, and sorbet; Creamed soups; Milk, soy, in cereal; Milk, soy, not in coffee/tea; Milk, soy in coffee or tea; Milk, rice, in cereal; Milk, rice, not in coffee/tea; Milk, rice in coffee or tea; Milkshakes/sodas; Fried chicken, dark meat/no skin; Fried chicken, dark meat/with skin; Fried chicken, white meat/no skin; Fried chicken, white meat/with skin; Pizza with meat; Sausage; Bacon, regular; Bacon, lean/Canadian; Burgers; Gravy; Salad dressing, low fat on salad and vegetables; Salad dressing, nonfat on salad and vegetables; Salad dressing, regular on salad and vegetables; Mayonnaise, diet on salad; Mayonnaise, fat free on salad; Mayonnaise, regular on salad; Mayonnaise, diet on sandwich; Mayonnaise, fat free on sandwich; Mayonnaise, regular on sandwich; White sauce; Crackers; Potato/corn/other chips-low fat; Potato chips with olestra ; Potato/corn/other chips; Popcorn; Candy, chocolate; Candy, not chocolate; Saccharine in coffee and tea; Margarine, diet; Margarine, fat free; Margarine, regular; Margarine, diet on bread;  USA-NHANES: Flavoured milk; Ice cream; Vanilla yogurt; Fruit yogurt; Milk shake; Cream substitute; Sour cream; Pudding; Mousse; Cheese spread; bacon; sausage; sweetened cereals; Processed meats; Egg substitute; Cakes; Sweetened beverages; Margarine; Biscuits; Salted;  UK:  Low calorie drink; Fizzy drink; Squash; instant coffee; Cappuccino; Latte; Espresso; Flavoured milk; Other drink; Low calorie hot chocolate; Hot chocolate; Oat crunch; Sweetened cereal; Scotch egg; Sliced bread; Bap; Bread roll; Oatcakes; Other bread; Snackpot; Double crust pastry; Single crust pastry; Pizza; Pancake; Scotch pancake; Yorkshire pudding; Indian snacks; Croissant; Danish pastry; Scone; Yogurt a; Ice-cream; Milk-based pudding; Other milk-based pudding intake; Soya dessert; Fruitcake; Cake; Doughnut; Sponge pudding; Cheesecake; Other dessert; Chocolate bar; White chocolate; Milk chocolate; Dark chocolate; Chocolate-covered raisin; Chocolate sweet; Diet sweets; Sweets; Chocolate-covered biscuits; Chocolate biscuits; Sweet biscuits; Other sweets; Savoury biscuits; Cheesy biscuits; Other savoury snack; Powdered/instant soup; Canned soup; Sausage; Crumbed or deep-fried poultry; Bacon; Vegetarian sausages/burgers; Quorn; Other vegetarian alternative; Fried potatoes | No financial disclosures were reported by the authors of this paper. |
| Jalali- 2024 | Iran | NR | - Breads - Salty snacks - Fast foods - Sweetened beverages - Sweets and desserts - Dairy - Others | Baguette bread, Toast bread; Crackers, Puff, chips; French fries, Hamburger, Kielbasa, Sausage, Pizza; Industrial beverages, cola; Cookies, Yazdi cake, Homemade cakes, other cakes, industrial jams, dried sweets, cream sweets, GAZ, Candies, SOHAN, Chocolate, Caramel cream, sesame pudding, NOGHL, Donuts; Cacao milk, yogurt cream, cream cheese, traditional ice cream, non-traditional ice cream; Margarine, Ketchup, Mayonnaise | No financial disclosures were documented. |
| Nilson- 2022 | Brazil | NR | - Margarine; - Salted crackers; - Breads; - Cookies; - Hams; - Confectionary deserts; - Soft drinks; - Hotdogs, hamburgers and sandwiches; - Sweetened dairy drinks; - Pizza; - Fried and baked snacks; - Other sugar sweetened beverages; - Ready or semi-ready meals; - Sauces; - Cakes and pies; - Others | NR | No financial disclosures were reported by the authors of this paper. |
| Mitra- 2024 | Minnesota, USA | NR | NR | more than 30 non-ultra-processed plant food included in the intervention (not mentioned). | funded by an innovative scholarship award from St. Catherine University to MB and AM. |
| Rezende-Alves 2021 | Brazil | NR | - Ultra-processed dairy products - Sausages - Ultra-processed breads - Margarine - Sweetened beverages - Distilled alcoholics beverages - Ultra-processed fast foods and sweets | Plain curd, light curd, plain yogurt, light/low fat yogurt  Bologna/salami/fatty ham, turkey/Chester, sausages, frankfurter/sausage, bacon  Sliced white bread, toast, Brazilian cheese bread, sweet bread, whole bread (rye/wheat/oats) light bread, breakfast cereal  Margarine, light margarine/mayonnaise, mayonnaise  Soft drinks, diet/light/zero calories soft drinks, industrialized fruit juice (can/box/powder instant), diet/light industrialized juice  Vodka/rum/whisky, cachaça  Pizza, hot dog/red meat/chicken hamburger, fried finger foods (chicken croquet/pastry/risole/croquet), pastry/pie/quiche, popcorn, snacks such as industrialized chips, lasagne/cannelloni/rondelli, ice cream, light ice cream, soy milk, dark chocolate (50 – 70% cocoa), milk chocolate/ bonbon/Brazilian fudge balls, cereal bar, chocolate milk, pudding/ambrosia/dulce de leche/sweet rice/flan, sweet delicacies/maria-mole (a dessert popular in Brazil that is similar to a marshmallow)/merengue/candy , mustard | Financial support: This research was funded by FAPEMIG (Minas Gerais, Brazil). |
| Bonaccio-2021 | Italy | NR | - Processed meat - Pizza - Cakes, pies, pastries, puddings (non-milk based) - Ice-cream - Crispbread/rusks - Fruit yoghurts - Dry cakes, biscuits - Snacks - Chocolate - Breakfast cereals - Candy bars - Confectionery non chocolate - Margarine - Salty biscuits, aperitif biscuits, crackers - Mayonnaise and similar - Fruit drinks - Carbonated/soft/isotonic drinks, diluted syrups - Spirits, brandy | NR | By Associazione Cuore Sano Onlus (Campobasso, Italy |
| Rezende-Alves- 2023 | Brazil | NR | - Ultra-processed dairy products - Sausages - Ultra-processed breads - Margarine - Sweetened beverages - Distilled alcoholics beverages - Ultra-processed fast foods and sweets | Plain curd, light curd, plain yogurt, light/low fat yogurt  Bologna/salami/fatty ham, turkey/Chester, sausages, frankfurter/sausage, bacon  Sliced white bread, toast, Brazilian cheese bread, sweet bread, whole bread (rye/wheat/oats) light bread, breakfast cereal  Margarine, light margarine/mayonnaise, mayonnaise  Soft drinks, diet/light/zero calories soft drinks, industrialized fruit juice (can/box/powder instant), diet/light industrialized juice  Vodka/rum/whisky, cachaça  Pizza, hot dog/red meat/chicken hamburger, fried finger foods (chicken croquet/pastry/risole/croquet), pastry/pie/quiche, popcorn, snacks such as industrialized chips, lasagne/cannelloni/rondelli, ice cream, light ice cream, soy milk, dark chocolate (50 – 70% cocoa), milk chocolate/ bonbon/Brazilian fudge balls, cereal bar, chocolate milk, pudding/ambrosia/dulce de leche/sweet rice/flan, sweet delicacies/maria-mole (a dessert popular in Brazil that is similar to a marshmallow)/merengue/candy , mustard | This project was funded by FAPEMIG (Minas Gerais, Brazil). |
| Li -2024 | UK | NR | - Beverages - Dairy based products - Cereal and starchy food - Sugary products - Salty snacks - Meat - Spread and sauces | Low calorie drink, carboned drink, squash, fruit juice, fruit smoothie, dairy smoothie, low calorie hot chocolate, hot chocolate, spirits  Flavored milk, yogurt, ice-cream, milk-based pudding, other milk-based pudding, milk chocolate  Porridge, muesli, oat crunch, sweetened cereal, plain cereal, bran cereal, whole-wheat cereal, other cereal, slice bread, crispbread, pizza, snack pot  Soya dessert, fruitcake, cake, doughnut, sponge pudding, cheesecake, other dessert, chocolate bar, white chocolate, dark chocolate, chocolate-covered raisin, chocolate sweet, diet sweet, sweets, chocolate-covered biscuits, chocolate biscuits, sweet biscuits, cereal bar, other sweets  Crisp, savory biscuits, cheesy biscuits, other savory snack, powdered/instant soup, canned soup  Sausage, crumbed or deep-fried poultry, bacon, ham  Peanut butter, yeast extract, brown sauce, mayonnaise, low fat mayonnaise, salad dressing, olive-based spread, polyunsaturated margarine, dairy spread, soya margarine, unknown margarine, hard margarine, other margarine | funded by grants from National Natural Science Foundation of China |
| Li -2022 | China | NR | NR | NR | received no external funding. |
| Silva-2023 | Brazil | NR | - Soft drinks and artificial juices - Ultra-processed breads - Ultra-processed dairy products - Pasta, pizza, and snacks - Sweets and treats - Cakes and sweet biscuits - Sausages and processed meats - Salty biscuits and crackers - Margarine and mayonnaise - Instant soup - Cereal bars - Distilled beverages | Light bread, white/pita bread, sweet/homemade bread, whole grain/rye bread, Brazilian cheese bread, cake, stuffed cake, crackers, sweet biscuit w/ filling, sweet  biscuit w/o filling, light mayonnaise, regular mayonnaise, light yogurt, regular yogurt, light cream cheese, regular cream cheese, margarine, sausage/chorizo/Vienna sausage, hamburger (beef), light cold cuts, ham/mortadella/salami, pizza, instant  noodles, baked snacks, fried snacks, acarajé, hot dog, instant soup, ice cream, fruit popsicles, carame  l/candy, gelatin, chocolate powder, chocolate/bonbons/sweets, pudding/mousse, jam/jelly, cereal bars, diet soda, regular soda, processed juice w/  sugar, processed juice w/o sugar, processed juice w/ sweetener, artificial juice w/ sugar, artificial juice w/  o sugar, artificial juice w/ sweetener, whisky, vodka,  cachaça | supported by the Brazilian Ministry of  Health and the  Brazilian Ministry of Science, Technology and Innovation |
| Wang- 2023 | USA | NR | - HighFiberCereal_GDay - ColdCereal_GDay - Grits_GDay - HotCereal_GDay - WhiteBreads_GDay - DarkBreads_GDay - CornBreadTortilla_GDay - Biscuits_GDay - FriedPotatoes_GDay - BaconSausage_GDay - FriedBeef_GDay - OtherGroundBeef_GDay - BeefMixedDishes_GDay - Beef_GDay - FriedChicken_GDay - ChickenTurkey_GDay - ChickenMixedDishes_GDay - FriedSeafood_GDay - LunchMeats_GDay - HotdogSausage_GDay - MeatSubstitute_GDay - Margarine_Gday - RegDressingMayo_GDay - LowFatDressingMayo_GDay - Gravy_GDay - Cream_GDay - IceCream_GDay - FroYoSherbet_GDay - Cookies_GDay - Cake_GDay - PiesCobblers_GDay - DoughnutPastry_GDay - Chocolate_GDay - SaltySnacks_GDay - CrackersPretzels_GDay - Popcorn_GDay - SweetenedFruitDrinks_GDay - RegSoda_GDay - DietSoda_GDay - RegCoffee_GDay - DecafCoffee_GDay - Pizza_GDay - Soups_GDay - Hamburger_GDay - MacCheese_GDay - PeanutButter_GDay | Bran or high fiber cereals such as All Bran, Raisin Bran, or Shredded Wheat  Other cold cereals such as Corn Flake, Cheerios, or Product 19  Grits  Oatmeal, cream of wheat, or other hot cereals  White bread, rolls, dinner rolls, buns, or bagels (including sandwiches)  Dark or whole grain breads (including sandwiches)  Corn bread, corn muffins, corn tortillas, or hush puppies  Biscuits  Fried potatoes such as French fries, home fries, hash browns, or tater tots  Bacon or breakfast sausage (including breakfast sandwiches)  Fried beef (including chicken fried steak and steak with gravy)  Other ground beef such as meatloaf, meatballs, or patties  Beef in mixed dishes such as stew, pot pies, casserole, or stir fry  Roast beef, steak, or beef barbeque (including frozen dinners and sandwiches)  Fried chicken or chicken nuggets  Baked, broiled, or boiled chicken or turkey  Chicken in mixed dishes, casserole, stir-fries, or chicken pot pie  Fried fish, shrimp, or seafood (including sandwiches)  Bologna, salami, or other lunch meats  Hot dog sausage such as Kielbasa, Italian, Polish, Vienna, etc.  (do not include breakfast sausages)  Meat substitutes such as veggie-burgers, soy products, or tofu  Margarine (added to foods such as breads, grits, rice, and vegetables)  Regular salad dressing or mayonnaise (added to salads or sandwiches)  Low fat or reduced fat salad dressing or mayonnaise  Gravy added to potatoes, meat, or biscuits  Cream or whipped cream (including added to coffee or tea or on desserts)  Ice cream  Frozen yogurt, ice milk, or sherbet  Cookies  Cake  Baked or fried pies or cobblers  Doughnuts, sweet rolls, pastry, Danish, muffins, or croissants  Chocolate candy or candy bars  Potato chips, corn chips, fried pork skins, or cheese curls  Crackers or pretzels (including cheese and peanut butter crackers)  Popcorn  Fruit flavored drinks such as Kool-Aid, punch, Tang, lemonade, or Sunny Delight  Carbonated regular soft drinks such as Coke, Sprite, etc.  Diet coke, diet sodas, or other diet drinks  Regular coffee (brewed or instant)  Decaffeinated coffee (brewed or instant)  Pizza (homemade, frozen, or from a restaurant)  Soups or chowders such as tomato, vegetable, noodle, rice, etc.  Hamburgers, cheeseburgers, or sloppy joes  Macaroni and cheese  Peanut butter (include sandwiches) | supported by the National Cancer Institute of the National Institutes of Health |
| Mendoza- 2024 | USA | NR | - Ultra-processed bread and cereals - Sauces, spreads, and condiments - Packaged sweet snacks and desserts - Packaged savoury snacks - Sugar-sweetened beverages - Processed red meat, poultry, and fish - Ready-to-eat/heat mixed dishes - Yogurt and dairy based-desserts (information on UP yogurt is only available from 1994 and beyond) - Hard liquors - Artificially-sweetened beverages | Rye, pumpernickel bread  Whole-grain bread  English muffins, bagels, rolls  White bread  Cream cheese  Ketchup  Margarine  Mayonnaise (regular and low fat)  Non-dairy coffee whitener  Red chili sauce  Salad dressings  Salsa  Soy sauce  Spread butter  Candy bar with chocolate  Candy bar without chocolate  Chocolate bars  Dark chocolate bars  Breakfast bar  Brownies  Cookies, brownie, ready-made  Cookies, fat free, reduced fat  Doughnuts  Energy bars  High protein, low carb candy bars  Muffins or biscuits  Ready-made cakes  Ready-made cookies  Ready-made sweet rolls and coffee cakes  Applesauce  Canned peaches  Canned pears  Jams, jellies, preserves, honey  Ready-made pies  Fat-free popcorn  Fat-free, light crackers  Regular crackers  7-up  Coke or Pepsi with caffeine and sugar  Coke or Pepsi without caffeine but with sugar  Hawaiian punch with sugar  Other carbonated beverage  Dairy coffee drinks  Bacon  Beef, pork hotdogs  Breaded fish cakes, pieces, sticks  Chicken or turkey hot dogs  Processed meats, sausages  Salami, bologna, processed meat sandwiches  Chowder or cream soup  French fries potatoes  Pizza  Ready-made soup from cans  Soup made with bouillon  Artificially sweetened yogurt  Flavoured yogurt without Nutrasweet  Frozen yogurt  Ice cream  Ice cream  Sherbet  Liquors, including whiskey, gin, rum, and others  Caffeinated, caffeine-free, carbonated and non-carbonated low-calorie soda  Other low-calorie Cola with caffeine | The NHS, NHSII, and HPFS are supported by National Institutes of Health (NIH). |
| Dehghan-2023 | North and South America, Europe, Africa, the Middle East, and Asia | NR | NR | **North America and Europe**  **Canada**: Non-dairy creamer, milk shake, flavored yogurt, ice cream, fruit drinks, hot chocolate, pizza, French fries, margarine, ham, bacon, hot dog, sausages, luncheon ham, other luncheon meat, packaged breads, cold breakfast cereals, crackers, muffins, crisp snacks, cake, doughnuts, puddings, pies, tarts, cookies, chocolate, candy, gravy, ketchup, sauces, mayonnaise, sugar substitutes, soft drinks, salad dressings  **Sweden**: Ice cream, margarine, pizza, blood puddings, cold cuts, sausages, liver paste, luncheon meat, packaged breads, cold breakfast cereals, crisp bread, French fries, sweet rolls, cookies, biscuits, baked goods, chocolate, candy, sweets, chips, salad dressings, mayonnaise, tomato ketchup, marmalades, soft drinks  **Poland:**Ice cream, margarine, lard, Finea/Masmix, sausages, hot dog, luncheon meat, pork sausage, pork ham, slaska, krakowska, turkey ham, head cheese white and black, chicken pate, packaged breads, cold breakfast cereal, cakes, biscuits, halva with vanilla, sweets, drops, candy, chocolate, mayonnaise, salad with mayonnaise, fruit drinks, soft drinks | NR |
|  |  |  |  | **South America**  **Argentina**: Ice cream, margarine, bacon, blood sausage, hamburger, luncheon meat, Chile meat, cold breakfast cereal, cracker, packaged breads, ravioli, Gnocci, biscuit, lemon pie, alfajor chocolate tidbit, Budin, candy, Pasta Frola, chocolate, Dulce de leche, Bizocho, pastelito, facture, mayonnaise, soft drinks  **Brazil:** Ice cream, margarine, bacon, nuggets, pork-wieners, luncheon meat, Indlaids, Chile meat, cold breakfast cereal, cracker, packaged breads, cereal bar, brownies, candy, biscuits, stuffed biscuits, chocolate, cookies, mayonnaise, soft drinks  **Chile**: Margarine, salami, sausages, turkey ham, pate pork, cold breakfast cereal, packaged bread, cracker, Kuchen, candy, cookies, biscuits, chocolate, mayonnaise, soft drinks  **Colombia**: Margarine, chorizo, mortadella, salchicha, candy, cookies, packaged bread, cold breakfast cereal, French fries, cake, chocolate, biscuit, pastries, soft drinks |  |
|  |  |  |  | **Africa**  **South Africa:** Ice cream, margarine, sausages, hamburger, cold breakfast cereal, French fries, packaged snacks, fudge, chocolate, Mahewe, salad dressing, soft drinks  **Tanzania:** Margarine, deep fried foods, packaged bread, soft drinks  **Zimbabwe:** Margarine, cake, biscuits, candy, deep fried foods, packaged bread, Maheu, soft drinks |  |
|  |  |  |  | **Middle East**  **Iran:** Ice cream, margarine, sausages, kalbas, hamburger, samosa, French fries, cake, ghotab, sohan, biscuit, pofak, mayonnaise, sundis, delster, soft drinks  **Palestine**: Ice cream, pizza, cold cuts, cold breakfast cereal, packaged bread, French fries, biscuits, crackers, Arabic sweets, knafeh, hareesh, cake, hallaweh, energy drinks, soft drinks  **Turkey:** Ice cream, margarine, salami, sausages, pastrami, sucuk, pizza, French fries, ready made soups, cold breakfast cereals, fried sweets, halva, cake, pastries with/without meat, packaged bread, chips, crackers, biscuits, candy, chocolate, Gofret, soft drinks  **Saudi Arabia**: Ice cream, margarine, biscuits, cold breakfast cereal, cookies, mayonnaise, French fries, deep fried foods (e.g. samosa, falafel), pizza, biscuits, packaged bread, Arabic sweets, Aaseedah, Arabic Omani halva, Balaleet, Betheeth, kanafa, Khabeesah, Lagaimaat, Mahlabiyyeh, Halwa tahina, chocolate, soft drinks  **UAE:** Ice cream, margarine, deep fried foods (e.g. samosa, falafel), French fries, pizza, cracker, cookies, cake, mayonnaise, chocolate, ghee, cold breakfast cereal, packaged bread, Arabic sweets, Balaleet, Betheeth, Elba, kanafa, Lagaimaat, Mahlabiyyeh, Halwa tahina, Mehaiwah, soft drinks |  |
|  |  |  |  | **South Asia**  **Bangladesh:** Ice-cream, margarine, hamburgers, French fries, cold breakfast cereal, chocolates, candy, cakes, cookies, halwa, samosa, pakora, shingara, mayonnaise, soft drinks  **India:** Ice-cream, sausages, salty biscuit, sweet biscuits, chocolate, namkeen, payasam kheer, puddings, cake, pastries, candy, other Indian sweets, soft drink  **Pakistan:** Margarine, hamburger, pizza, halwa, biscuits, cakes, custard, zarda, brownies, toffee, chocolate, soft drinks |  |
|  |  |  |  | **South East Asia**  **Malaysia:** luncheon meat, hamburger, nuggets, sausages, cokodok pisang, ham, crackers, biscuit, packaged bread, curry puff, cucur udang, sri muka/lopes/lapis, chocolate drink, keropok, chocolate, oyster sauce, soy sauce, chili sauce, malted drink, soft drinks  **Philippines:** Ice cream, soy milk powder, margarine, processed meats, pizza, cake, biscuit, cracker, chips, curis, instant noodles, canton, Chicharron, Lechon, street foods, candy, chocolate, mayonnaise, salad dressing, Halo-Halo, toyo sauce, ketchup, bagoong, patis, soft drinks  **China:** Ice-cream, sausages, candy, cake, soy sauce, soft drinks |  |
| Juul 2021 | North America | NR NR | Bread, Sweets and desserts, Ultra-processed meats, Salty snack foods, Sugar-sweetened beverages, Low-calorie soft drinks, Fast foods, Breakfast cereals, Yoghurt, non-dairy coffee whitener, margarine, liquor, and chili sauce |  | NR |

**Supplementary Table 5.** Effect estimates prospective cohort studies examined the association between ultra-processed food consumption and cardiovascular diseases outcomes among adults

| **Author, Year** | **Outcome** | **Case** | **Estimate with 95% CI** | **Dose response** | |
| --- | --- | --- | --- | --- | --- |
|  |  |  |  | **Units**  **increased** | **Estimate (95% CI)** |
| Cordova 2023 | CVD | 10939 | 1.06 (1.04, 1.08) | 1SD (∼260 g/day) | 1.06 (1.04, 1.08) |
| Cordova 2023 |  | 10939 | 1.07 (1.04, 1.11) | Not reported | 1.07 (1.04, 1.11) |
| Zhong 2021 | CVD | 5490 | Q2: 1.00 (0.91, 1.09)  Q3: 1.01 (0.92, 1.11)  Q4: 1.2 (1.09, 1.31)  Q5: 1.5 (1.36, 1.64) | NA | NA |
|  |  | 5490 | Q2: 0.91 (0.81, 1.03)  Q3: 1.00 (0.89, 1.13)  Q4: 1.04 (0.92, 1.17)  Q5: 1.21 (1.07, 1.37) | NA | NA |
|  | Heart disease | 3985 | Q2: 1.01 (0.9, 1.12)  Q3: 0.99 (0.89, 1.11)  Q4: 1.26 (1.13, 1.41)  Q5: 1.68 (1.5, 1.87) | NA | NA |
|  |  | 3985 | Q2: 0.91 (0.79, 1.06)  Q3: 1.07 (0.92, 1.23)  Q4: 1.14 (0.98, 1.31)  Q5: 1.34 (1.16, 1.55) | NA | NA |
|  | Cerebrovascular disease | 1126 | Q2: 0.91 (0.76, 1.1)  Q3: 1.04 (0.86, 1.25)  Q4: 1.00 (0.82, 1.21)  Q5: 0.94 (0.76, 1.17) | NA | NA |
|  |  | 1126 | Q2: 0.85 (0.67, 1.09)  Q3: 0.74 (0.57, 0.96)  Q4: 0.79 (0.61, 1.02)  Q5: 0.87 (0.67, 1.13) | NA | NA |
| Chen 2022 | CVD | 6048 | Q2: 1.05 (0.97, 1.12)  Q3: 1.07 (0.99, 1.15)  Q4: 1.15 (1.07, 1.24) | NA | NA |
|  | Coronary heart disease | 5327 | Q2: 1.03 (0.96, 1.12)  Q3: 1.10 (1.01, 1.19)  Q4: 1.18 (1.08, 1.28) | NA | NA |
|  | Cerebrovascular disease | 1503 | Q2: 1.05 (0.91, 1.22)  Q3: 1.02 (0.88, 1.19)  Q4: 1.29 (1.1, 1.51) | NA | NA |
|  | CVD | 384 | Q2: 1.16 (0.86, 1.57)  Q3: 1.23 (0.91, 1.66)  Q4: 1.07 (0.77, 1.49) | NA | NA |
| Monge 2021 | Hypertension | 3752 | Q2: 0.96 (0.86, 1.07)  Q3: 0.92 (0.84, 1.02)  Q4: 0.95 (0.85, 1.06)  Q5: 0.98 (0.84, 1.14) | NA | NA |
|  |  | 3752 | Q2: 0.95 (0.85, 1.06)  Q3: 0.91 (0.82, 1)  Q4: 0.92 (0.83, 1.03)  Q5: 0.93 (0.8, 1.09) | NA | NA |
|  |  | 3752 | Q2: 0.95 (0.87, 1.03)  Q3: 1.04 (0.96, 1.13)  Q4: 0.96 (0.81, 1.14)  Q5: 1.12 (0.97, 1.3) | NA | NA |
| Du 2021 | Coronary Artery Disease | 2006 | Q2: 1.05 (0.92, 1.19)  Q3: 1.08 (0.95, 1.23)  Q4: 1.19 (1.05, 1.35) | 4.02 servings/d | 1.03 (0.74, 5.74) |
| Kityo 2023 | CVD-specific deaths | 539 | Q2: 0.97 (0.73, 1.29)  Q3: 0.99 (0.73, 1.34)  Q4: 0.9 (0.66, 1.25) | NA | NA |
|  | CVD-specific deaths (ICD-10 code I00-I99) | 539 | Q2: 0.67 (0.45, 0.99)  Q3: 0.96 (0.66, 1.38)  Q4: 0.81 (0.54, 1.21) | NA | NA |
| Srour 2019 | CVD | 1409 | Q2: 1.04 (0.91, 1.19)  Q3: 1.06 (0.92, 1.23)  Q4: 1.23 (1.04, 1.45) | 10% UPF(grams/day) | 1.12 (1.05, 1.20) |
|  | Myocardial infarction | 665 | Q2: 1.06 (0.87, 1.29)  Q3: 1.18 (0.96, 1.45)  Q4: 1.18 (0.93, 1.52) | 10% UPF(grams/day) | 1.12 (1.02, 1.24) |
|  | Cerebrovascular disease | 829 | Q2: 1.01 (0.85, 1.21)  Q3: 0.99 (0.82, 1.2)  Q4: 1.23 (1, 1.53) | 10% UPF(grams/day) | 1.11 (1.01, 1.21) |
| Li 2023 | CVD events (ICD-10 codes: CVD codes I20-I25 and I60-I64, I69) | 7006 | Q2: 1.02 (0.95, 1.09)  Q3: 1.03 (0.96, 1.11)  Q4: 1.19 (1.11, 1.27) | 10% UPF(grams/day) | 1.07 (1.04, 1.09) |
|  | CHD codes I20-I25 | 5217 | Q2: 1.08 (1, 1.16)  Q3: 1.11 (1.03, 1.2)  Q4: 1.24 (1.14, 1.34) | 10% UPF(grams/day) | 1.07 (1.04, 1.10) |
|  | Cerebrovascular disease | 2147 | Q2: 0.88 (0.78, 1)  Q3: 0.88 (0.78, 1)  Q4: 1.10 (0.97, 1.24) | 10% UPF(grams/day) | 1.06 (1.01, 1.11) |
|  | CVD death codes | 1124 | Q2: 0.98 (0.83, 1.16)  Q3: 0.98 (0.82, 1.17)  Q4: 1.46 (1.24, 1.73) | 10% UPF(grams/day) | 1.15 (1.08, 1.22) |
| Bonaccio 2023 | CVD mortality included deaths from diseases of the circulatory system when the underlying cause of death included ICD-9 codes 390–459. | 129 | Q2: 1.15 (0.66, 2)  Q3: 1.84 (1.07, 3.15)  Q4: 2.55 (1.53, 4.24) | 5% UPF(grams/day) | 1.45 (1.24, 1.70) |
|  | CVD mortality included deaths from diseases of the circulatory system when the underlying cause of death included ICD-9 codes 390–459. | 129 | Q2: 1.31 (0.78, 2.19)  Q3: 1.25 (0.72, 2.15)  Q4: 2.03 (1.22, 3.4) | 5% UPF(grams/day) | 1.03 (1.00, 1.06) |
| Wang 2024 | Incident cases of CVD was defined by hospital admission. Dates and causes for hospitalisation were identified by linking to Hospital Episode Statistics for England, Scottish Morbidity Records, and the Patient Episode Database for Wales. | 3055 | Q2: 1.07 (0.98, 1.17)  Q3: 1.13 (1.03, 1.23) | 10% UPF(grams/day) | 1.03 (1.00, 1.06) |
|  | Incident cases of hypertension was defined by hospital admission. Dates and causes for hospitalization were identified by linking to Hospital Episode Statistics for England, Scottish Morbidity Records, and the Patient Episode Database for Wales. | 6751 | Q2: 0.97 (0.91, 1.03)  Q3: 1.01 (0.95, 1.07) | 10% UPF(grams/day) | 1.01 (0.99, 1.04) |
|  | CMM was defined as the coexistence of at least two CMDs (T2D, CVD, and hypertension) | 3248 | Q2: 0.99 (0.91, 1.08)  Q3: 1.14 (1.05, 1.24) | 10% UPF(grams/day) | 1.07 (1.04, 1.10) |
| Sandra González‐Palacios 2023 | Systolic BP |  | Q2: 0.61 (0.02, 1.2)  Q3: 0.41 (-0.22, 1.05)  Q4: 0.45 (-0.25, 1.14) | 100 grm UPF | -0.18 (-0.37, 0.01) |
|  | Diastolic BP |  | Q2: 0.47 (0.15, 0.8)  Q3: 0.37 (0.02, 0.72)  Q4: 0.67 (0.29, 1.06) | 100 grm UPF | 0.02 (-0.09, 0.12) |
| Bonaccio 2022 |  | 792 | 1.07 (0.9, 1.28) | NA | NA |
|  |  | 792 | 1.21 (0.99, 1.47) | NA | NA |
|  |  | 792 | 1.27 (1.02, 1.58) | NA | NA |
|  | Cerebrovascular disease mortality | 426 | 1.14 (0.9, 1.44) | NA | NA |
|  |  | 426 | 1.22 (0.93, 1.61) | NA | NA |
|  |  | 426 | 1.39 (1.03, 1.88) | NA | NA |
|  | Cardiovascular mortality included deaths from diseases of the circulatory system, when the underlying cause of death included ICD-9 codes 390-459. | 792 | 1.33 (1.05, 1.67) | NA | NA |
| Fang 2024 | CVD mortality (ICD-8 codes 390-459) | 11416 | Q2: 1.03 (0.98, 1.09)  Q3: 1.01 (0.96, 1.07)  Q4: 1.05 (0.99, 1.11) | 4.4 servings/day (median) | 1.04 (0.99, 1.10) |
|  | CVD mortality (ICD-8 codes 390-459) | 6200 | Q2: 0.99 (0.92, 1.06)  Q3: 1.01 (0.94, 1.08)  Q4: 1.04 (0.97, 1.12) | 4.1 servings/day (median) | 1.05 (0.98, 1.13) |
|  | CVD mortality (ICD-8 codes 390-459) | 5216 | Q2: 1.09 (1.01, 1.18)  Q3: 1.02 (0.94, 1.11)  Q4: 1.06 (0.98, 1.15) | 4.8 servings/day (median) | 1.04 (0.96, 1.13) |
| Bonaccio 2021 | CVD mortality included underlying cause of death ( ICD-9 codes 390–459). | 439 | Q2: 1 (0.78, 1.28)  Q3: 1.10 (0.86, 1.41)  Q4: 1.58 (1.23, 2.03) | NA | NA |
|  | cerebrovascular disease and ischemic heart disease cause of death ( ICD-9 codes 430–438 and ICD-9 codes 410–414 and 429) | 255 | Q2: 1.03 (0.75, 1.42)  Q3: 1.02 (0.74, 1.41)  Q4: 1.52 (1.1, 2.09) |  |  |
| Torres-Collado 2024 | CVD (ICD-10: I00–I99) | 114 | Q2: 1.03 (0.66, 1.6)  Q3: 1.39 (0.8, 2.41) | 10% | 1.19 (0.9, 1.58) |
| Kermani-Alghoraishi 2024Iran | Cardiovascular events | 819 | Q2: 1.05 (0.88, 1.27)  Q3: 1.01 (0.83, 1.24)  Q4: 1.08 (0.88, 1.34) | NA | NA |
|  | MI +UA | 548 | Q2: 1.13 (0.9, 1.43)  Q3: 1.12 (0.88, 1.44)  Q4: 1.12 (0.87, 1.46) |  |  |
|  | Stroke | 172 | Q2: 1.09 (0.75, 1.6)  Q3: 0.89 (0.57, 1.38)  Q4: 0.93 (0.58, 1.47) |  |  |
|  | CVD mortality | 181 | Q2: 0.91 (0.62, 1.32)  Q3: 0.79 (0.51, 1.22)  Q4: 0.95 (0.61, 1.47) |  |  |
| Sullivan 2023 | CVD | 406 | Q2: 1.02 (0.79, 1.31) | NA | NA |
|  |  |  | Q3: 1.09 (0.85, 1.4) |  |  |
| Carmen Romero Ferreiro 2022 | CVD mortality | NR | Q2: 1.03 (0.98, 1.08) | NA | NA |
|  |  |  | Q2: 1.14 (1.01, 1.29) |  |  |
| Passinho 2023 | High CVR (cardiovascular risk | NR | Q2: 1.52 (0.52, 4.43)  Q3: 3.15 (1.24, 8.02)  Q4: 2.33 (0.91, 5.94)  Q5: 2.76 (1.1, 6.88) | NA | NA |
|  | High CVR | NR | Q2: 0.9 (0.32, 2.56)  Q3: 2.05 (0.85, 4.91)  Q4: 2.12 (0.88, 5.12)  Q5: 3.38 (1.41, 8.09) |  |  |
|  | Medium CVR | NR | Q2: 1.99 (1.25, 3.18)  Q3: 2.00 (1.26, 3.18)  Q4: 1.47 (0.92, 2.35)  Q5: 1.52 (0.95, 2.44) |  |  |
|  | Medium CVR | NR | Q2: 1.47 (0.93, 2.31)  Q3: 1.07 (0.67, 1.72)  Q4: 1.56 (0.99, 2.46)  Q5: 1.85 (1.17, 2.93) |  |  |
| Orlich 2022 | CVD mortality | 3388 | Q5: 1.09 (1, 1.18) | NA | NA |
| Oladele 2024 | Hypertension | 2134 | Q2: 1.02 (0.87, 1.21)  Q3: 1.17 (0.99, 1.38)  Q4: 1.2 (1.01, 1.42) | NA | NA |
|  | Hypertension | 2134 | Q2: 1.01 (0.85, 1.19)  Q3: 1.02 (0.86, 1.21)  Q4: 1.15 (0.97, 1.39) |  |  |
| Pant 2024 | Incident CVD | 1,038 | Q2: 0.92 (0.71, 1.2)  Q3: 1.21 (0.93, 1.56)  Q4: 1.00 (0.76, 1.32)  Q5: 1.22 (0.92, 1.61) | NA | NA |
|  | Incident hypertension | 4204 | Q2: 1.26 (1.02, 1.55)  Q3: 1.26 (1.02, 1.56)  Q4: 1.32 (1.06, 1.65)]  Q5: 1.39 (1.1, 1.74) |  |  |
| Moreira 2022 | Reduction in CVD mortality | 215028 | Q2: 196436 (160973, 232858) | NA | NA |
|  | Reduction in CHD mortality | 115229 | Q2: 129381 (104489, 154874) | NA | NA |
|  | Reduction in Stroke mortality by 2048 (I60-I64 and I69) | 99799 | Q2: 67055 (56485, 77984) | NA | NA |
| Hang 2024 | CVD mortality | 335 | Q2: 1.47 (1.05, 2.06)  Q3: 1.01 (0.7, 1.46)  Q4: 1.6 (1.13, 2.27)  Q5: 1.65 (1.13, 2.4) | NA | NA |
| Tu 2023 | incident atrial fibrillation | 4579 | Q2: 1.03 (0.94, 1.13)  Q3: 1.06 (0.97, 1.17)  Q4: 1.03 (0.94, 1.14)  Q5: 1.13 (1.02, 1.24) | 10% | 1.05 (1.01, 1.08) |
| Golzarand 2024 | HTN | 688 | T2: 1.07 (0.88, 1.3)  T3: 1.48 (1.23, 1.79) | per 1-serving/day | 1.03 (1.01, 1.05) |
| Rivera 2024 | HTN | 7018 | Q2: 1.01 (0.94, 1.08)  Q3: 1.01 (0.95, 1.08)  Q4: 1.12 (1.04, 1.2) | per 1-serving/day | 1.02 (1.01, 1.03) |
| Bonaccio 2021b | CVD mortality | 178 | Q2: 0.87 (0.56, 1.35)  Q3: 1.24 (0.8, 1.91)  Q4: 1.65 (1.07, 2.55) | NA | NA |
|  | IHD/cerebrovascular | 114 | Q2: 0.82 (0.47, 1.45)  Q3: 1.4 (0.83, 2.38)  Q4: 1.65 (0.96, 2.85) |  |  |
| Mendonca 2017 | HTN | 1702 | T2: 0.99 (0.88, 1.12)  T3: 1.21 (1.06, 1.37) | NA | NA |
| ShuaiYuan 2023 | VTE | 4235 | Q2: 1.01 (0.91, 1.11)  Q3: 0.98 (0.88, 1.09)  Q4: 1.12 (1.01, 1.24)  Q5: 1.05 (0.94, 1.17) | 1 SD | 1.01 (0.98, 1.05) |
|  | deep vein thrombosis [DVT] | 1855 | Q2: 1.06 (0.92, 1.21)  Q3: 0.97 (0.84, 1.12)  Q4: 1.15 (1.01, 1.33)  Q5: 1.02 (0.88, 1.18) | 1 SD | 1.00 (0.95, 1.04) |
|  | pulmonary embolism [PE]) | 2380 | Q2: 0.94 (0.81, 1.11)  Q3: 0.98 (0.84, 1.15)  Q4: 1.07 (0.91, 1.25)  Q5: 1.09 (0.93, 1.28) | 1 SD | 1.03 (0.98, 1.08) |
|  | VTE : Incident venous thromboembolism risk defined by the International Classification of Disease (ICD)-9 and 10 codes | 4235 | Q2: 0.94 (0.85, 1.04)  Q3: 1.01 (0.92, 1.12)  Q4: 1.08 (0.98, 1.19)  Q5: 1.10 (1, 1.22) | 1 SD | 1.06 (1.03, 1.09) |
|  | Deep vein thrombosis [DVT] | 1855 | Q2: 0.93 (0.81, 1.06)  Q3: 1.00 (0.88, 1.14)  Q4: 0.97 (0.85, 1.11)  Q5: 1.05 (0.92, 1.19) | 1 SD | 1.04 (1.00, 1.08) |
|  | Pulmonary embolism [PE]) | 2380 | Q2: 0.94 (0.8, 1.1)  Q3: 1.03 (0.89, 1.21)  Q4: 1.22 (1.05, 1.41)  Q5: 1.19 (1.02, 1.38) | 1 SD | 1.09 (1.04, 1.14) |
|  | VTE : incident venous thromboembolism risk defined by the International Classification of Disease (ICD)-9 and 10 codes | 4235 | Q2: 1.02 (0.92, 1.12)  Q3: 0.99 (0.9, 1.09)  Q4: 1.09 (0.99, 1.2)  Q5: 1.15 (1.05, 1.27) | 1 SD | 1.05 (1.02, 1.08) |
|  | deep vein thrombosis [DVT] | 1855 | Q2: 1.05 (0.92, 1.19)  Q3: 0.96 (0.84, 1.1)  Q4: 1.11 (0.97, 1.26)  Q5: 1.12 (0.98, 1.27) | 1 SD | 1.04 (1.00, 1.08) |
|  | pulmonary embolism [PE]) | 2380 | Q2: 0.98 (0.84, 1.14)  Q3: 1.04 (0.9, 1.2)  Q4: 1.08 (0.93, 1.25)  Q5: 1.2 (1.04, 1.39) | 1 SD | 1.06 (1.01, 1.11) |
| Rauber 2024 | Incident cardiovascular disease | 7806 | Q2: 1.09 (1.02, 1.17)  Q3: 1.17 (1.1, 1.25)  Q4: 1.23 (1.15, 1.31) | 10% | 1.06 (1.04, 1.08) |
|  | Coronary heart disease | 6006 | Q2: 1.18 (1.09, 1.27)  Q3: 1.24 (1.15, 1.34)  Q4: 1.31 (1.21, 1.41) | 10% | 1.07 (1.05, 1.09) |
|  | Cerebrovascular disease | 2112 | Q2: 0.89 (0.79, 1.01)  Q3: 0.98 (0.87, 1.1)  Q4: 1.00 (0.88, 1.13) | 10% | 1.01 (0.98, 1.04) |
|  | CVD mortality | 529 | Q2: 1.3 (1, 1.68)  Q3: 1.35 (1.04, 1.74)  Q4: 1.42 (1.1, 1.84) | 10% | 1.09 (1.02, 1.16) |
|  | Coronary heart disease mortality | 348 | Q2: 1.49 (1.07, 2.07)  Q3: 1.64 (1.18, 2.27)  Q4: 1.65 (1.19, 2.28) | 10% | 1.13 (1.05, 1.23) |
|  | Cerebrovascular disease Mortality | 181 | Q2: 1.06 (0.7, 1.6)  Q3: 0.97 (0.63, 1.48)  Q4: 1.12 (0.75, 1.72) | 10% | 1.01 (0.90, 1.12) |
| Hyun-JuKim 2019 | CVD | 648 | Q2: 1.09 (0.69, 1.74)  Q3: 0.92 (0.6, 1.41)  Q4: 1.10 (0.74, 1.67) | NA | NA |
| Du 2024 | CVD | 3709 | Q2: 1.01 (0.9, 1.12)  Q3: 1.00 (0.89, 1.11)  Q4: 1.03 (0.92, 1.15)  Q5: 1.1 (0.99, 1.22) | 1sd (176.76 g/day) | 1.05 (1.01, 1.08) |
| Li 2023 | CVD | 6236 | Q2: 1.03 (0.95, 1.11)  Q3: 1.02 (0.94, 1.1)  Q4: 1.18 (1.08, 1.29) | 1SD (211g/day) | 1.07 (1.04, 1.11) |
|  | CHD | 3566 | Q2: 1.04 (0.94, 1.14)  Q3: 1.03 (0.92, 1.15)  Q4: 1.2 (1.07, 1.35) | 1SD (211g/day) | 1.07 91.03, 1.11) |
|  | Ischemic Stroke | 3272 | Q2: 1.03 (0.93, 1.14)  Q3: 1 (0.89, 1.12)  Q4: 1.17 (1.03, 1.32) | 1SD (211g/day) | 1.08 (1.04, 1.13) |
| Zhao 2024 | CVD | 11827 | Q2: 0.98 (0.92, 1.03)  Q3: 1.07 (1.02, 1.12)  Q4: 1.17 (1.06, 1.28) | NA | NA |
|  | CVD (ICD codes) | 8099 | Q2: 0.98 (0.92, 1.04)  Q3: 1.09 (1.02, 1.16)  Q4: 1.12 (1.05, 1.19) |  |  |
|  | CVD (ICD codes) | 1979 | Q2: 0.98 (0.86, 1.12)  Q3: 1.03 (0.9, 1.17)  Q4: 1.28 (1.13, 1.45) |  |  |
|  | CVD (ICD codes) | 1749 | Q2: 0.91 (0.74, 1.12)  Q3: 0.96 (0.78, 1.17)  Q4: 1.11 (0.92, 1.34) |  |  |
| Jalali 2024 | CVD incidents | 208 | T2: 1.12 (0.79, 1.63) | 50 gm/day | 1.22 (1.03, 1.45) |
|  |  |  | T3: 1.68 (1.14, 2.48) |  |  |
| Nilson 2022 | Premature CVD deaths | 19,200 | Q2: 19200 (7097, 32353) | NA | NA |
|  | IHD |  | Q2: 11400 (4301, 19220) | NA | NA |
|  | Stroke |  | Q2: 7800 (2795, 13133) | NA | NA |
| Mitra 2024 | SBP |  | Q2: 113.7 ± 11.3 vs 118.87 ± 11.39 (NA, NA) | NA | NA |
|  | DBP |  | Q2: 78.6 ± 7.91 vs 85.4 ± 7.05 (NA, NA) | NA | NA |
| Rezende-Alves 2021 | Hypertension | 370 | Q2: 1.22 (0.91, 1.64)  Q3: 1.15 (0.86, 1.56)  Q4: 0.99 (0.72, 1.36)  Q5: 1.35 (1.01, 1.82) | NA | NA |
| Rezende-Alves 2023 | Maintained or increased SBP versus decreased SBP |  | Q2: 1.02 (0.64, 1.63)  Q3: 1.09 (0.67, 1.77)  Q4: 1.15 (0.7, 1.88)  Q5: 0.88 (0.54, 1.42) | NA | NA |
|  | Increased SBP versus maintained SBP |  | Q2: 1.23 (0.78, 1.95)  Q3: 0.96 (0.59, 1.56)  Q4: 1.25 (0.78, 2.01)  Q5: 1.11 (0.68, 1.82) |  |  |
|  | Maintained or increased DBP versus decreased DBP |  | Q2: 0.86 (0.52, 1.43)  Q3: 0.66 (0.4, 1.1)  Q4: 0.85 (0.5, 1.44)  Q5: 0.90 (0.52, 1.54) |  |  |
|  | Increased DBP versus maintained DBP |  | Q2: 1.5 (0.94, 2.38)  Q3: 1.35 (0.84, 2.17)  Q4: 1.97 (1.25, 3.1)  Q5: 1.79 (1.12, 2.86) |  |  |
| Li 2024 | Overall cardiovascular disease | 1089 | Q2: 0.95 (0.8, 1.14)  Q3: 1.07 (0.9, 1.27)  Q4: 1.28 (1.08, 1.51) | Per 10% increase | 1.10 (1.04, 1.15) |
|  | Coronary heart disease | 829 | Q2: 0.97 (0.8, 1.18)  Q3: 1.06 (0.87, 1.29)  Q4: 1.28 (1.06, 1.56) | Per 10% increase | 1.10 (1.04, 1.16) |
|  | Stroke | 225 | Q2: 0.91 (0.63, 1.31)  Q3: 0.92 (0.63, 1.34)  Q4: 0.99 (0.68, 1.43) | Per 10% increase | 1.01 (0.9, 1.12) |
|  | Heart failure | 310 | Q2: 0.91 (0.65, 1.28)  Q3: 1.14 (0.83, 1.58)  Q4: 1.46 (1.07, 2) | Per 10% increase | 1.14 (1.05, 1.25) |
| Li 2022 | Hypertension | 4329 | Q2: 1 (0.9, 1.12)  Q3: 1.17 (1.03, 1.33)  Q4: 1.19 (1.06, 1.35) | NA | NA |
| Dehghan 2023 | Total mortality | **9227** | Q2: 1.08 (1.02, 1.15)  Q3: 1.18 (1.06, 1.31)  Q4: 1.28 (1.15, 1.42) | per serving | 1.05 (1.03, 1.07) |
|  | CV mortality | 3073 | Q2: 1.11 (1.01, 1.23)  Q3: 1.2 (1, 1.44)  Q4: 1.17 (0.98, 1.41) | per serving | 1.04 (1.01, 1.08) |
|  | Non-CV mortality | 6273 | Q2: 1.08 (1, 1.16)  Q3: 1.18 (1.04, 1.35)  Q4: 1.32 (1.17, 1.5) | per serving | 1.05 (1.02, 1.07) |
|  | Major CVD | 7934 | Q2: 0.99 (0.93, 1.05)  Q3: 1.01 (0.9, 1.13)  Q4: 1.01 (0.9, 1.12) | per serving | 1.01 (0.99, 1.03) |
| Silva 2023 |  |  | 1.10 (1.01, 1.19) | 10% increment | 1.10 (1.01, 1.19) |
|  | NCD mortality | 433 | 1.11 (1.02, 1.21) | 10% increment | 1.11 (1.02, 1.21) |
|  | CVD mortality | 143 | 0.97 (0.82, 1.14) | 10% increment | 0.97 (0.82, 1.14) |
| Wang 2023 |  |  |  |  |  |
|  | CHD mortality | 2208 | Q2: 0.98 (0.87, 1.1)  Q3: 0.97 (0.85, 1.1)  Q4: 0.88 (0.76, 1.01) | Per 10% increase | 0.96(0.93,1.00) |
|  | Stroke mortality | 867 | Q2: 1.12 (0.92, 1.36)  Q3: 1.09 (0.88, 1.35)  Q4: 1.15 (0.91, 1.44) | Per 10% increase | 1.04(0.99,1.10) |
|  | Diabetes mortality | 997 | Q2: 1.04 (0.86, 1.25)  Q3: 1.13 (0.94, 1.38)  Q4: 1.32 (1.07, 1.62) | Per 10% increase | 1.09(1.04,1.15) |
| Mendoza 2024 | Total cardiovascular disease | 16800 | Q2: 1.05 (1, 1.1)  Q3: 1.04 (0.99, 1.09)  Q4: 1.05 (1, 1.1)  Q5: 1.11 (1.06, 1.16) | NA | NA |
|  | Coronary heart disease | 10401 | Q2: 1.05 (0.99, 1.11)  Q3: 1.05 (0.99, 1.12)  Q4: 1.08 (1.01, 1.14)  Q5: 1.16 (1.09, 1.24) |  |  |
|  | Stroke | 6758 | Q2: 1.05 (0.97, 1.13)  Q3: 1.02 (0.95, 1.1)  Q4: 1.01 (0.94, 1.1)  Q5: 1.04 (0.96, 1.12) |  |  |
| Juul 2021 | Hard CVD | 251 | - | 2.9 SD servings/day  2.9 SD servings/day  2.9 SD servings/day  2.9 SD servings/day | 1.07 (1.03, 1.12)  1.09 (1.04, 1.15)  1.05 (1.02, 1.08)  1.09 (1.02, 1.16) |
|  | Hard CHD | 163 | - |  |  |
|  | Overall CVD morbidity | 648 | - |  |  |
|  | CVD mortality | 108 | - |  |  |

### **Supplementary Table 6**: Meta-regression results for the observed heterogeneity between studies in the association between ultra-processed food consumption and cardiovascular diseases outcomes

| term | Category | RR | 95% CI | P-value |
| --- | --- | --- | --- | --- |
| Intercept |  | 1.01 | 0.48, 2.14 | 0.977 |
| Region | Asia | Ref | Ref |  |
|  | European | 1.12 | 0.68, 2.04 | 0.659 |
|  | Middle East | 1.23 | 0.65, 2.33 | 0.300 |
|  | Global | 1.19 | 0.63, 2.26 | 0.498 |
|  | North America | 1.07 | 0.65, 1.59 | 0.953 |
|  | Oceania | 1.33 | 0.66, 2.19 | 0.542 |
|  | South America | 1.00 | 0.64, 1.71 | 0.867 |
| Sex | Both | Ref | Ref |  |
|  | Female | 0.90 | 0.53, 1.154 | 0.954 |
|  | Male | 1.11 | 0.62, 2.00 | 0.718 |
| UPF unit of analysis | Calorie | Ref | Ref |  |
|  | Weight | 0.94 | 0.73, 1.22 | 0.320 |
| Dietary Assessment | 24-hours recall | Ref | Ref |  |
|  | Mixed | 0.95 | 0.55, 1.63 | 0.676 |
|  | FFQ | 0.95 | 0.59, 1.54 | 0.641 |
| Outcome type | Cerebrovascular | Ref | Ref |  |
|  | CVD | 1.21 | 0.62, 2.39 | 0.574 |
|  | CHD | 1.45 | 0.73, 2.88 | 0.284 |
|  | Hypertension | 1.21 | 0.62, 2.39 | 0.574 |
|  | Thromboembolism | 1.05 | 0.37, 2.96 | 0.930 |
| Event type | Morbidity | Ref | Ref |  |
|  | Mortality | 0.92 | 0.82, 1.04 | 0.181 |
| Energy adjustment method | Residual and Standard | Ref | Ref |  |
|  | Standard | 1.04 | 1.002, 1.07 | 0.038 |
| Follow-up duration (in years) | ≥ 10 | Ref | Ref |  |
|  | < 10 | 0.98 | 0.84, 1.14 | 0.810 |

Studies from databases/registers **(n =** **24962)**

PsycINFO (n = 7059)

Embase (n = 5326)

Web of Science (n = 4728)

MEDLINE (n = 3148)

ClinicalTrials.gov (n = 2101)

CINAHL (n = 2049)

CENTRAL (n = 249)

Citation searching (n = 178)

Google Scholar (n = 124)

References from other sources **(n = )**

Citation searching (n = )

Grey literature (n = )

**Identification**

Included studies ongoing **(n = 0)**

Studies awaiting classification **(n = 0)**

Studies included in review **(n = 46)**

Studies excluded **(n = 12792)**

Studies not retrieved **(n = 0)**

Studies assessed for eligibility **(n = 248)**

Studies sought for retrieval **(n = 248)**

Studies screened **(n =** **13055)**

References removed **(n = 11907)**

Duplicates identified manually (n = 0)

Duplicates identified by Covidence (n = 11907)

Marked as ineligible by automation tools (n = 0)

Other reasons (n = )

**Screening**

Studies excluded **(n = 195)**

Preprint (n = 1)

Duplicate (n = 10)

Editorial (n = 6)

manuscript (n = 1)

Author Notes (n = 4)

RCT proposal (n = 1)

Wrong outcomes (n = 39)

Systematic review (n = 17)

Cross-sectional design (n = 2)

Full-text not available (n= 3)

Wrong intervention (n = 30)

Wrong study design (n = 50)

Conference proceeding (n = 20)

Paediatric population (n = 11)

Full text not available (n = 2)

Full text is not available (n = 4)

**Included**

**Supplementary Figure 1**. PRISMA

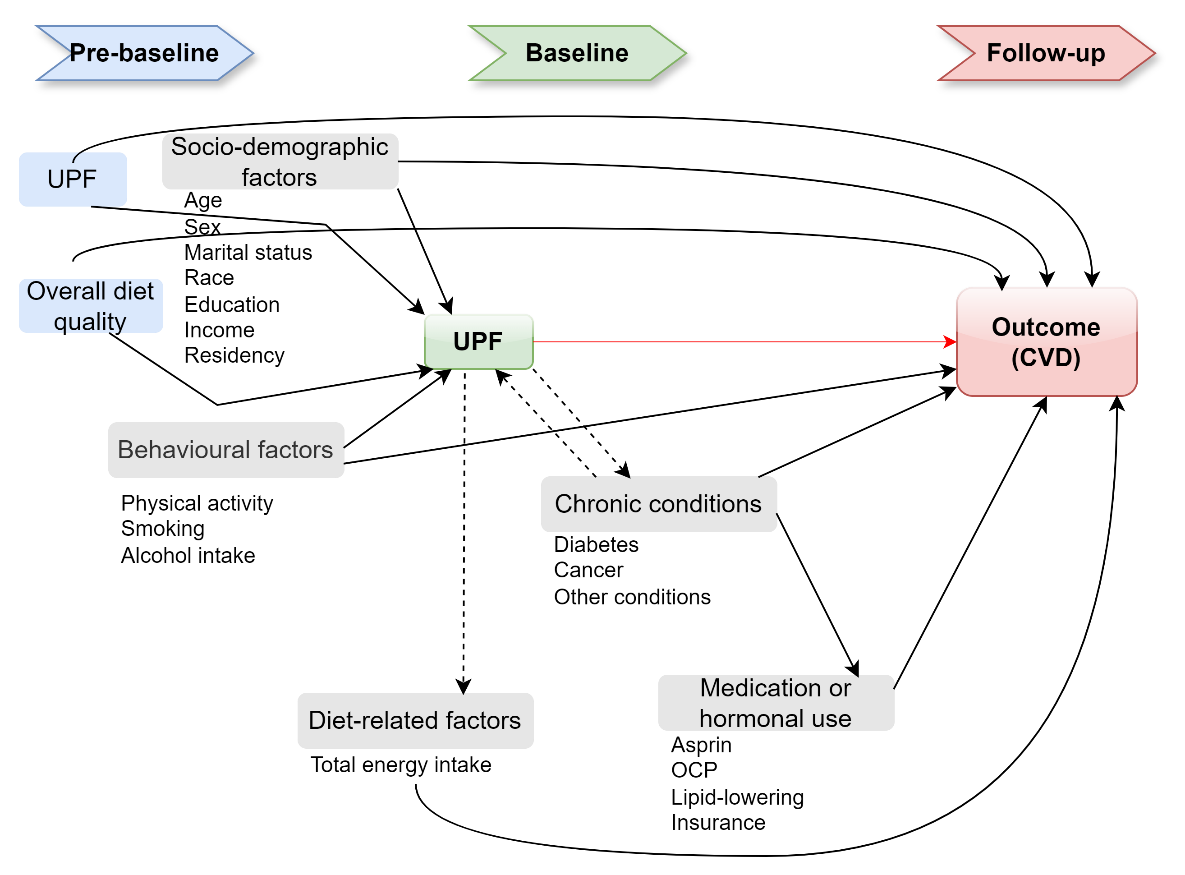

**Supplementary Figure 2**. Simplified direct acyclic graph for the association between ultra-processed food (UPF) and cardiovascular disease (CVD)

**Note**: Minimal adjustment of UPF–CVD models should include socio-demographic characteristics (age, sex, marital status, race, education, income, and residency), and behavioural factors (physical activity, smoking, and alcohol intake). Chronic conditions require careful consideration, as they may act as mediators; therefore, participants with CVD at baseline should be excluded. Adjustment for medication use may improve precision but does not alter the effect estimate itself. While adjusting for pre-baseline dietary exposure would be ideal, this approach was rarely applied in existing studies and was therefore not considered a necessary requirement for optimal adjustment. Similarly, adjusting for total energy intake is generally inappropriate due to its potential mediating effect; however, when studies did not use percentage of kilocalories or grams of intake, adjustment for total energy intake may be acceptable to maintain a constant caloric intake. Additionally adjusting for BMI, lipid profile markers, inflammatory and metabolic markers also can lead to a biased estimate of UPF and CVD outcomes.

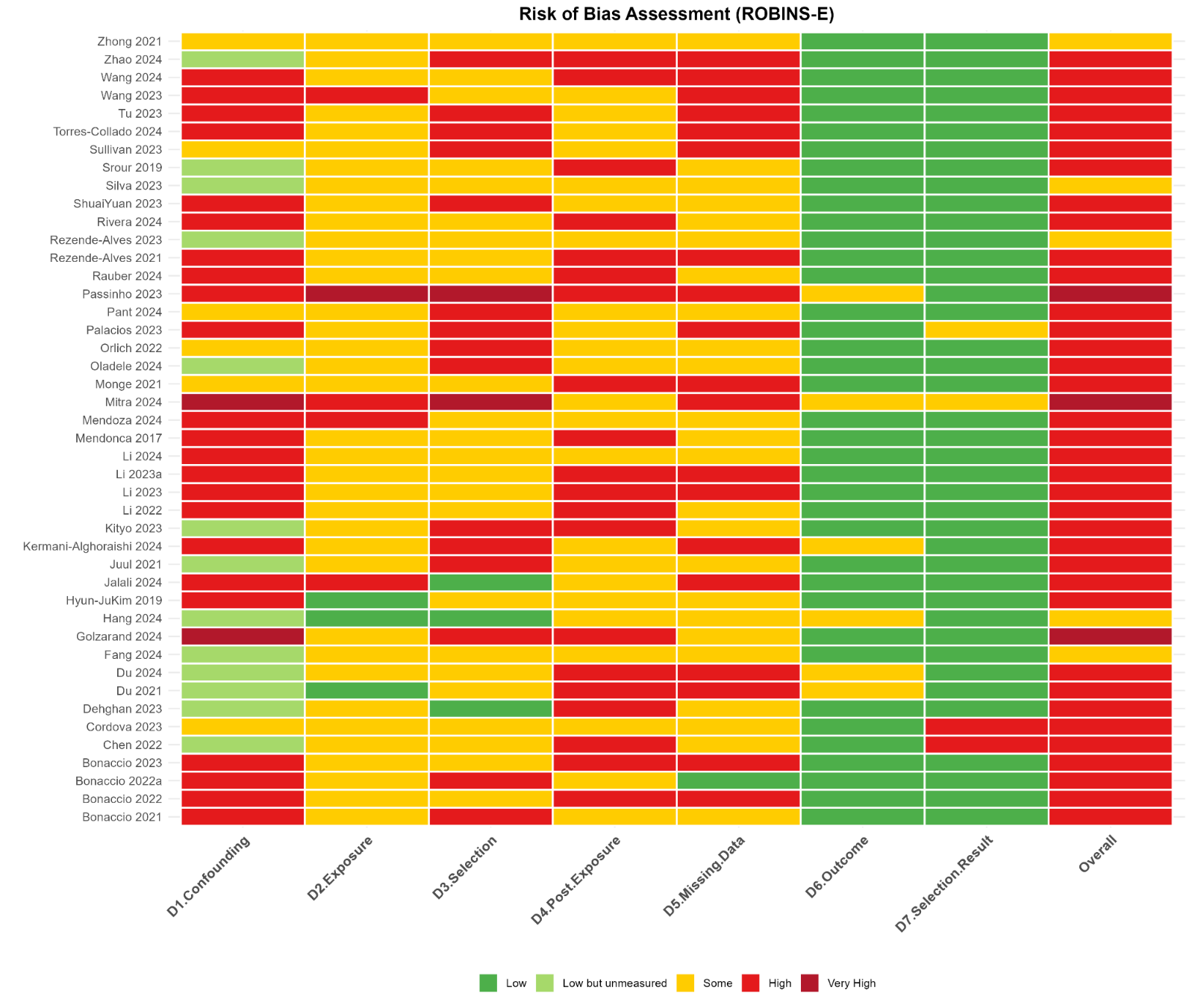

**Supplementary Figure 3.** Assessing the risk of bias of included studies for the ultra-processed food exposure and cardiovascular diseases and hypertension outcomes using the Risk Of Bias In Non-randomised Studies – of Exposures (ROBINS-E) tool

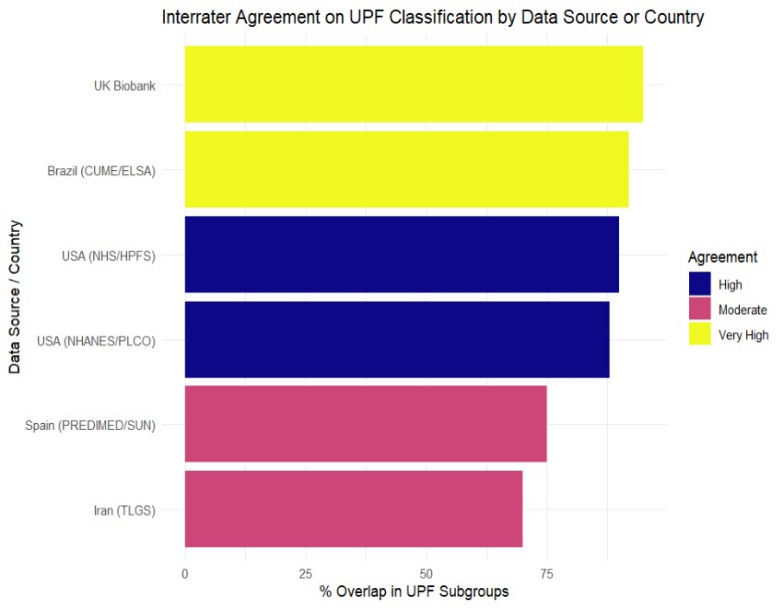

**Supplementary Figure 4**. Interrater agreement of ultra-processed foods within cohorts

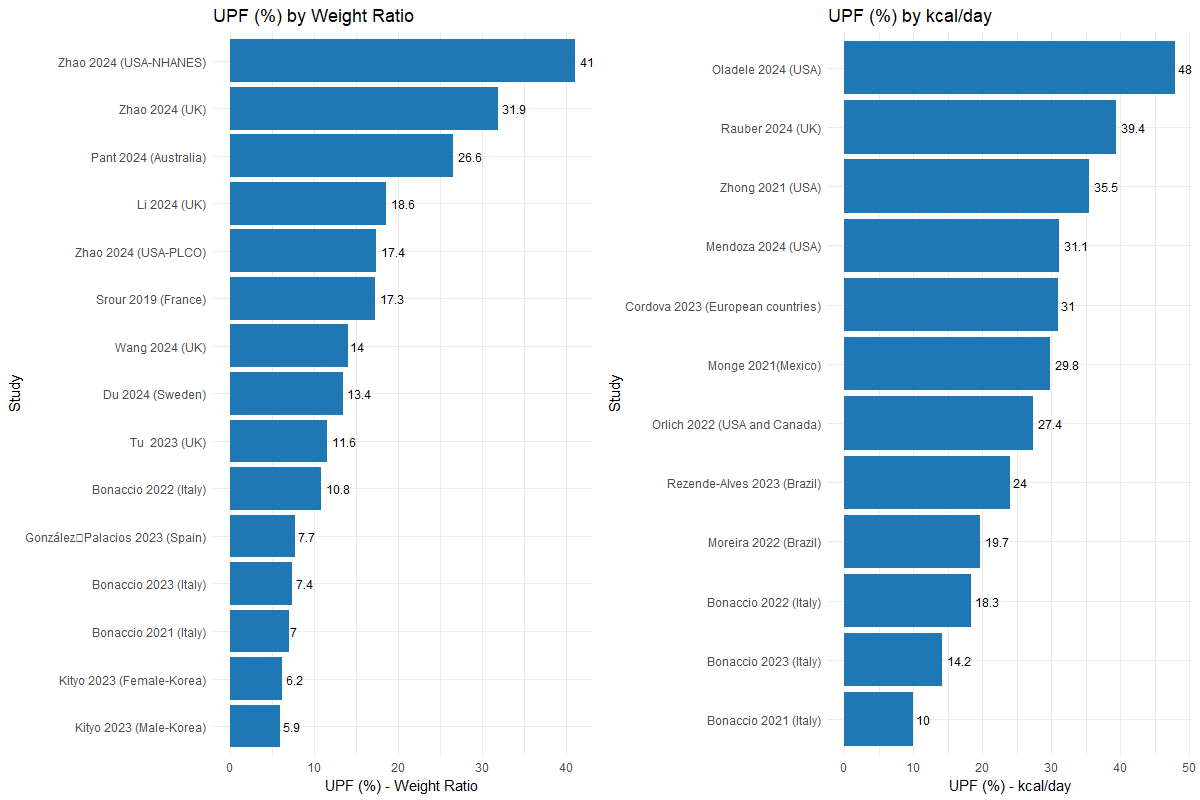

**Supplementary Figure 5.** Percentage of weight and energy intake from ultra-processed food by study (country)

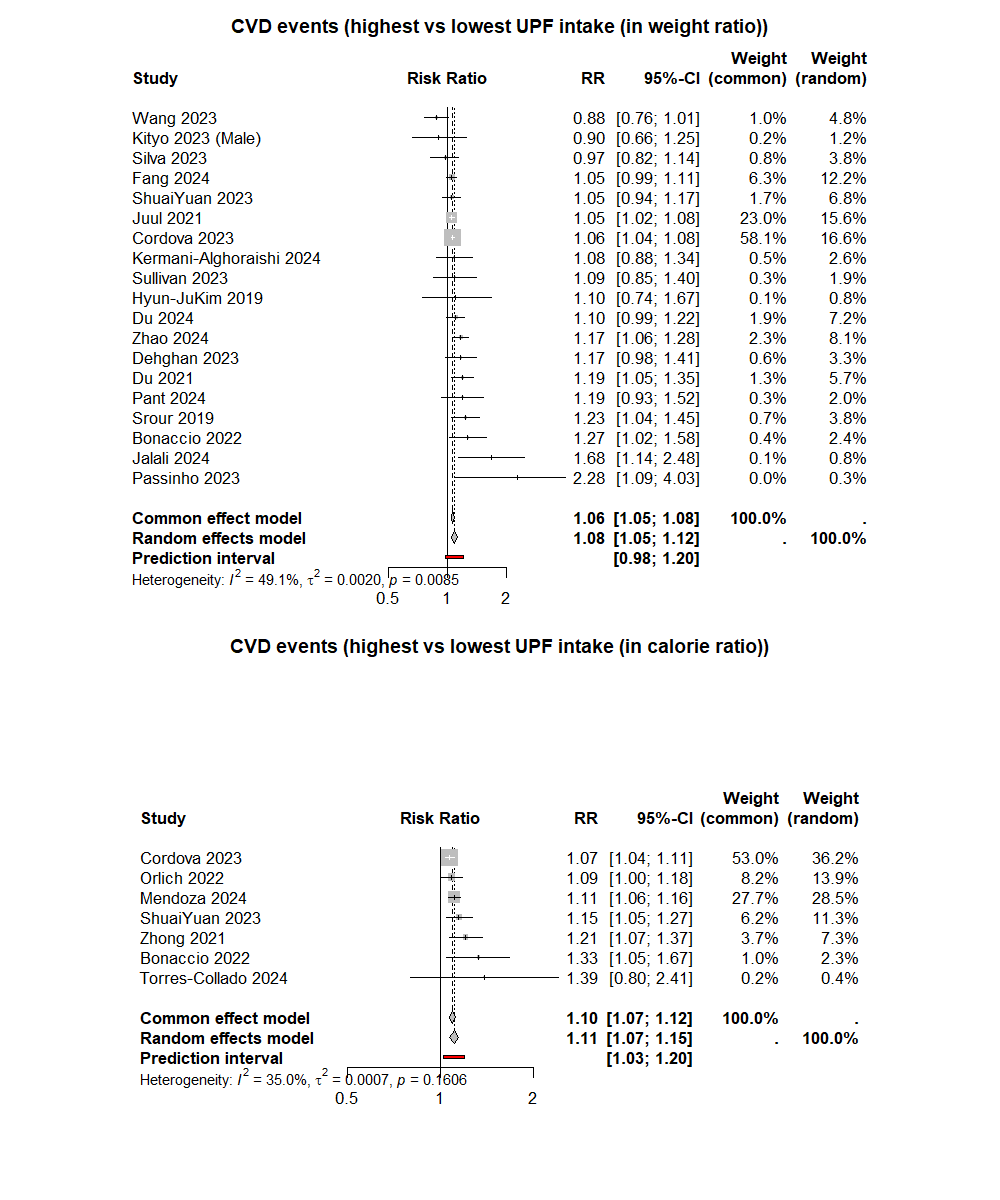

**Supplementary Figure 6.** Pooled effect estimate of ultra-processed food on cardiovascular disease (by quantile of energy and weight ratio)

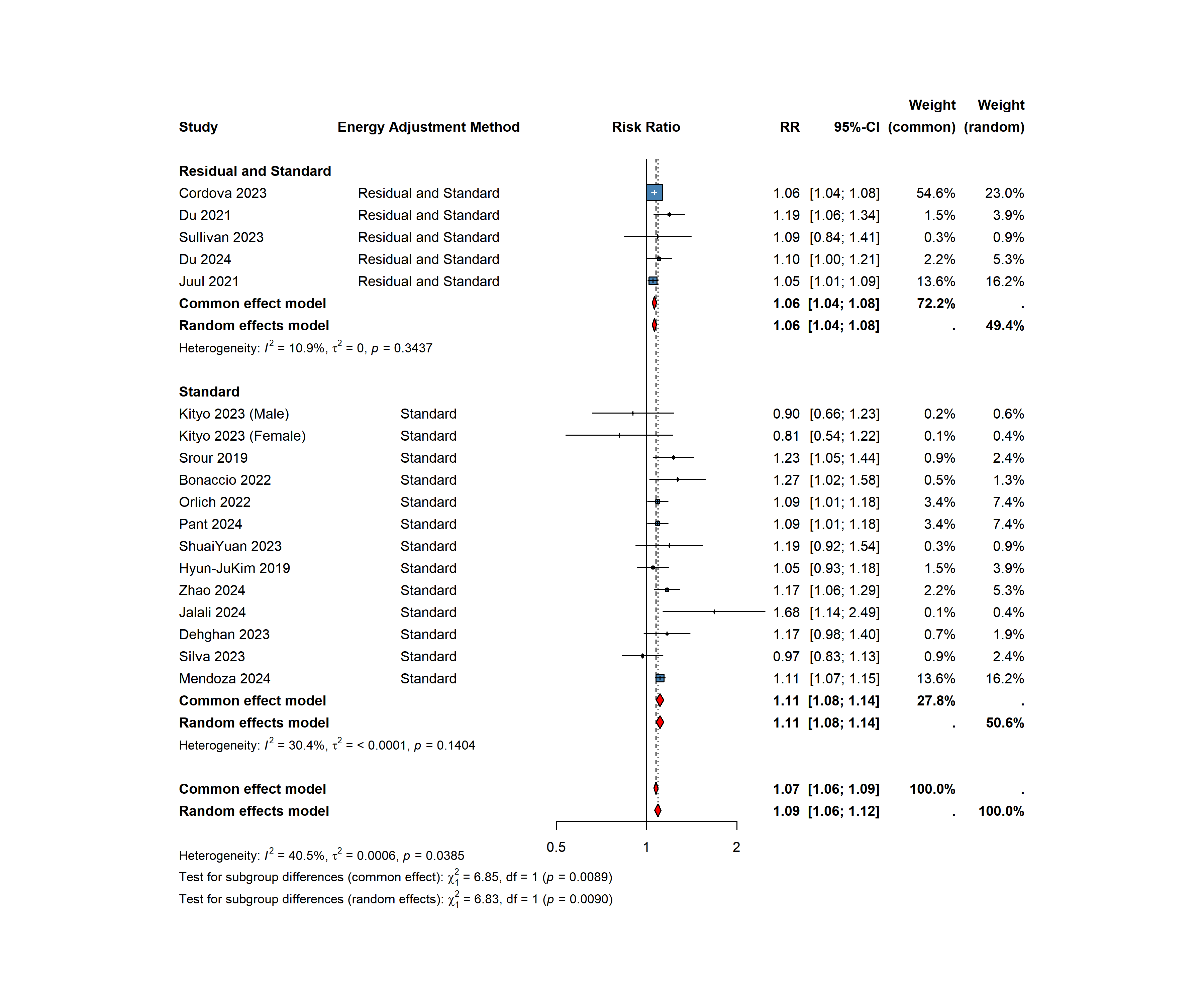

**Supplementary Figure 7**. Pooled effect estimates the association between ultra-processed food consumption and risk of cardiovascular disease outcome by total dietary energy adjustment methods

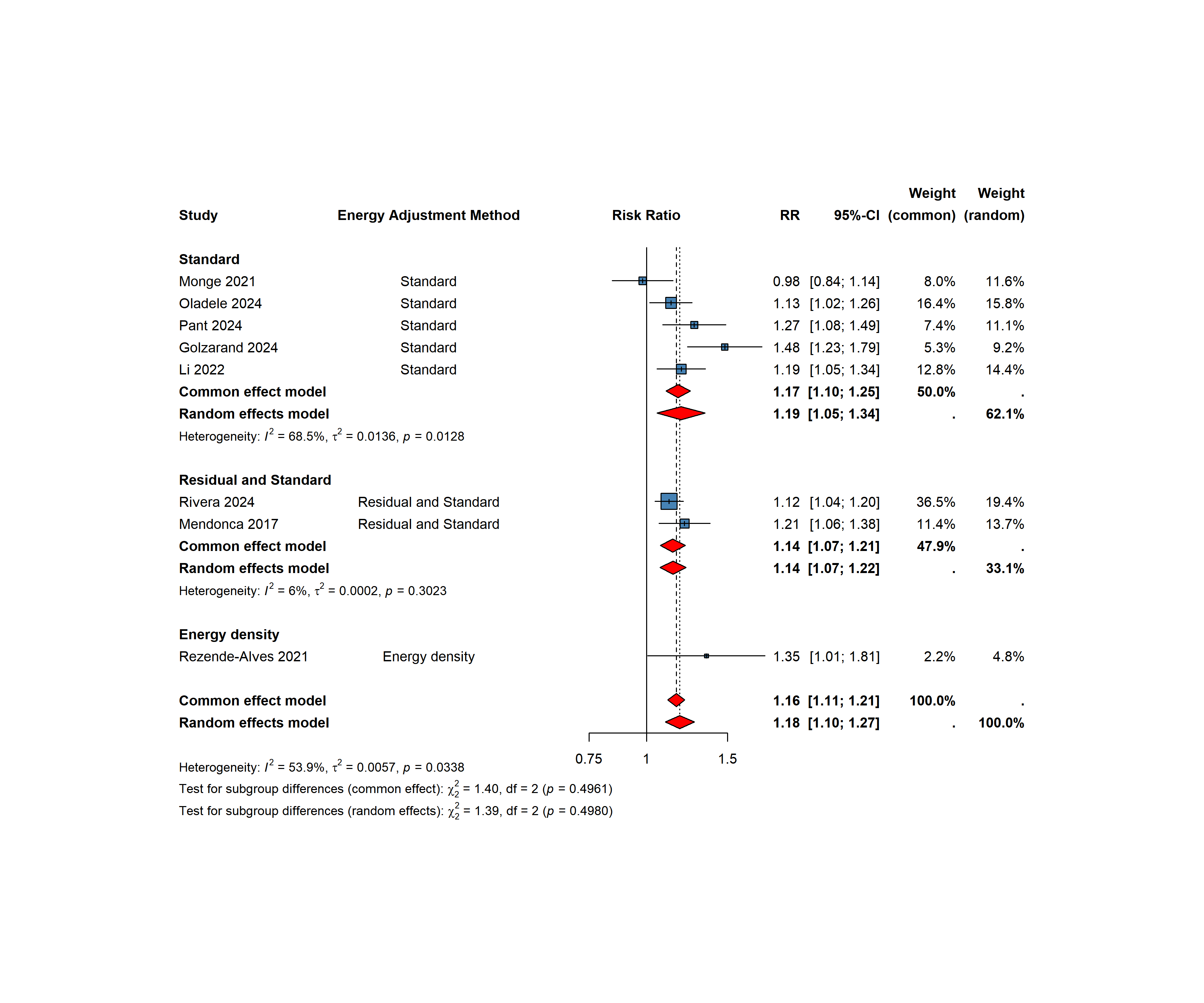

**Supplementary Figure 8**. Pooled effect estimates the association between ultra-processed food consumption and risk of hypertension by total dietary energy adjustment methods

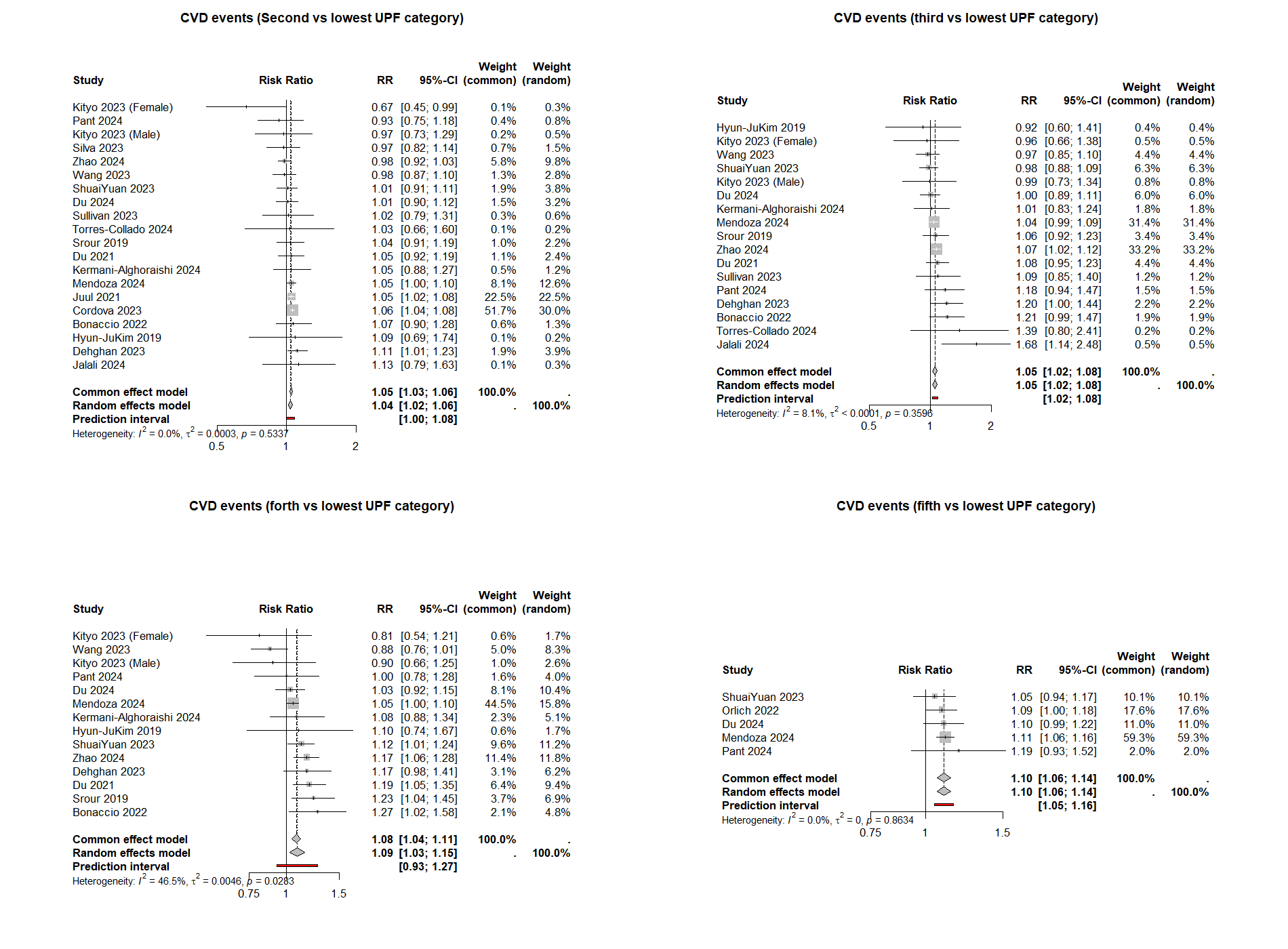

**Supplementary Figure 9**. Pooled effect estimate of ultra-processed food on cardiovascular disease by categories of ultra-processed food intake

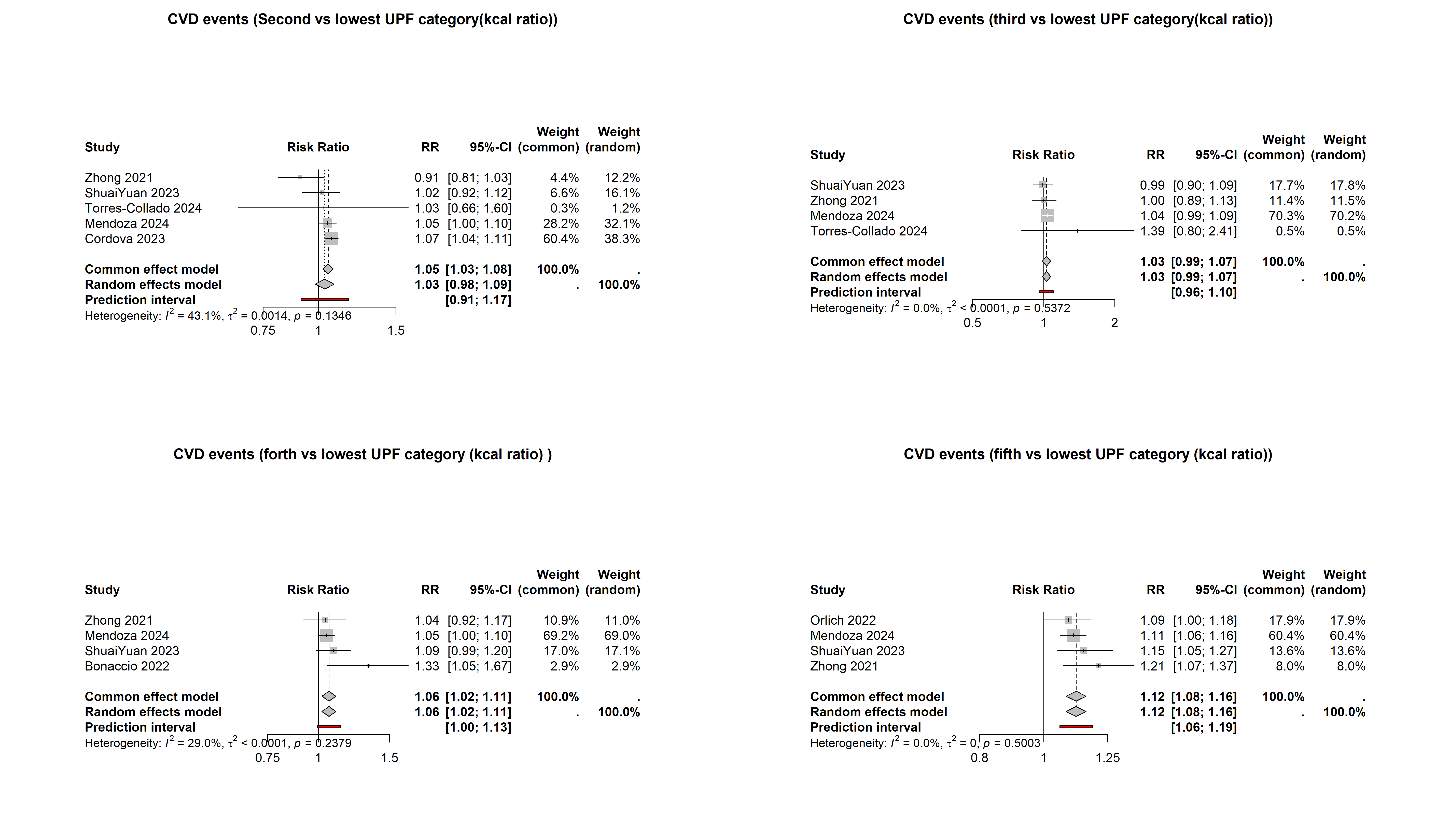

**Supplementary Figure 10**. Pooled effect estimate of ultra-processed food on cardiovascular disease by quantile of energy intake

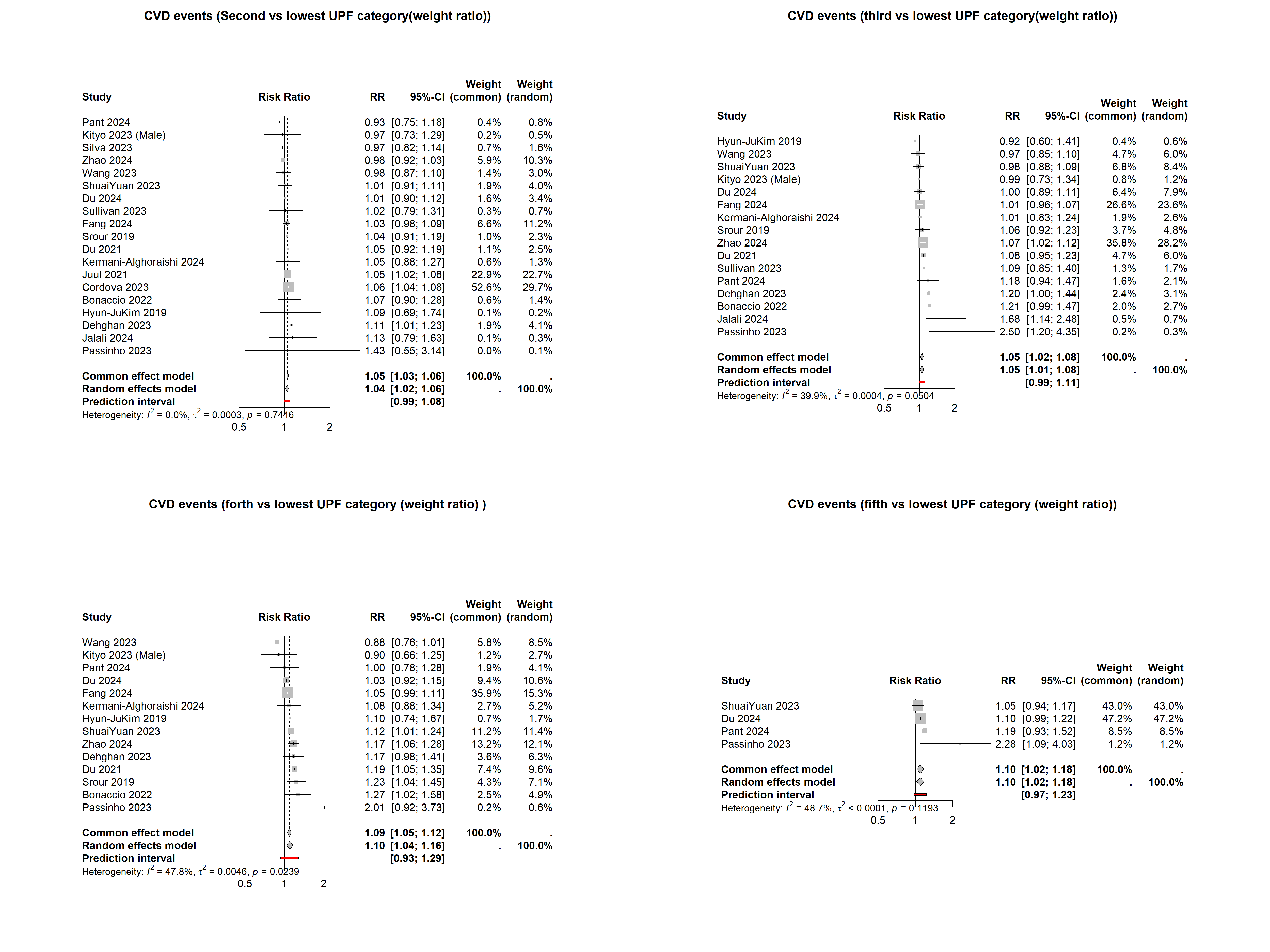

**Supplementary Figure 11**. Pooled effect estimate of ultra-processed food on cardiovascular disease by quantile of weight

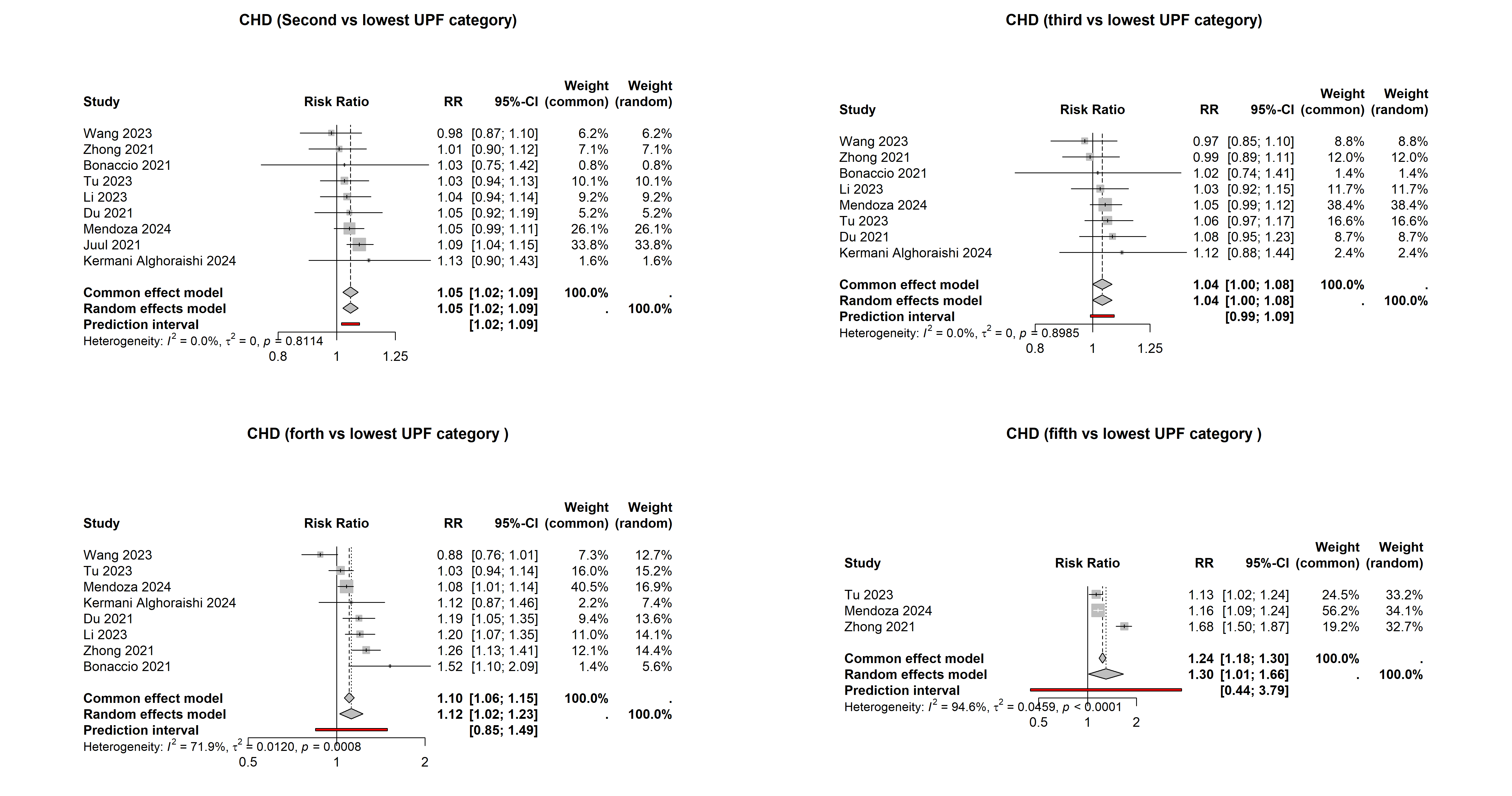

**Supplementary Figure 12**. Pooled effect estimate of ultra-processed food on coronary heart disease (quantile of both energy and weight intake)

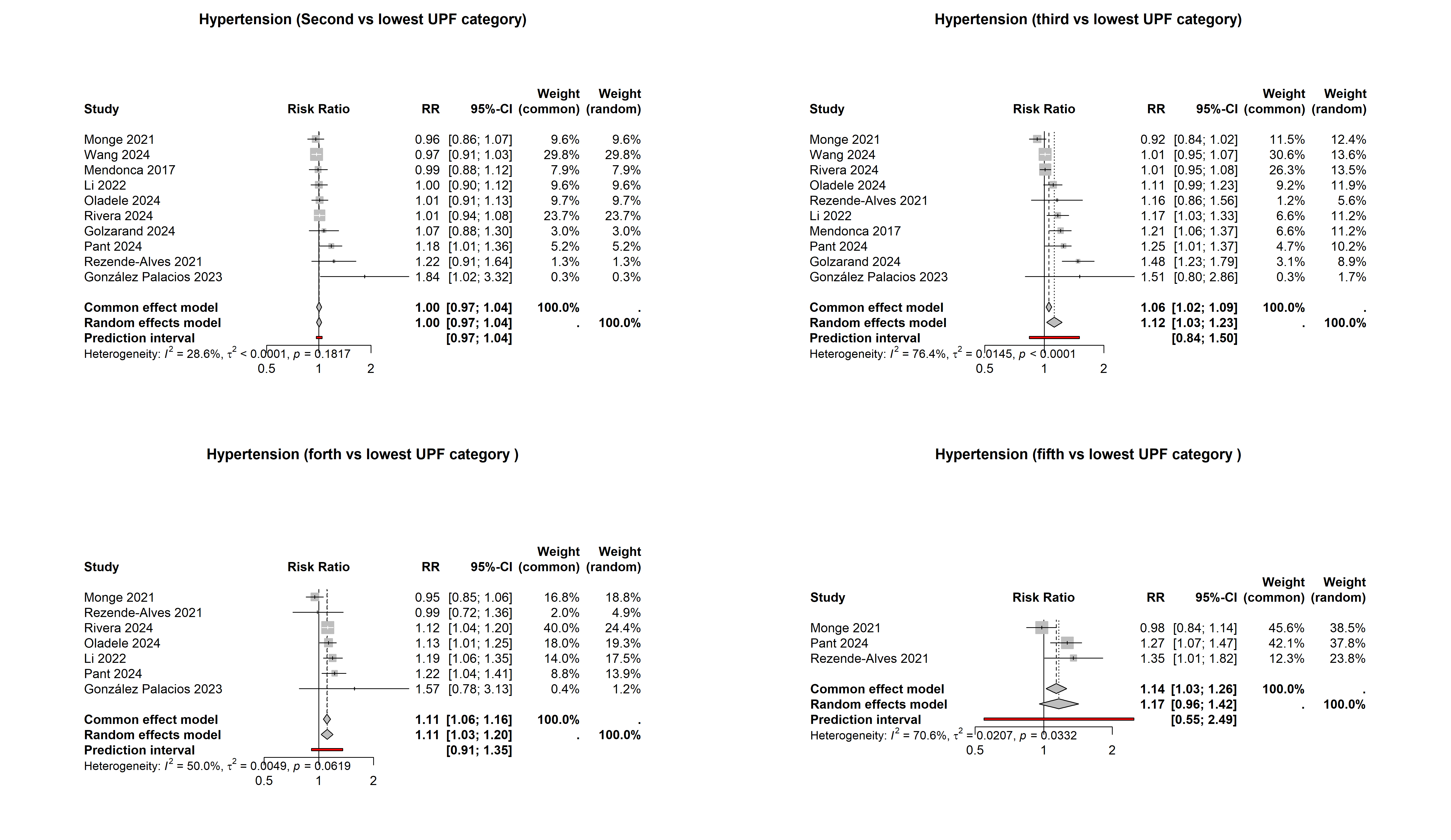

**Supplementary Figure 13**. Pooled effect estimate of ultra-processed food on hypertension (quantile of both energy and weight intake)

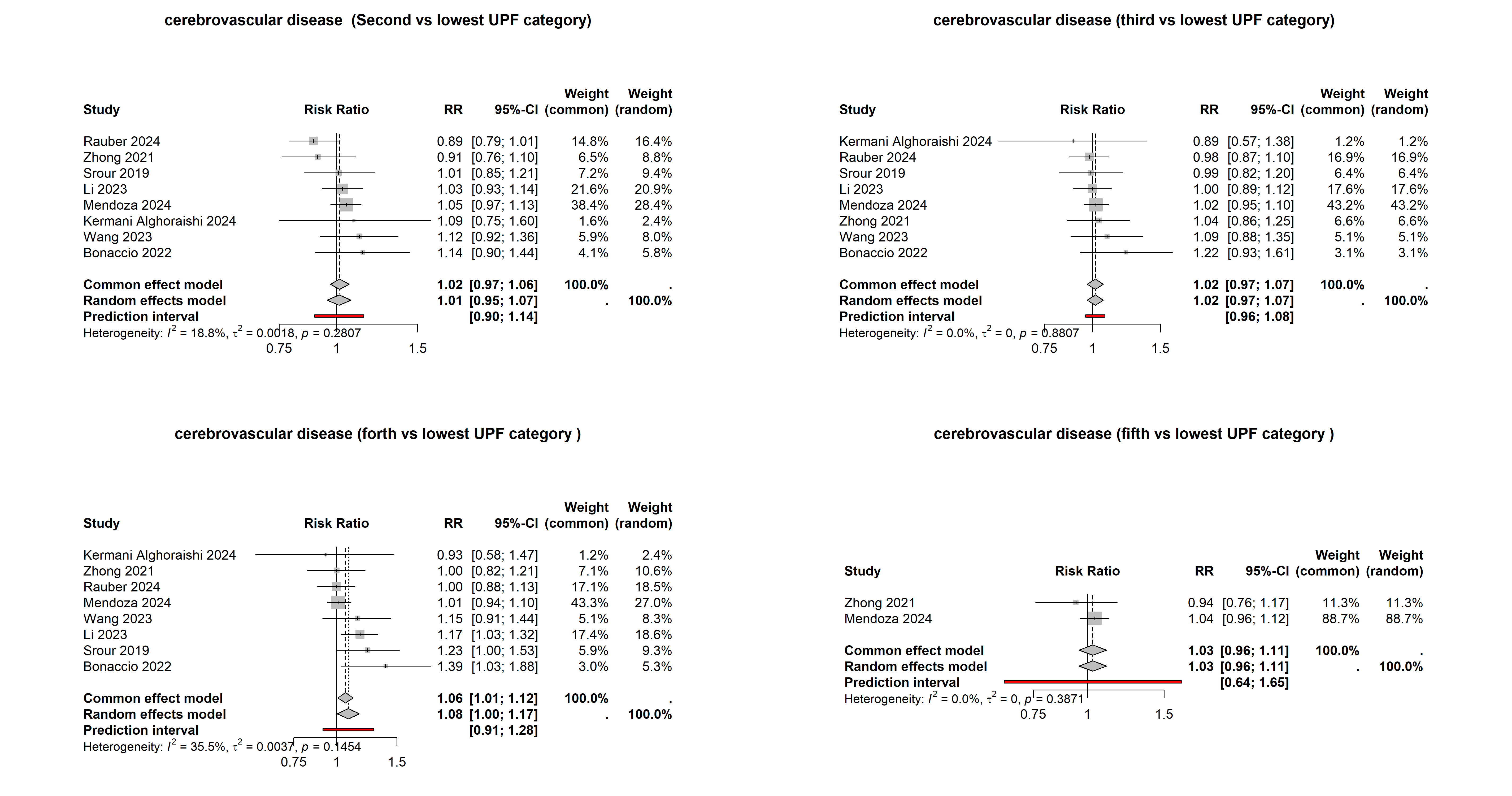

**Supplementary Figure 14**. Pooled effect estimate of ultra-processed food on cerebrovascular disease (quantile of both energy and weight intake

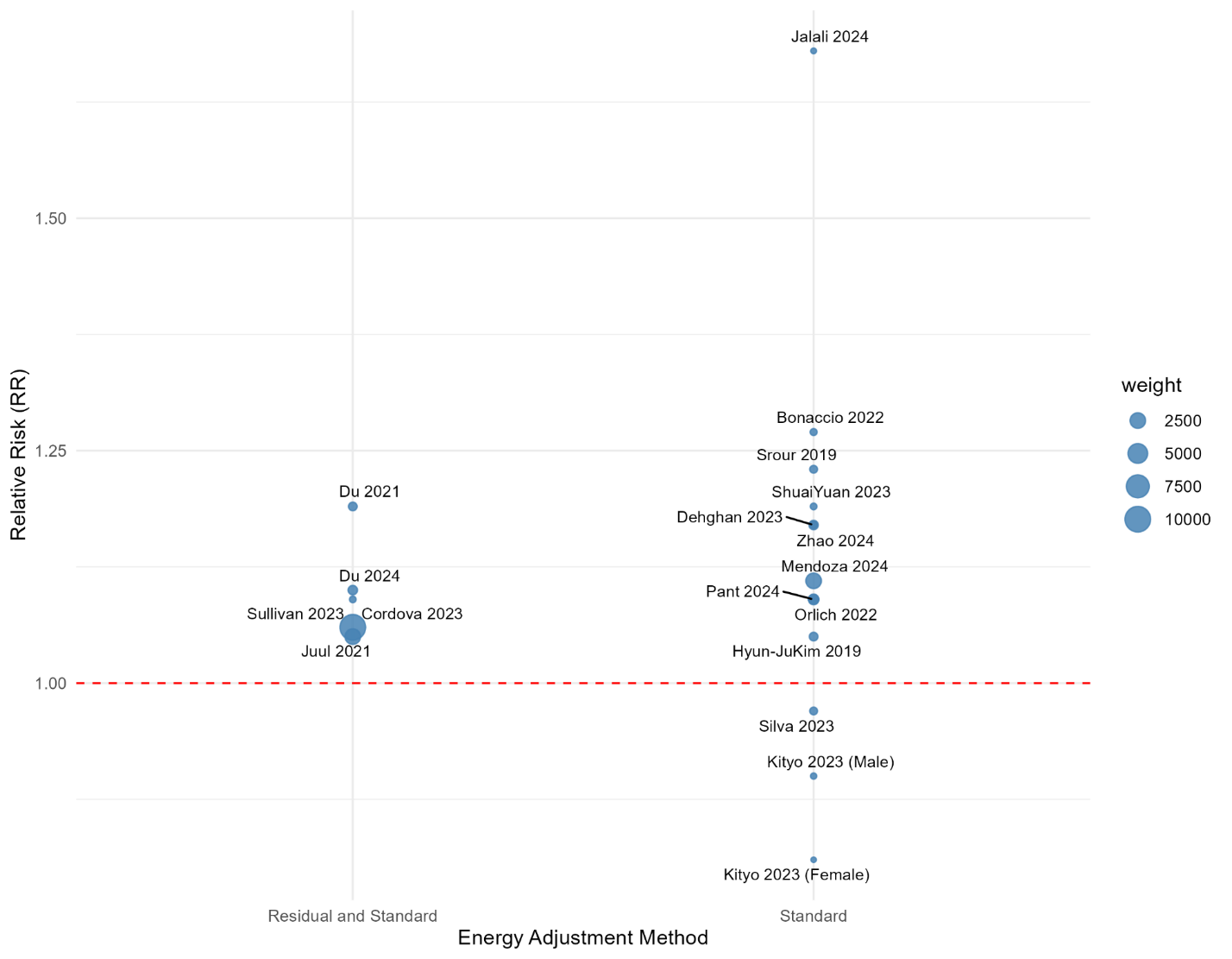

**Supplementary Figure 15**. Bubble plot from meta-regression examining the association between ultra-processed food consumption and cardiovascular disease events stratified by energy adjustment method. Each bubble represents an individual study, with bubble size proportional to the precision of the effect estimate (inverse of the variance). The vertical axis displays the effect size (RR) for the association between ultra-processed food consumption and cardiovascular disease events, and the horizontal axis represents the type of energy adjustment methods. The red line represents the null value. Heterogeneity in effect estimates across studies may partly reflect methodological differences in how energy intake was accounted for in the primary analyses

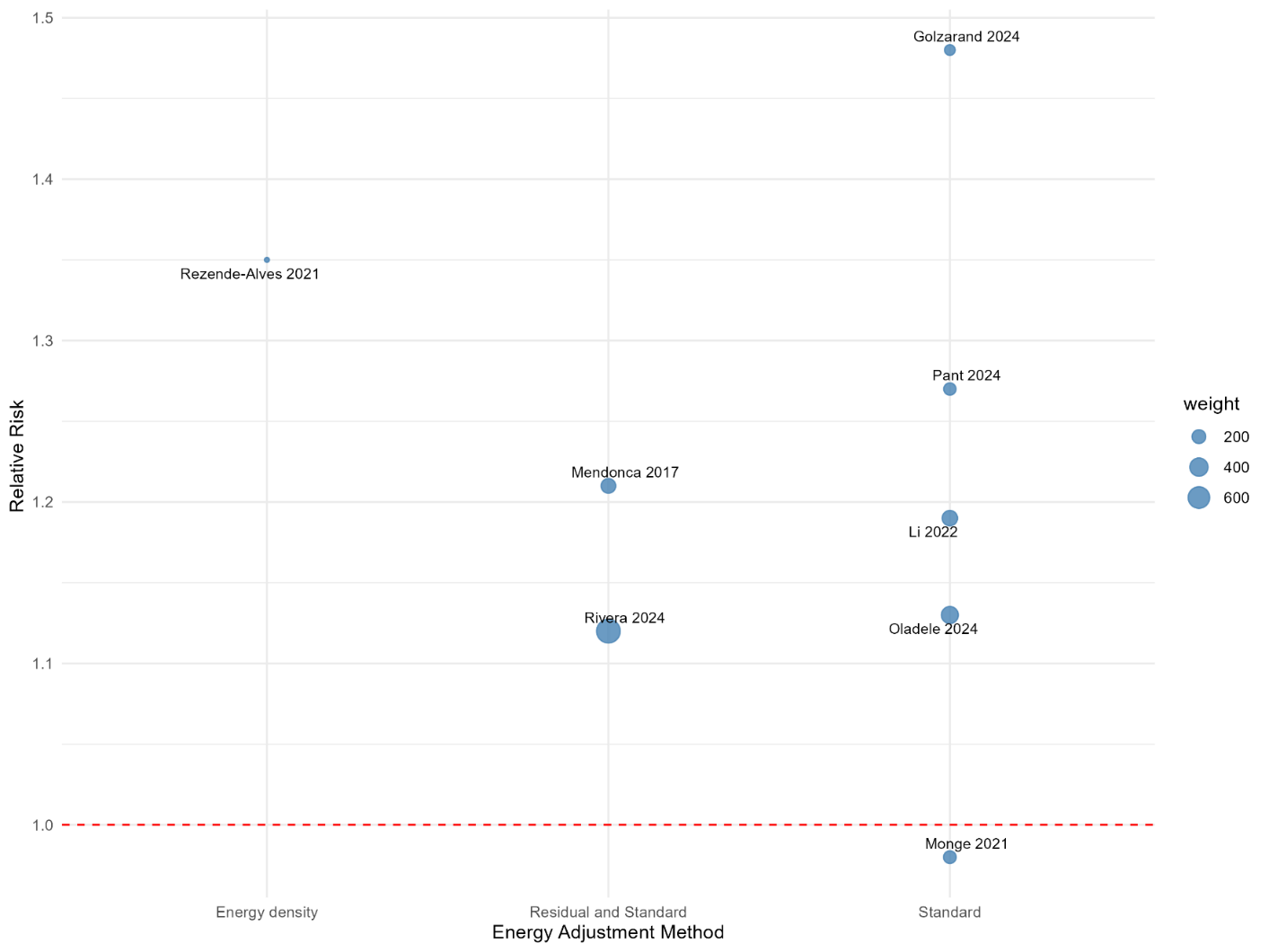

**Supplementary Figure 16**. Bubble plot from meta-regression examining the association between ultra-processed food consumption hypertension, stratified by energy adjustment method. Each bubble represents an individual study, with bubble size proportional to the precision of the effect estimate (inverse of the variance). The vertical axis displays the effect size (RR) for the association between ultra-processed food consumption and cardiovascular disease events, and the horizontal axis represents the type of energy adjustment methods. The red line represents the null value. Heterogeneity in effect estimates across studies may partly reflect methodological differences in how energy intake was accounted for in the primary analyses.

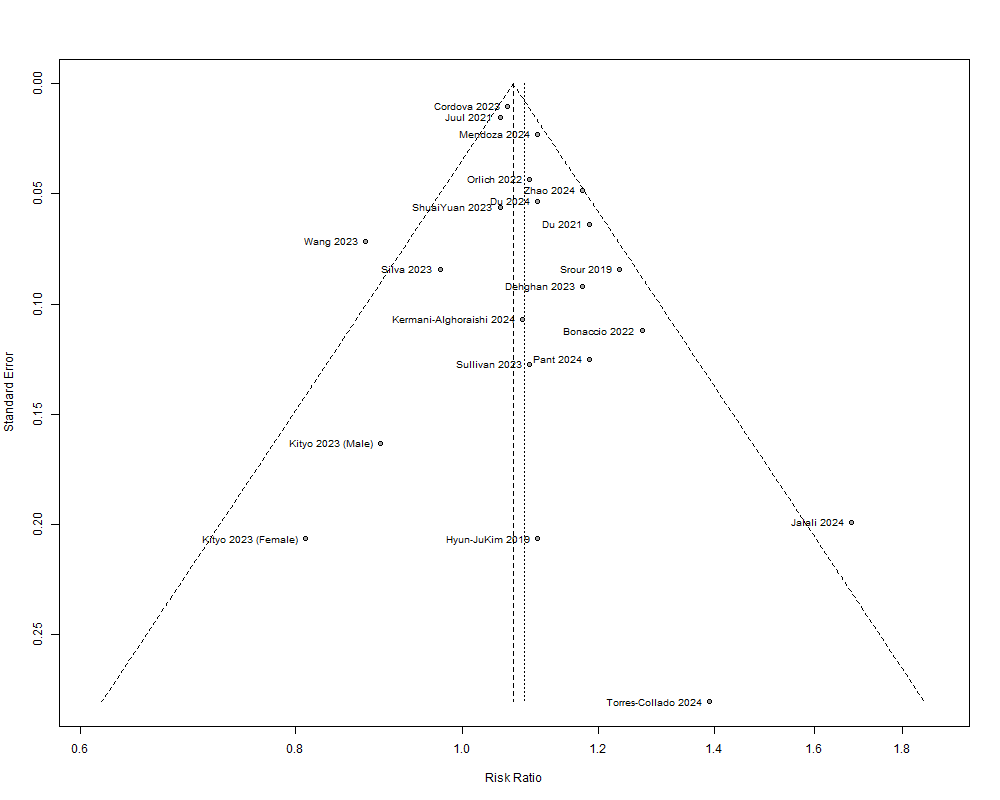

**Supplementary Figure 17.** Funnel plot (cardiovascular disease)

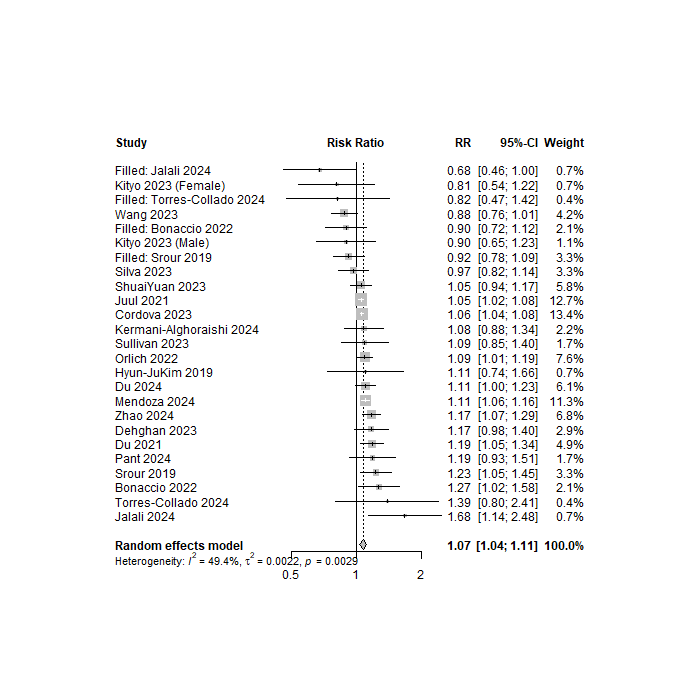

**Supplementary Figure 18**. Trim-and-fill analysis

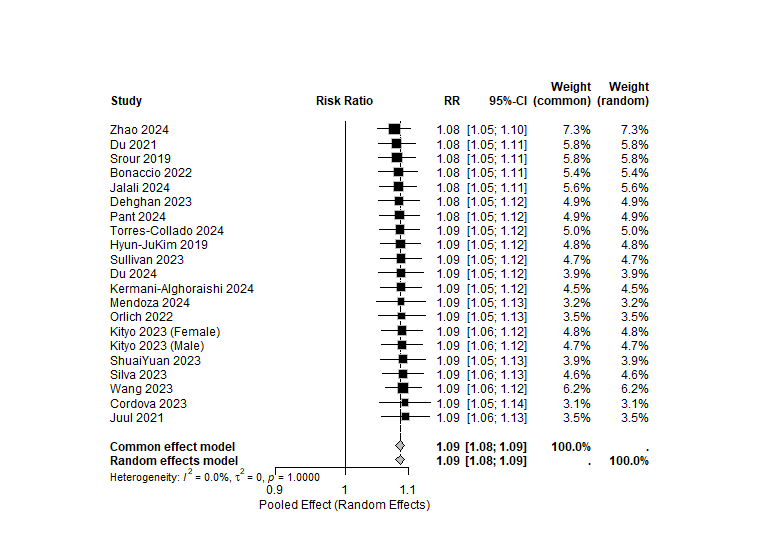

**Supplementary Figure 19**. Leave-one-out analysis

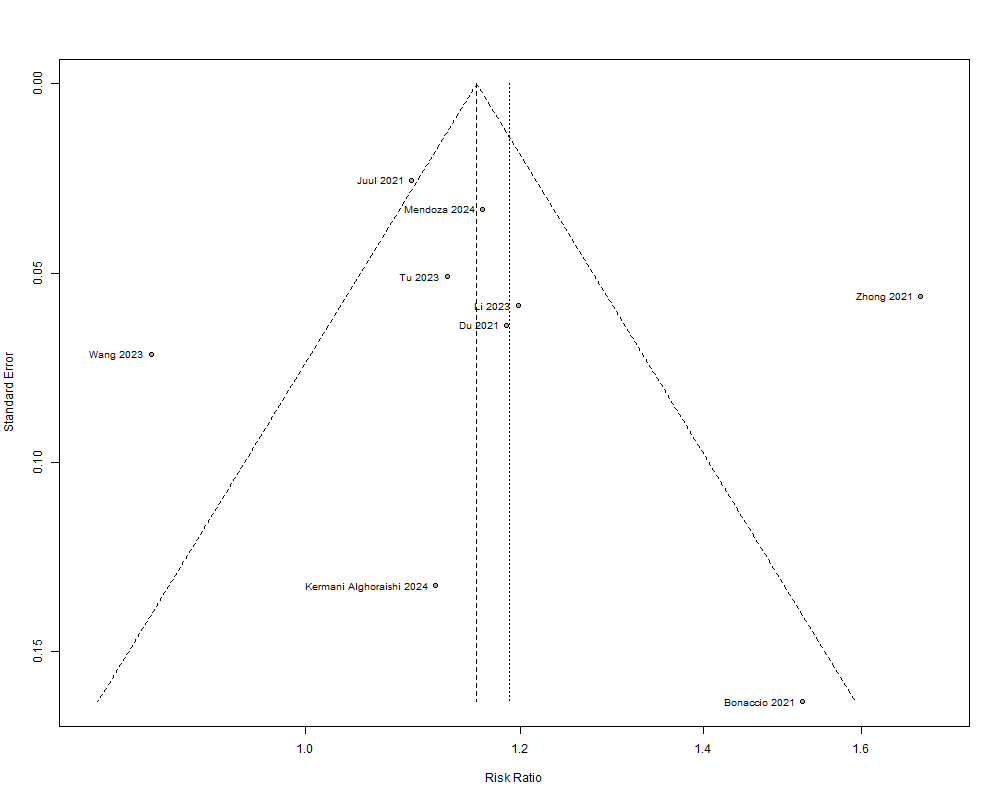

**Supplementary Figure 20.** Funnel plot (coronary heart disease)

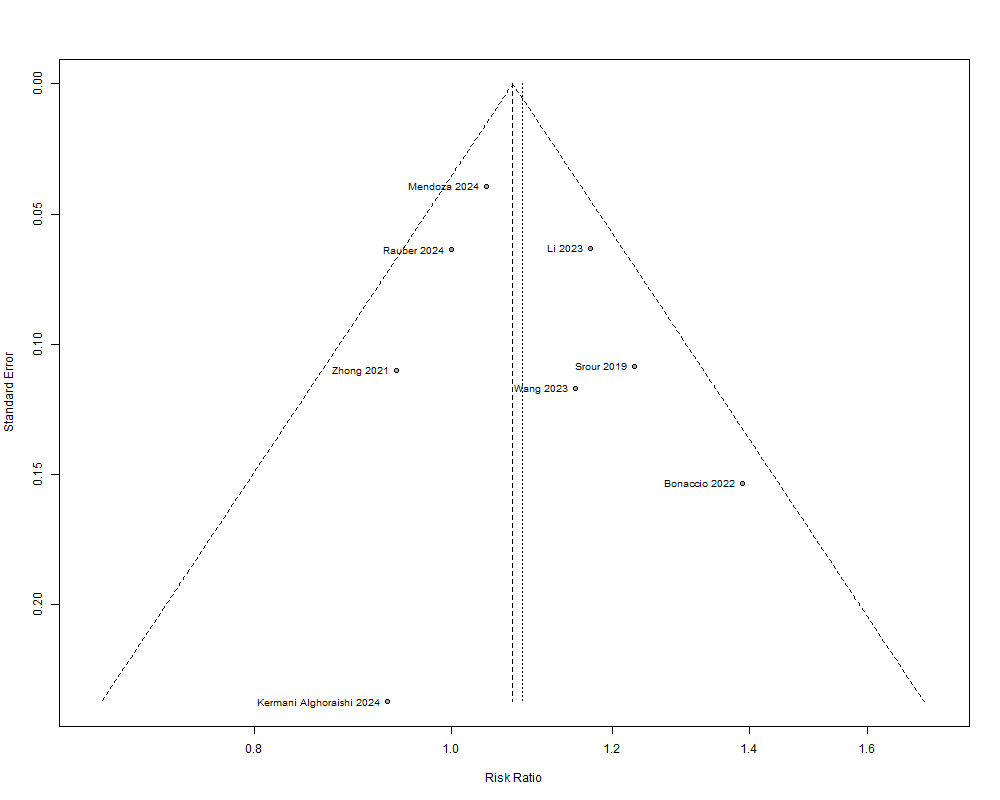

**Supplementary Figure 21.** Funnel plot (cerebrovascular disease)

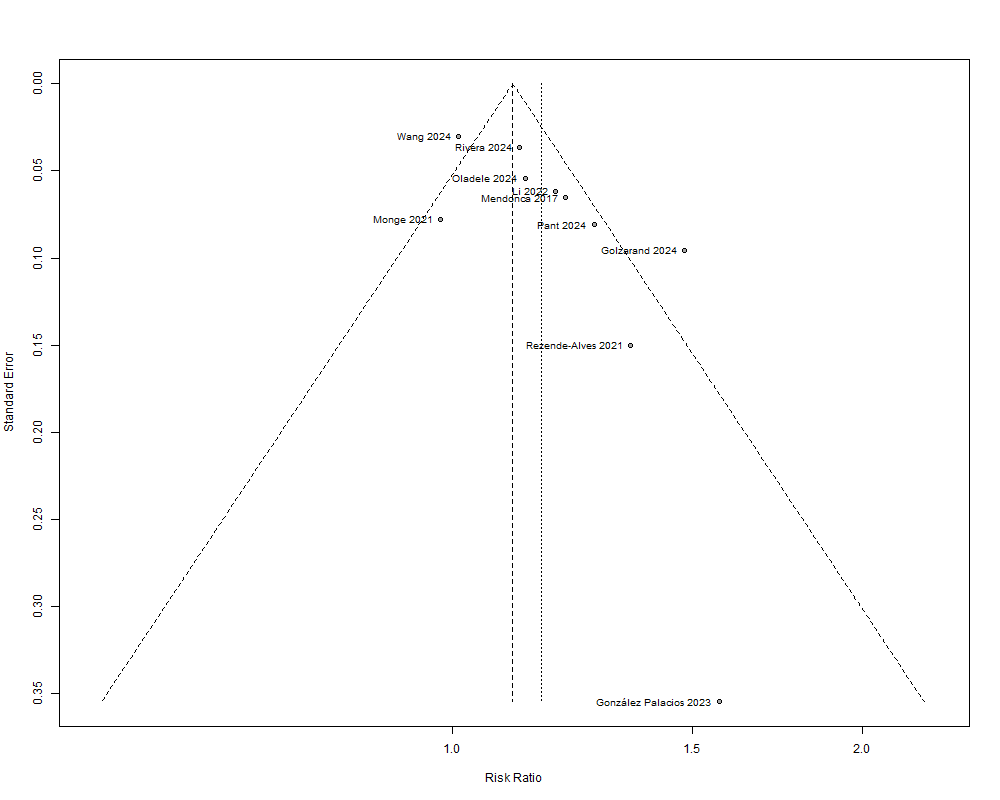

**Supplementary Figure 22.** Funnel plot (hypertension)

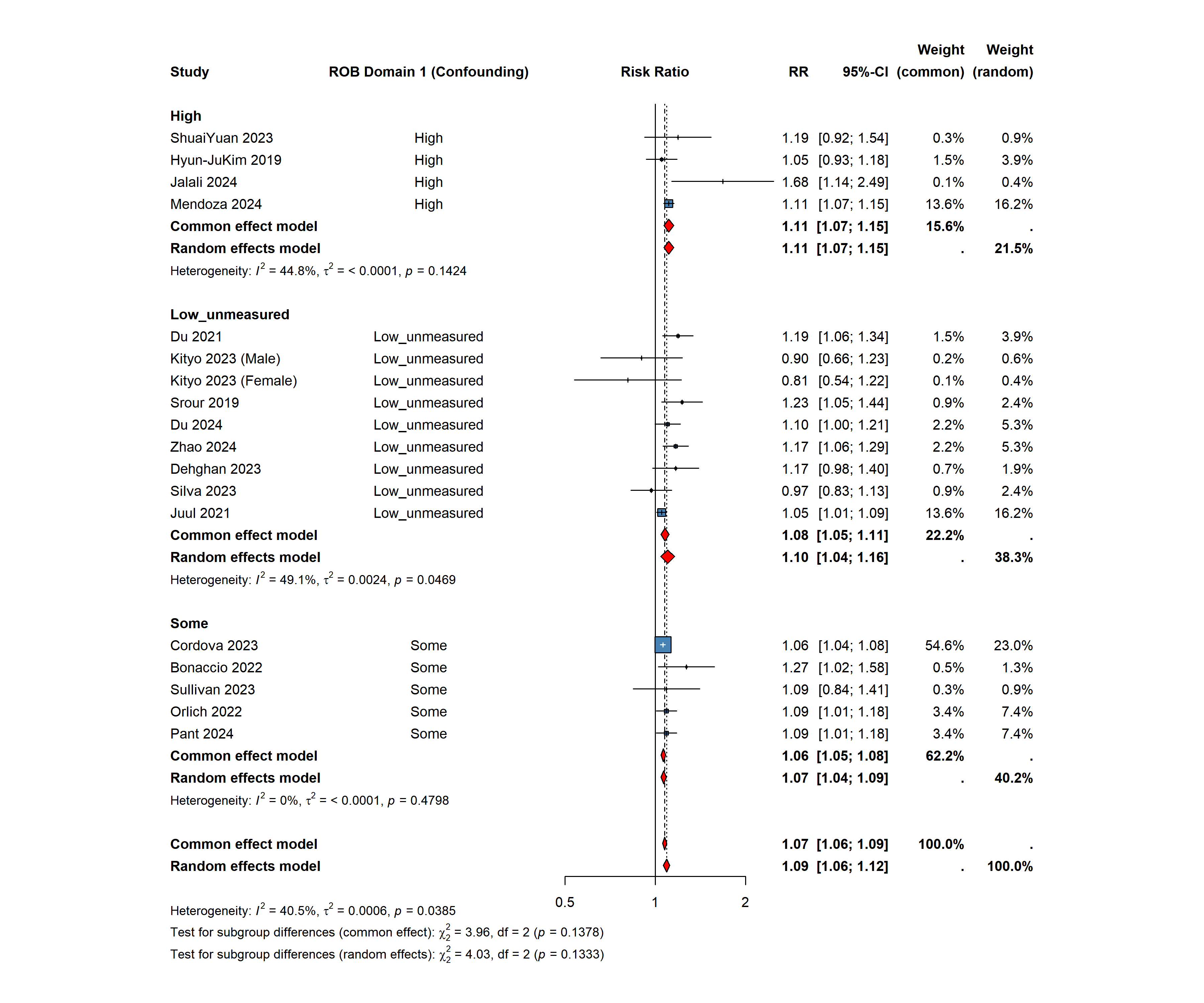

**Supplementary Figure 23.** Pooled effect estimates of ultra-processed food consumptions and risk of cardiovascular disease outcomes by risk of bias due to confounding

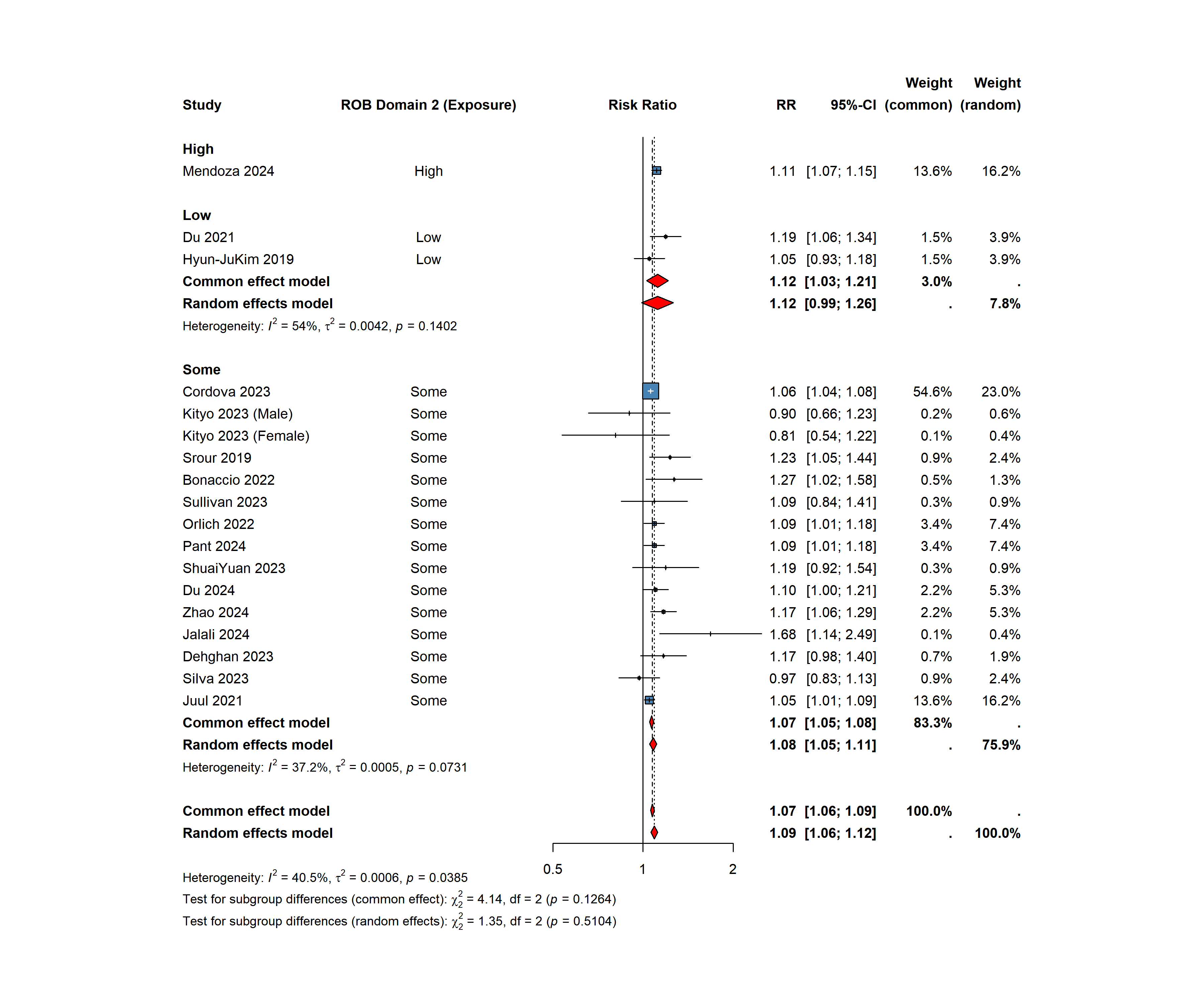

**Supplementary Figure 24.** Pooled effect estimates of ultra-processed food consumptions and risk of cardiovascular disease outcomes by risk of bias due to exposure measurement

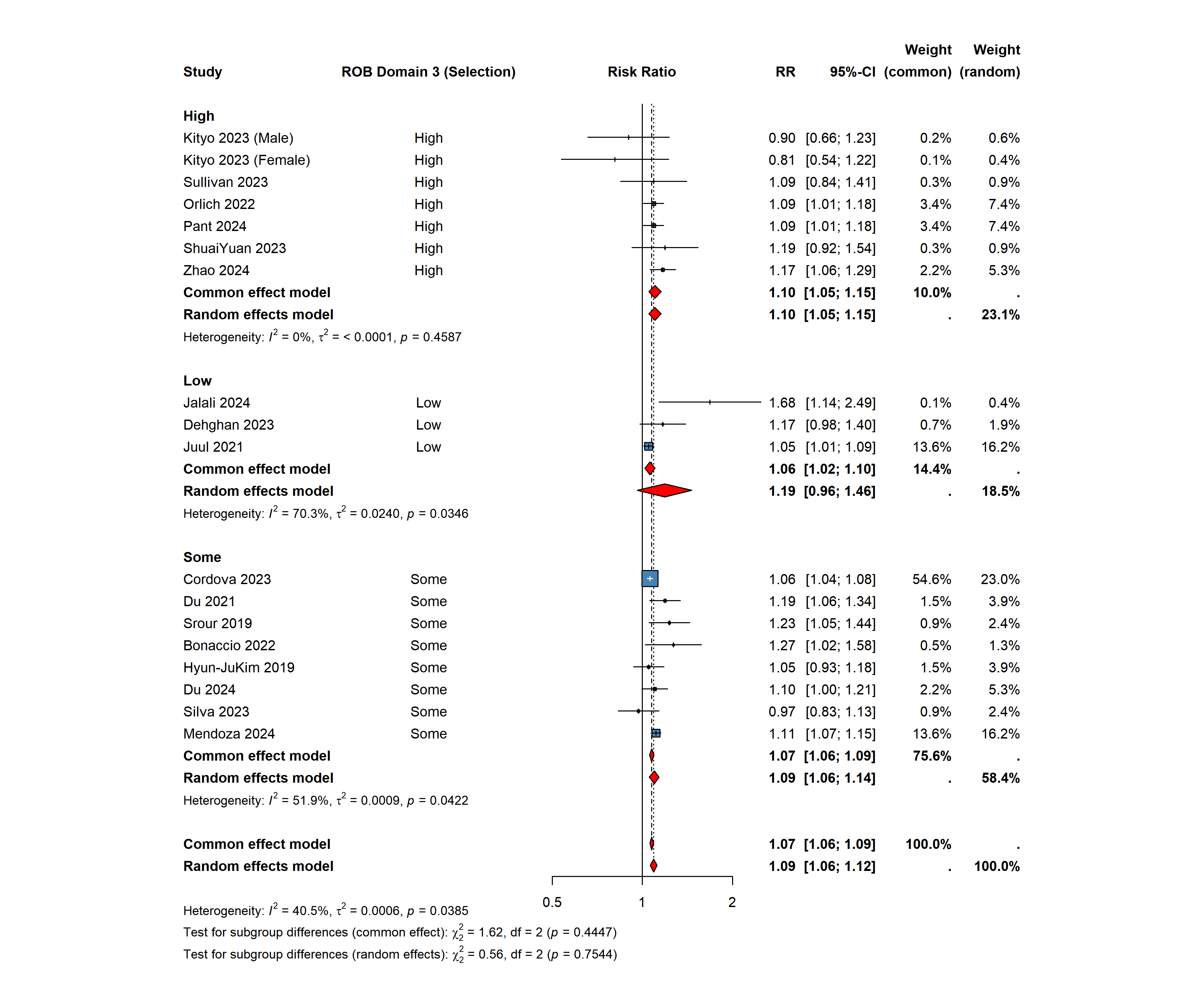

**Supplementary Figure 25.** Pooled effect estimates of ultra-processed food consumptions and risk of cardiovascular disease outcomes by risk of bias due to selection of participants

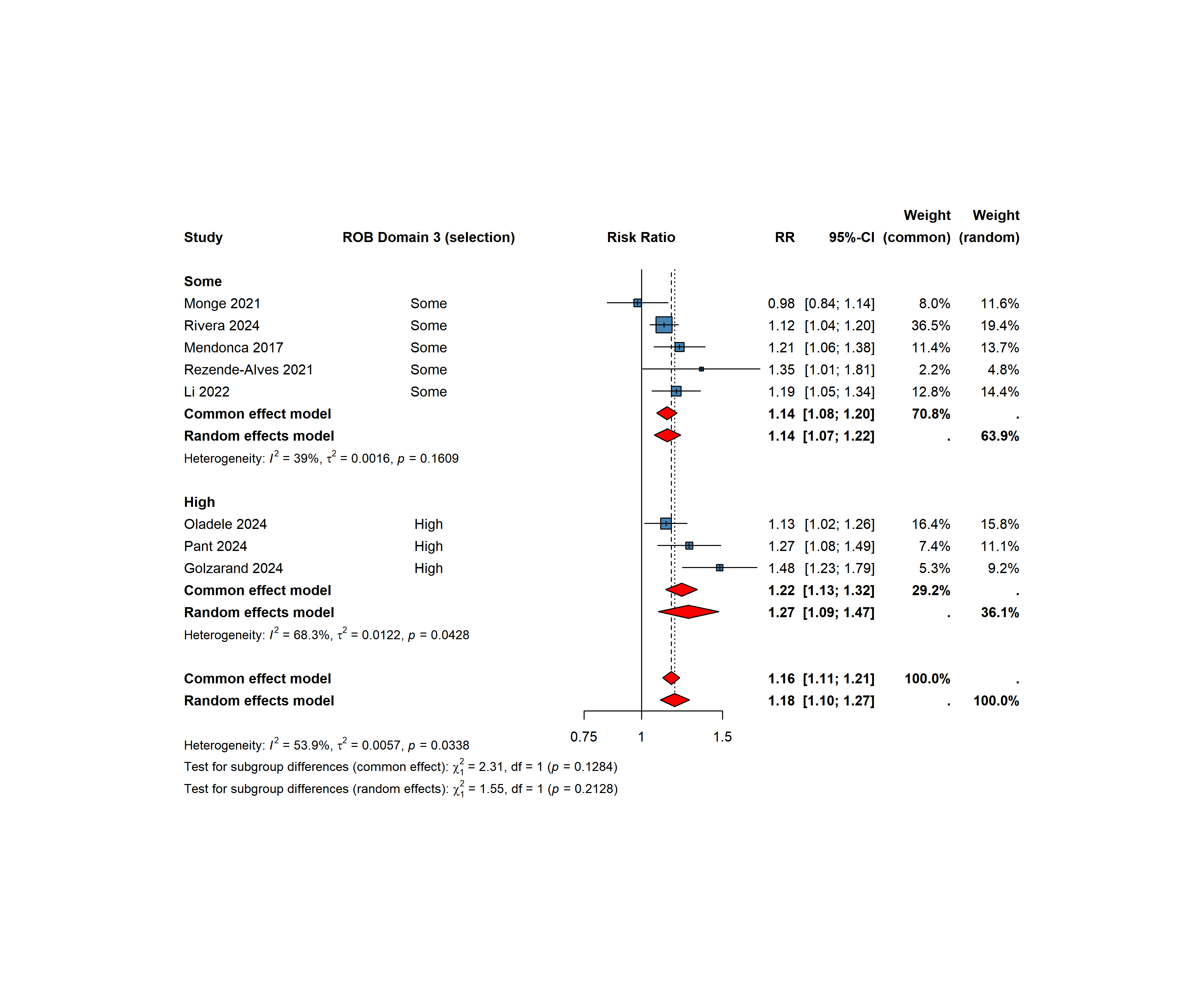

**Supplementary Figure 26.** Pooled effect estimates of ultra-processed food consumptions and risk of hypertension by risk of bias due to selection of participants

\# =============================================================================

\# SUPPLEMENTARY R SCRIPT

### # Title: A systematic review to critically appraise methodological rigour in research on ultra-processed food and #cardiovascular disease and hypertension

\# Journal: Critical Review in Food Science and Nutrition

\# Date: May 2026

\# =============================================================================

\# DESCRIPTION:

\#	This script reproduces all meta-analyses, forest plots, funnel plots,

\#	publication bias tests, leave-one-out sensitivity analyses, and

\#	meta-regression analyses reported in the main manuscript and

\#	supplementary materials.

\#

\# #

\# SOFTWARE:

\#	R version >= 4.2.0

\#	Key packages: meta (>= 6.0), ggplot2, gridExtra, grid, officer, flextable

\# =============================================================================

\# -----------------------------------------------------------------------------

\# SECTION 0: SETUP — Load Libraries & Global Settings

\# -----------------------------------------------------------------------------

library(meta)	# Meta-analysis

library(ggplot2)	# Plotting

library(gridExtra)	# Arranging multiple plots

library(grid)	# Grid graphics

library(gridGraphics)# Grid-based graphics capture

library(tidyverse)	# Data wrangling

library(broom)	# Tidy model outputs

library(flextable)	# Table formatting

library(officer)	# Word document export

\# Set global meta-analysis display options

settings.meta(digits = 2)

\# Set seed for reproducibility

set.seed(1234)

\# =============================================================================

\# SECTION 1: CVD EVENTS — HIGHEST vs LOWEST UPF INTAKE

\# (Regardless of unit of analysis; excluding hypertension; no duplicates)

\# =============================================================================

\# Load dataset

upf_cvd <- UPF_CVD_Regardless_UA_Nodup_No_HTN

\# -----------------------------------------------------------------------------

\# 1.1 Primary Meta-Analysis (Highest vs Lowest UPF Intake)

\# -----------------------------------------------------------------------------

Fplot <- metagen(

TE	= h_L_logrr,

lower	= LB_h_l,

upper	= UB_h_l,

studlab = Author_Name_Year,

data	= upf_cvd,

sm	= "RR",

method.tau = "REML"

)

summary(Fplot)

\# Forest plot — random effects, sorted by effect size

png("forest_plot_CVD_highest_vs_lowest.png",

width = 1000, height = 760, res = 100)

grid.arrange(

grid.grabExpr(

forest(Fplot,

sortvar	= TE,

random	= TRUE,

common	= TRUE,

prediction = TRUE,

digits	= 2,

leftcols	= "studlab")

),

top = textGrob(

"CVD events (highest vs lowest UPF intake)",

gp = gpar(fontsize = 14, fontface = "bold")

)

)

dev.off()

\# -----------------------------------------------------------------------------

\# 1.2 Dose–Response Analysis (CVD events per serving increment)

\# -----------------------------------------------------------------------------

\# Select studies with dose-response data

upf_cvd_dose <- UPF_CVD_Regardless_UA_Nodup[

c(1, 3, 6, 7, 9, 15, 17, 19, 21, 24, 25, 26), ]

upf_cvd_dose$Dose_res <- as.numeric(upf_cvd_dose$Dose_res)

upf_cvd_dose$LB_dose <- as.numeric(upf_cvd_dose$LB_dose)

upf_cvd_dose$UB_dose <- as.numeric(upf_cvd_dose$UB_dose)

Fplotdose <- metagen(

TE	= log(Dose_res),

lower	= log(LB_dose),

upper	= log(UB_dose),

studlab = Author_Name_Year,

data	= upf_cvd_dose,

sm	= "RR",

method.tau = "REML"

)

png("forest_plot_CVD_dose_response.png", width = 800, height = 600)

forest(Fplotdose,

sortvar	= TE,

random	= TRUE,

common	= TRUE,

prediction = TRUE,

digits	= 2)

dev.off()

\# =============================================================================

\# SECTION 2: CVD EVENTS BY UPF INTAKE CATEGORY

\# (Regardless of unit of analysis; no duplicates)

\# =============================================================================

upf_cvd <- UPF_CVD_Regardless_UA_Nodup

\# -----------------------------------------------------------------------------

\# 2.1 Second vs Lowest UPF Category

\# -----------------------------------------------------------------------------

upf_cvd_cat2 <- upf_cvd[-10, ]	# Exclude row 10 (missing category 2 data)

Fplot_cat2 <- metagen(

TE	= log(effect1),

lower	= log(LB1),

upper	= log(UB1),

studlab = Author_Name_Year,

data	= upf_cvd_cat2,

sm	= "RR",

method.tau = "REML"

)

\# -----------------------------------------------------------------------------

\# 2.2 Third vs Lowest UPF Category

\# -----------------------------------------------------------------------------

upf_cvd_cat3 <- upf_cvd[-c(1, 10, 18, 21), ]	# Exclude rows missing cat 3

Fplot_cat3 <- metagen(

TE	= log(effect2),

lower	= log(LB2),

upper	= log(UB2),

studlab = Author_Name_Year,

data	= upf_cvd_cat3,

sm	= "RR",

method.tau = "REML"

)

\# -----------------------------------------------------------------------------

\# 2.3 Fourth vs Lowest UPF Category

\# -----------------------------------------------------------------------------

upf_cvd_cat4 <- upf_cvd[-c(1, 7, 9, 10, 16, 18, 21), ]

Fplot_cat4 <- metagen(

TE	= log(effect3),

lower	= log(LB3),

upper	= log(UB3),

studlab = Author_Name_Year,

data	= upf_cvd_cat4,

sm	= "RR",

method.tau = "REML"

)

\# -----------------------------------------------------------------------------

\# 2.4 Fifth vs Lowest UPF Category

\# -----------------------------------------------------------------------------

upf_cvd_cat5 <- upf_cvd[c(10, 11, 12, 14, 20), ]

Fplot_cat5 <- metagen(

TE	= log(effect4),

lower	= log(LB4),

upper	= log(UB4),

studlab = Author_Name_Year,

data	= upf_cvd_cat5,

sm	= "RR",

method.tau = "REML"

)

\# -----------------------------------------------------------------------------

\# 2.5 Combined Forest Plot — All CVD Categories (2^nd^–5^th^ vs Lowest)

\# -----------------------------------------------------------------------------

FP_cat2 <- grid.grabExpr(

forest(Fplot_cat2, sortvar = TE, random = TRUE, common = TRUE,

prediction = TRUE, digits = 2, leftcols = "studlab"))

FP_cat3 <- grid.grabExpr(

forest(Fplot_cat3, sortvar = TE, random = TRUE, common = TRUE,

prediction = TRUE, digits = 2, leftcols = "studlab"))

FP_cat4 <- grid.grabExpr(

forest(Fplot_cat4, sortvar = TE, random = TRUE, common = TRUE,

prediction = TRUE, digits = 2, leftcols = "studlab"))

FP_cat5 <- grid.grabExpr(

forest(Fplot_cat5, sortvar = TE, random = TRUE, common = TRUE,

prediction = TRUE, digits = 2, leftcols = "studlab"))

FP_cat2 <- arrangeGrob(FP_cat2, top = textGrob("CVD events (2nd vs lowest UPF category)",

gp = gpar(fontsize = 14, fontface = "bold"), just = "top"), padding = unit(5, "mm"))

FP_cat3 <- arrangeGrob(FP_cat3, top = textGrob("CVD events (3rd vs lowest UPF category)",

gp = gpar(fontsize = 14, fontface = "bold"), just = "top"), padding = unit(5, "mm"))

FP_cat4 <- arrangeGrob(FP_cat4, top = textGrob("CVD events (4th vs lowest UPF category)",

gp = gpar(fontsize = 14, fontface = "bold"), just = "top"), padding = unit(15, "mm"))

FP_cat5 <- arrangeGrob(FP_cat5, top = textGrob("CVD events (5th vs lowest UPF category)",

gp = gpar(fontsize = 14, fontface = "bold"), just = "top"), padding = unit(15, "mm"))

png("ForestPlots_CVD_UPF_Categories.png",

width = 19, height = 14, units = "in", res = 100)

grid.arrange(FP_cat2, FP_cat3, FP_cat4, FP_cat5, ncol = 2)

dev.off()

\# =============================================================================

\# SECTION 3: CVD EVENTS BY UPF INTAKE (WEIGHT RATIO)

\# =============================================================================

weight <- UPF_CVD_weight

\# -----------------------------------------------------------------------------

\# 3.1 Highest vs Lowest (Weight Ratio)

\# -----------------------------------------------------------------------------

Fplotwt_hl <- metagen(

TE	= logrr_h_l,

lower	= Lb_h_l,

upper	= Ub_h_l,

studlab = Author_Name_Year,

data	= UPF_CVD_NO_HTN_weight,

sm	= "RR",

method.tau = "REML"

)

png("forest_plot_CVD_weight_highest_vs_lowest.png", width = 800, height = 600)

forest(Fplotwt_hl, sortvar = TE, random = TRUE, common = TRUE,

prediction = TRUE, digits = 2, leftcols = "studlab")

dev.off()

\# -----------------------------------------------------------------------------

\# 3.2 Categories 2–5 vs Lowest (Weight Ratio)

\# -----------------------------------------------------------------------------

\# Second category

Fplotwt_cat2 <- metagen(

TE = log(effect1), lower = log(LB1), upper = log(UB1),

studlab = Author_Name_Year, data = weight, sm = "RR", method.tau = "REML"

)

\# Third category (exclude rows 1, 17, 19)

wt_cat3 <- weight[-c(1, 17, 19), ]

Fplotwt_cat3 <- metagen(

TE = log(effect2), lower = log(LB2), upper = log(UB2),

studlab = Author_Name_Year, data = wt_cat3, sm = "RR", method.tau = "REML"

)

\# Fourth category (exclude rows 1, 8, 15, 17, 19)

wt_cat4 <- weight[-c(1, 8, 15, 17, 19), ]

Fplotwt_cat4 <- metagen(

TE = log(effect3), lower = log(LB3), upper = log(UB3),

studlab = Author_Name_Year, data = wt_cat4, sm = "RR", method.tau = "REML"

)

\# Fifth category (select rows 9, 10, 11, 13)

wt_cat5 <- weight[c(9, 10, 11, 13), ]

Fplotwt_cat5 <- metagen(

TE = log(effect4), lower = log(LB4), upper = log(UB4),

studlab = Author_Name_Year, data = wt_cat5, sm = "RR", method.tau = "REML"

)

\# -----------------------------------------------------------------------------

\# 3.3 Dose–Response (Weight Ratio)

\# -----------------------------------------------------------------------------

wt_dose <- weight[c(1, 3, 5, 6, 8, 14, 16, 18, 20, 22, 23, 24), ]

wt_dose$logrr_doae <- as.numeric(wt_dose$logrr_doae)

wt_dose$lb_dose	<- as.numeric(wt_dose$lb_dose)

wt_dose$ub_dose	<- as.numeric(wt_dose$ub_dose)

Fplotwt_dose <- metagen(

TE = logrr_doae, lower = lb_dose, upper = ub_dose,

studlab = Author_Name_Year, data = wt_dose, sm = "RR", method.tau = "REML"

)

png("forest_plot_CVD_weight_dose_response.png", width = 700, height = 450)

forest(Fplotwt_dose, sortvar = TE, random = TRUE, common = TRUE,

prediction = TRUE, digits = 2)

dev.off()

\# -----------------------------------------------------------------------------

\# 3.4 Combined Forest Plot — All Weight-Ratio Categories

\# -----------------------------------------------------------------------------

FPwt2 <- grid.grabExpr(forest(Fplotwt_cat2, sortvar = TE, backtransf = TRUE, digits = 2))

FPwt3 <- grid.grabExpr(forest(Fplotwt_cat3, sortvar = TE, backtransf = TRUE, digits = 2))

FPwt4 <- grid.grabExpr(forest(Fplotwt_cat4, sortvar = TE, backtransf = TRUE, digits = 2))

FPwt5 <- grid.grabExpr(forest(Fplotwt_cat5, sortvar = TE, backtransf = TRUE, digits = 2))

FPwt2 <- arrangeGrob(FPwt2, top = textGrob("CVD events (2nd vs lowest UPF category — weight ratio)",

gp = gpar(fontsize = 14, fontface = "bold"), just = "top"), padding = unit(5, "mm"))

FPwt3 <- arrangeGrob(FPwt3, top = textGrob("CVD events (3rd vs lowest UPF category — weight ratio)",

gp = gpar(fontsize = 14, fontface = "bold"), just = "top"), padding = unit(5, "mm"))

FPwt4 <- arrangeGrob(FPwt4, top = textGrob("CVD events (4th vs lowest UPF category — weight ratio)",

gp = gpar(fontsize = 14, fontface = "bold"), just = "top"), padding = unit(15, "mm"))

FPwt5 <- arrangeGrob(FPwt5, top = textGrob("CVD events (5th vs lowest UPF category — weight ratio)",

gp = gpar(fontsize = 14, fontface = "bold"), just = "top"), padding = unit(15, "mm"))

png("ForestPlots_CVD_Weight_Categories.png",

width = 20, height = 15, units = "in", res = 250)

grid.arrange(FPwt2, FPwt3, FPwt4, FPwt5, ncol = 2)

dev.off()

\# =============================================================================

\# SECTION 4: CVD EVENTS BY UPF INTAKE (CALORIE/KCAL RATIO)

\# =============================================================================

kcal <- UPF_CVD_No_HTN_Kcal

\# -----------------------------------------------------------------------------

\# 4.1 Highest vs Lowest (Calorie Ratio)

\# -----------------------------------------------------------------------------

Fplotkcal_hl <- metagen(

TE	= logrr_h_l,

lower	= LB_h_l,

upper	= UB_h_l,

studlab = Author_Name_Year,

data	= UPF_CVD_Kcal,

sm	= "RR",

method.tau = "REML"

)

png("forest_plot_CVD_kcal_highest_vs_lowest.png", width = 800, height = 500)

forest(Fplotkcal_hl, sortvar = TE, random = TRUE, common = TRUE,

prediction = TRUE, digits = 2)

dev.off()

\# -----------------------------------------------------------------------------

\# 4.2 Categories 2–5 vs Lowest (Calorie Ratio)

\# -----------------------------------------------------------------------------

\# Second category (exclude rows 3, 5)

kcal_cat2 <- kcal[-c(3, 5), ]

Fplotkcal_cat2 <- metagen(

TE = log(effect1), lower = log(LB1), upper = log(UB1),

studlab = Author_Name_Year, data = kcal_cat2, sm = "RR", method.tau = "REML"

)

\# Third category (exclude rows 1, 3, 5)

kcal_cat3 <- kcal[-c(1, 3, 5), ]

Fplotkcal_cat3 <- metagen(

TE = log(effect2), lower = log(LB2), upper = log(UB2),

studlab = Author_Name_Year, data = kcal_cat3, sm = "RR", method.tau = "REML"

)

\# Fourth category (exclude rows 1, 4, 5)

kcal_cat4 <- kcal[-c(1, 4, 5), ]

Fplotkcal_cat4 <- metagen(

TE = log(effect3), lower = log(LB3), upper = log(UB3),

studlab = Author_Name_Year, data = kcal_cat4, sm = "RR", method.tau = "REML"

)

\# Fifth category (exclude rows 1, 3, 4)

kcal_cat5 <- kcal[-c(1, 3, 4), ]

Fplotkcal_cat5 <- metagen(

TE = log(effect4), lower = log(LB4), upper = log(UB4),

studlab = Author_Name_Year, data = kcal_cat5, sm = "RR", method.tau = "REML"

)

\# -----------------------------------------------------------------------------

\# 4.3 Combined Forest Plot — All Calorie-Ratio Categories

\# -----------------------------------------------------------------------------

FPkcal2 <- grid.grabExpr(forest (Fplotkcal_cat2, sortvar = TE, backtransf = TRUE, digits = 2))

FPkcal3 <- grid.grabExpr(forest (Fplotkcal_cat3, sortvar = TE, backtransf = TRUE, digits = 2))

FPkcal4 <- grid.grabExpr(forest (Fplotkcal_cat4, sortvar = TE, backtransf = TRUE, digits = 2))

FPkcal5 <- grid.grabExpr(forest (Fplotkcal_cat5, sortvar = TE, backtransf = TRUE, digits = 2))

FPkcal2 <- arrangeGrob(FPkcal2, top = textGrob("CVD events (2nd vs lowest UPF category — kcal ratio)",

gp = gpar(fontsize = 14, fontface = "bold"), just = "top"), padding = unit(5, "mm"))

FPkcal3 <- arrangeGrob(FPkcal3, top = textGrob("CVD events (3rd vs lowest UPF category — kcal ratio)",

gp = gpar(fontsize = 14, fontface = "bold"), just = "top"), padding = unit(5, "mm"))

FPkcal4 <- arrangeGrob(FPkcal4, top = textGrob("CVD events (4th vs lowest UPF category — kcal ratio)",

gp = gpar(fontsize = 14, fontface = "bold"), just = "top"), padding = unit(15, "mm"))

FPkcal5 <- arrangeGrob(FPkcal5, top = textGrob("CVD events (5th vs lowest UPF category — kcal ratio)",

gp = gpar(fontsize = 14, fontface = "bold"), just = "top"), padding = unit(15, "mm"))

png("ForestPlots_CVD_Kcal_Categories.png",

width = 19, height = 11, units = "in", res = 300)

grid.arrange(FPkcal2, FPkcal3, FPkcal4, FPkcal5, ncol = 2)

dev.off()

\# -----------------------------------------------------------------------------

\# 4.4 Combined Forest Plot — Weight vs Calorie Ratio (Highest vs Lowest)

\# -----------------------------------------------------------------------------

FP_wt_hl	<- grid.grabExpr(forest(Fplotwt_hl, sortvar = TE, random = TRUE,

common = TRUE, prediction = TRUE, digits = 2, leftcols = "studlab"))

FP_kcal_hl <- grid.grabExpr(forest(Fplotkcal_hl, sortvar = TE, random = TRUE,

common = TRUE, prediction = TRUE, digits = 2, leftcols = "studlab"))

FP_wt_hl <- arrangeGrob(FP_wt_hl,

top = textGrob("CVD events — highest vs lowest UPF intake (weight ratio)",

gp = gpar(fontsize = 14, fontface = "bold"), just = "top"),

padding = unit(5, "mm"))

FP_kcal_hl <- arrangeGrob(FP_kcal_hl,

top = textGrob("CVD events — highest vs lowest UPF intake (calorie ratio)",

gp = gpar(fontsize = 14, fontface = "bold"), just = "top"),

padding = unit(15, "mm"))

png("ForestPlots_CVD_Weight_vs_Kcal_Merged.png",

width = 10, height = 12, units = "in", res = 100)

grid.arrange(FP_wt_hl, FP_kcal_hl, ncol = 1)

dev.off()

\# =============================================================================

\# SECTION 5: HYPERTENSION — UPF INTAKE

\# =============================================================================

UPF_HTN <- UPF_hypertention_MA

\# -----------------------------------------------------------------------------

\# 5.1 Highest vs Lowest (Hypertension)

\# -----------------------------------------------------------------------------

FplotHTN_hl <- metagen(

TE	= logrr_h_l,

lower	= lb_h_l,

upper	= ub_h_l,

studlab = Author_Name_Year,

data	= UPF_HTN,

sm	= "RR",

method.tau = "REML"

)

png("forest_plot_HTN_highest_vs_lowest.png", width = 800, height = 600)

forest(FplotHTN_hl, sortvar = TE, random = TRUE, common = TRUE,

prediction = TRUE, digits = 2,

text.random = "Hypertension (random effects model)",

text.common = "Hypertension (fixed effects model)")

dev.off()

\# -----------------------------------------------------------------------------

\# 5.2 Categories 2–5 vs Lowest (Hypertension)

\# -----------------------------------------------------------------------------

\# Second category

FplotHTN_cat2 <- metagen(

TE = log(effect1), lower = log(LB1), upper = log(UB1),

studlab = Author_Name_Year, data = UPF_HTN, sm = "RR", method.tau = "REML"

)

\# Third category

FplotHTN_cat3 <- metagen(

TE = log(effect2), lower = log(LB2), upper = log(UB2),

studlab = Author_Name_Year, data = UPF_HTN, sm = "RR", method.tau = "REML"

)

\# Fourth category (exclude rows 2, 6, 8)

UPF_HTN_cat4 <- UPF_HTN[-c(2, 6, 8), ]

FplotHTN_cat4 <- metagen(

TE = log(effect3), lower = log(LB3), upper = log(UB3),

studlab = Author_Name_Year, data = UPF_HTN_cat4, sm = "RR", method.tau = "REML"

)

\# Fifth category (select rows 1, 5, 9)

UPF_HTN_cat5 <- UPF_HTN[c(1, 5, 9), ]

FplotHTN_cat5 <- metagen(

TE = log(effect4), lower = log(LB4), upper = log(UB4),

studlab = Author_Name_Year, data = UPF_HTN_cat5, sm = "RR", method.tau = "REML"

)

\# -----------------------------------------------------------------------------

\# 5.3 Combined Forest Plot — All Hypertension Categories

\# -----------------------------------------------------------------------------

FPHTN2 <- grid.grabExpr(forest(FplotHTN_cat2, sortvar = TE, random = TRUE,

common = TRUE, prediction = TRUE, digits = 2, leftcols = "studlab"))

FPHTN3 <- grid.grabExpr(forest(FplotHTN_cat3, sortvar = TE, random = TRUE,

common = TRUE, prediction = TRUE, digits = 2, leftcols = "studlab"))

FPHTN4 <- grid.grabExpr(forest(FplotHTN_cat4, sortvar = TE, random = TRUE,

common = TRUE, prediction = TRUE, digits = 2, leftcols = "studlab"))

FPHTN5 <- grid.grabExpr(forest(FplotHTN_cat5, sortvar = TE, random = TRUE,

common = TRUE, prediction = TRUE, digits = 2, leftcols = "studlab"))

FPHTN2 <- arrangeGrob(FPHTN2, top = textGrob("Hypertension (2nd vs lowest UPF category)",

gp = gpar(fontsize = 14, fontface = "bold"), just = "top"), padding = unit(5, "mm"))

FPHTN3 <- arrangeGrob(FPHTN3, top = textGrob("Hypertension (3rd vs lowest UPF category)",

gp = gpar(fontsize = 14, fontface = "bold"), just = "top"), padding = unit(5, "mm"))

FPHTN4 <- arrangeGrob(FPHTN4, top = textGrob("Hypertension (4th vs lowest UPF category)",

gp = gpar(fontsize = 14, fontface = "bold"), just = "top"), padding = unit(15, "mm"))

FPHTN5 <- arrangeGrob(FPHTN5, top = textGrob("Hypertension (5th vs lowest UPF category)",

gp = gpar(fontsize = 14, fontface = "bold"), just = "top"), padding = unit(15, "mm"))

png("ForestPlots_HTN_UPF_Categories.png",

width = 19, height = 11, units = "in", res = 300)

grid.arrange(FPHTN2, FPHTN3, FPHTN4, FPHTN5, ncol = 2)

dev.off()

\# =============================================================================

\# SECTION 6: CORONARY HEART DISEASE (CHD) — UPF INTAKE

\# =============================================================================

UPF_CHD <- UPF_CHD_MA[-c(10, 11, 12), ]	# Exclude rows with missing outcome

\# -----------------------------------------------------------------------------

\# 6.1 Highest vs Lowest (CHD)

\# -----------------------------------------------------------------------------

FplotCHD_hl <- metagen(

TE	= logrr_h_l,

lower	= lb_h_l,

upper	= ub_h_l,

studlab = Author_Name_Year,

data	= UPF_CHD,

sm	= "RR",

method.tau = "REML"

)

png("forest_plot_CHD_highest_vs_lowest.png", width = 800, height = 500)

forest(FplotCHD_hl, sortvar = TE, random = TRUE, common = TRUE,

prediction = TRUE, digits = 2)

dev.off()

\# -----------------------------------------------------------------------------

\# 6.2 Categories 2–5 vs Lowest (CHD)

\# -----------------------------------------------------------------------------

\# Second category

FplotCHD_cat2 <- metagen(

TE = log(effect1), lower = log(LB1), upper = log(UB1),

studlab = Author_Name_Year, data = UPF_CHD, sm = "RR", method.tau = "REML"

)

\# Third category (exclude row 9)

CHD_cat3 <- UPF_CHD[-9, ]

FplotCHD_cat3 <- metagen(

TE = log(effect2), lower = log(LB2), upper = log(UB2),

studlab = Author_Name_Year, data = CHD_cat3, sm = "RR", method.tau = "REML"

)

\# Fourth category (same subset as third)

FplotCHD_cat4 <- metagen(

TE = log(effect3), lower = log(LB3), upper = log(UB3),

studlab = Author_Name_Year, data = CHD_cat3, sm = "RR", method.tau = "REML"

)

\# Fifth category (select rows 1, 5, 7)

UPF_CHD_cat5 <- UPF_CHD[c(1, 5, 7), ]

FplotCHD_cat5 <- metagen(

TE = log(effect4), lower = log(LB4), upper = log(UB4),

studlab = Author_Name_Year, data = UPF_CHD_cat5, sm = "RR", method.tau = "REML"

)

\# -----------------------------------------------------------------------------

\# 6.3 Combined Forest Plot — All CHD Categories

\# -----------------------------------------------------------------------------

FPCHD2 <- grid.grabExpr(forest(FplotCHD_cat2, sortvar = TE, random = TRUE,

common = TRUE, prediction = TRUE, digits = 2, leftcols = "studlab"))

FPCHD3 <- grid.grabExpr(forest(FplotCHD_cat3, sortvar = TE, random = TRUE,

common = TRUE, prediction = TRUE, digits = 2, leftcols = "studlab"))

FPCHD4 <- grid.grabExpr(forest(FplotCHD_cat4, sortvar = TE, random = TRUE,

common = TRUE, prediction = TRUE, digits = 2, leftcols = "studlab"))

FPCHD5 <- grid.grabExpr(forest(FplotCHD_cat5, sortvar = TE, random = TRUE,

common = TRUE, prediction = TRUE, digits = 2, leftcols = "studlab"))

FPCHD2 <- arrangeGrob(FPCHD2, top = textGrob("CHD (2nd vs lowest UPF category)",

gp = gpar(fontsize = 14, fontface = "bold"), just = "top"), padding = unit(5, "mm"))

FPCHD3 <- arrangeGrob(FPCHD3, top = textGrob("CHD (3rd vs lowest UPF category)",

gp = gpar(fontsize = 14, fontface = "bold"), just = "top"), padding = unit(5, "mm"))

FPCHD4 <- arrangeGrob(FPCHD4, top = textGrob("CHD (4th vs lowest UPF category)",

gp = gpar(fontsize = 14, fontface = "bold"), just = "top"), padding = unit(15, "mm"))

FPCHD5 <- arrangeGrob(FPCHD5, top = textGrob("CHD (5th vs lowest UPF category)",

gp = gpar(fontsize = 14, fontface = "bold"), just = "top"), padding = unit(15, "mm"))

png("ForestPlots_CHD_UPF_Categories.png",

width = 19, height = 10, units = "in", res = 300)

grid.arrange(FPCHD2, FPCHD3, FPCHD4, FPCHD5, ncol = 2)

dev.off()

\# =============================================================================

\# SECTION 7: CEREBROVASCULAR DISEASE (STROKE) — UPF INTAKE

\# =============================================================================

stroke <- UPF_Stroke_MA

\# -----------------------------------------------------------------------------

\# 7.1 Highest vs Lowest (Stroke)

\# -----------------------------------------------------------------------------

FplotStroke_hl <- metagen(

TE	= logrr_h_l,

lower	= lb_h_l,

upper	= ub_h_l,

studlab = Author_Name_Year,

data	= stroke,

sm	= "RR",

method.tau = "REML"

)

png("forest_plot_Stroke_highest_vs_lowest.png", width = 800, height = 500)

forest(FplotStroke_hl, sortvar = TE, random = TRUE, common = TRUE,

prediction = TRUE, digits = 2)

dev.off()

\# -----------------------------------------------------------------------------

\# 7.2 Categories 2–5 vs Lowest (Stroke)

\# -----------------------------------------------------------------------------

\# Second category

FplotStroke_cat2 <- metagen(

TE = log(effect1), lower = log(LB1), upper = log(UB1),

studlab = Author_Name_Year, data = stroke, sm = "RR", method.tau = "REML"

)

\# Third category

FplotStroke_cat3 <- metagen(

TE = log(effect2), lower = log(LB2), upper = log(UB2),

studlab = Author_Name_Year, data = stroke, sm = "RR", method.tau = "REML"

)

\# Fourth category

FplotStroke_cat4 <- metagen(

TE = log(effect3), lower = log(LB3), upper = log(UB3),

studlab = Author_Name_Year, data = stroke, sm = "RR", method.tau = "REML"

)

\# Fifth category (select rows 1, 8)

stroke_cat5 <- stroke[c(1, 8), ]

FplotStroke_cat5 <- metagen(

TE = log(effect4), lower = log(LB4), upper = log(UB4),

studlab = Author_Name_Year, data = stroke_cat5, sm = "RR", method.tau = "REML"

)

\# -----------------------------------------------------------------------------

\# 7.3 Combined Forest Plot — All Stroke Categories

\# -----------------------------------------------------------------------------

FPstroke2 <- grid.grabExpr(forest(FplotStroke_cat2, sortvar = TE, random = TRUE,

common = TRUE, prediction = TRUE, digits = 2, leftcols = "studlab"))

FPstroke3 <- grid.grabExpr(forest(FplotStroke_cat3, sortvar = TE, random = TRUE,

common = TRUE, prediction = TRUE, digits = 2, leftcols = "studlab"))

FPstroke4 <- grid.grabExpr(forest(FplotStroke_cat4, sortvar = TE, random = TRUE,

common = TRUE, prediction = TRUE, digits = 2, leftcols = "studlab"))

FPstroke5 <- grid.grabExpr(forest(FplotStroke_cat5, sortvar = TE, random = TRUE,

common = TRUE, prediction = TRUE, digits = 2, leftcols = "studlab"))

FPstroke2 <- arrangeGrob(FPstroke2, top = textGrob("Cerebrovascular disease (2nd vs lowest UPF category)",

gp = gpar(fontsize = 14, fontface = "bold"), just = "top"), padding = unit(5, "mm"))

FPstroke3 <- arrangeGrob(FPstroke3, top = textGrob("Cerebrovascular disease (3rd vs lowest UPF category)",

gp = gpar(fontsize = 14, fontface = "bold"), just = "top"), padding = unit(5, "mm"))

FPstroke4 <- arrangeGrob(FPstroke4, top = textGrob("Cerebrovascular disease (4th vs lowest UPF category)",

gp = gpar(fontsize = 14, fontface = "bold"), just = "top"), padding = unit(15, "mm"))

FPstroke5 <- arrangeGrob(FPstroke5, top = textGrob("Cerebrovascular disease (5th vs lowest UPF category)",

gp = gpar(fontsize = 14, fontface = "bold"), just = "top"), padding = unit(15, "mm"))

png("ForestPlots_Stroke_UPF_Categories.png",

width = 19, height = 10, units = "in", res = 300)

grid.arrange(FPstroke2, FPstroke3, FPstroke4, FPstroke5, ncol = 2)

dev.off()

\# -----------------------------------------------------------------------------

\# 7.4 Combined Forest Plot — CHD, Stroke, Hypertension (Highest vs Lowest)

\# -----------------------------------------------------------------------------

FP_CHD_hl	<- grid.grabExpr(forest(FplotCHD_hl, sortvar = TE, random = TRUE,

common = TRUE, prediction = TRUE, digits = 2, leftcols = "studlab"))

FP_Stroke_hl <- grid.grabExpr(forest(FplotStroke_hl, sortvar = TE, random = TRUE,

common = TRUE, prediction = TRUE, digits = 2, leftcols = "studlab"))

FP_HTN_hl	<- grid.grabExpr(forest(FplotHTN_hl, sortvar = TE, random = TRUE,

common = TRUE, prediction = TRUE, digits = 2, leftcols = "studlab"))

FP_CHD_hl	<- arrangeGrob(FP_CHD_hl,

top = textGrob("Coronary heart disease (highest vs lowest UPF intake)",

gp = gpar(fontsize = 14, fontface = "bold")))

FP_Stroke_hl <- arrangeGrob(FP_Stroke_hl,

top = textGrob("Cerebrovascular disease (highest vs lowest UPF intake)",

gp = gpar(fontsize = 14, fontface = "bold")))

FP_HTN_hl	<- arrangeGrob(FP_HTN_hl,

top = textGrob("Hypertension (highest vs lowest UPF intake)",

gp = gpar(fontsize = 14, fontface = "bold")))

png("ForestPlots_CHD_Stroke_HTN_Combined.png",

width = 11, height = 15, units = "in", res = 250)

grid.arrange(FP_CHD_hl, FP_Stroke_hl, FP_HTN_hl, ncol = 1)

dev.off()

\# =============================================================================

\# SECTION 8: SENSITIVITY ANALYSES

\# =============================================================================

\# Re-load primary dataset for sensitivity analyses

upf_cvd <- UPF_CVD_Regardless_UA_Nodup_No_HTN

Fplot <- metagen(

TE = h_L_logrr, lower = LB_h_l, upper = UB_h_l,

studlab = Author_Name_Year, data = upf_cvd, sm = "RR", method.tau = "REML"

)

\# -----------------------------------------------------------------------------

\# 8.1 Leave-One-Out (LOO) Sensitivity Analysis

\# -----------------------------------------------------------------------------

full_effect <- Fplot$TE.random

threshold	<- 0.1	# Threshold for flagging influential studies

loo_results <- data.frame(

Study_Left_Out = upf_cvd$Author_Name_Year,

TE_random	= NA,

lower_random	= NA,

upper_random	= NA,

stringsAsFactors = FALSE

)

for (i in 1:nrow(upf_cvd)) {

temp <- metagen(

TE = h_L_logrr, lower = LB_h_l, upper = UB_h_l,

studlab = Author_Name_Year, data = upf_cvd[-i, ], sm = "RR", method.tau = "REML"

)

loo_results$TE_random[i]	<- temp$TE.random

loo_results$lower_random[i] <- temp$lower.random

loo_results$upper_random[i] <- temp$upper.random

}

loo_results$Influential <- abs(loo_results$TE_random - full_effect) > threshold

\# Exponentiate and format for export

loo_exp <- loo_results %>%

mutate(

RR	= round(exp(TE_random), 2),

Lower_CI = round(exp(lower_random), 2),

Upper_CI = round(exp(upper_random), 2),

CI	= paste0(Lower_CI, " – ", Upper_CI)

) %>%

select(Study_Left_Out, RR, CI, Influential)

\# Export to Word

doc <- read_docx() %>%

body_add_par("Leave-One-Out Sensitivity Analysis Results", style = "heading 1") %>%

body_add_flextable(autofit(flextable(loo_exp)))

print(doc, target = "LOO_Sensitivity_Results.docx")

\# LOO Forest plot (influential studies highlighted in red)

loo_meta <- metagen(

TE = loo_results$TE_random, lower = loo_results$lower_random,

upper = loo_results$upper_random, studlab = loo_results$Study_Left_Out, sm = "RR"

)

n_studies <- length(loo_meta$TE)

col_flag <- ifelse(loo_results$Influential[1:n_studies], "red", "black")

png("ForestPlot_LOO_Sensitivity.png", width = 760, height = 560)

forest(loo_meta,

comb.random = FALSE,

xlab	= "Pooled Effect (random effects)",

leftcols	= "studlab",

sortvar	= loo_meta$TE,

col.square	= col_flag,

col.study	= col_flag,

refline	= full_effect)

dev.off()

\# -----------------------------------------------------------------------------

\# 8.2 Covariate Adjustment Sensitivity Analysis

\#	(Sub-optimal/optimal adjustment × overfitted/not-overfitted models)

\# -----------------------------------------------------------------------------

\# Sub-optimal and overfitted

Fplot_so_of <- metagen(

TE = h_L_logrr, lower = LB_h_l, upper = UB_h_l,

studlab = Author_Name_Year, data = sub_Optimal_Overfitted,

sm = "RR", method.tau = "REML"

)

\# Optimal and overfitted

Fplot_o_of <- metagen(

TE = h_L_logrr, lower = LB_h_l, upper = UB_h_l,

studlab = Author_Name_Year, data = Optimal_Overfitted,

sm = "RR", method.tau = "REML"

)

\# Sub-optimal and not overfitted

Fplot_so_nof <- metagen(

TE = h_L_logrr, lower = LB_h_l, upper = UB_h_l,

studlab = Author_Name_Year, data = Sub_Optimal_not_overfitted,

sm = "RR", method.tau = "REML"

)

FP_so_of <- grid.grabExpr(forest(Fplot_so_of, sortvar = TE, random = TRUE,

common = TRUE, prediction = TRUE, digits = 2))

FP_o_of	<- grid.grabExpr(forest(Fplot_o_of, sortvar = TE, random = TRUE,

common = TRUE, prediction = TRUE, digits = 2))

FP_so_nof <- grid.grabExpr(forest(Fplot_so_nof, sortvar = TE, random = TRUE,

common = TRUE, prediction = TRUE, digits = 2))

FP_so_of <- arrangeGrob(FP_so_of, top = textGrob(

"CVD — sub-optimal covariate adjustment & overfitted models",

gp = gpar(fontsize = 14, fontface = "bold"), just = "top"), padding = unit(5, "mm"))

FP_o_of	<- arrangeGrob(FP_o_of, top = textGrob(

"CVD — optimal covariate adjustment & overfitted models",

gp = gpar(fontsize = 14, fontface = "bold"), just = "top"), padding = unit(5, "mm"))

FP_so_nof <- arrangeGrob(FP_so_nof, top = textGrob(

"CVD — sub-optimal covariate adjustment & non-overfitted models",

gp = gpar(fontsize = 14, fontface = "bold"), just = "top"), padding = unit(15, "mm"))

png("ForestPlots_Covariate_Adjustment_Sensitivity.png",

width = 20, height = 14, units = "in", res = 300)

grid.arrange(FP_so_of, FP_o_of, FP_so_nof, ncol = 2)

dev.off()

\# =============================================================================

\# SECTION 9: PUBLICATION BIAS

\# =============================================================================

\# Helper function to run and export all publication bias tests

run_pub_bias <- function(meta_obj, label, k_min = 10) {

\# Funnel plot

png(paste0("FunnelPlot_", label, ".png"), width = 1000, height = 800)

funnel(meta_obj, xlab = "Risk Ratio", back = TRUE, studlab = TRUE)

dev.off()

\# Egger's test

cat("\n--- Egger's Test:", label, "---\n")

print(metabias(meta_obj, method.bias = "linreg", k.min = k_min))

\# Begg's rank correlation test

cat("\n--- Begg's Test:", label, "---\n")

print(metabias(meta_obj, method.bias = "rank", k.min = k_min))

\# Trim-and-fill

taf <- trimfill(meta_obj)

cat("\n--- Trim-and-Fill:", label, "---\n")

print(summary(taf))

taf_table <- data.frame(

Model	= c("Unadjusted", "Trim-and-Fill adjusted"),

RR	= round(exp(c(meta_obj$TE.random, taf$TE.random)), 2),

Lower_CI = round(exp(c(meta_obj$lower.random, taf$lower.random)), 2),

Upper_CI = round(exp(c(meta_obj$upper.random, taf$upper.random)), 2),

k	= c(meta_obj$k, taf$k)

) %>%

mutate(CI = paste0(Lower_CI, " – ", Upper_CI))

doc <- read_docx() %>%

body_add_par(paste("Trim-and-Fill Results:", label), style = "heading 1") %>%

body_add_flextable(autofit(flextable(taf_table)))

print(doc, target = paste0("TrimFill_", label, ".docx"))

png(paste0("FunnelPlot_TrimFill_", label, ".png"), width = 1200, height = 1000)

funnel(taf, xlab = paste("Risk Ratio — Trim and Fill (", label, ")"))

dev.off()

}

\# Run for all primary outcomes

run_pub_bias(Fplot, label = "CVD_Overall", k_min = 10)

run_pub_bias(FplotHTN_hl, label = "Hypertension", k_min = 10)

run_pub_bias(FplotCHD_hl, label = "CHD", k_min = 9)

run_pub_bias(FplotStroke_hl,label = "Stroke", k_min = 8)

run_pub_bias(Fplotwt_hl, label = "CVD_Weight", k_min = 10)

run_pub_bias(Fplotkcal_hl, label = "CVD_Kcal", k_min = 10)

\# =============================================================================

\# SECTION 10: META-REGRESSION

\# =============================================================================

upf_cvd <- UPF_CVD_Regardless_UA_Nodup_No_HTN

\# Fit random-effects model using standard error

REMA <- metagen(

TE	= h_L_logrr,

seTE	= Se_h_l,

studlab = Author_Name_Year,

data	= upf_cvd,

common = FALSE,

random = TRUE,

method.tau = "REML"

)

\# Multiple meta-regression: region, sex, unit of analysis,

\# dietary tool, outcome type, and follow-up duration

m_reg <- metareg(

REMA,

~ Region + Sex + UOA + Diet_tool + Outcome + FY_binary_10

)

summary(m_reg)

\# -----------------------------------------------------------------------------

\# 10.1 Bubble Plots (one per moderator)

\# -----------------------------------------------------------------------------

png("BubblePlots_MetaRegression_CVD.png", width = 1600, height = 1000, res = 100)

par(mfrow = c(2, 3))

bubble(m_reg, mod = "Region", xlab = "Region")

bubble(m_reg, mod = "Sex", xlab = "Sex")

bubble(m_reg, mod = "UOA", xlab = "Unit of analysis")

bubble(m_reg, mod = "Diet_tool", xlab = "Dietary assessment tool")

bubble(m_reg, mod = "Outcome", xlab = "CVD outcome type")

bubble(m_reg, mod = "FY_binary_10", xlab = "Follow-up ≥10 years")

dev.off()

\# -----------------------------------------------------------------------------

\# 10.2 Export Meta-Regression Results (Risk Ratios with 95% CI) to Word

\# -----------------------------------------------------------------------------

rr_table <- broom::tidy(m_reg, conf.int = TRUE) %>%

mutate(

RR	= round(exp(estimate), 2),

Lower_CI	= round(exp(conf.low), 2),

Upper_CI	= round(exp(conf.high), 2),

CI	= paste0(Lower_CI, " – ", Upper_CI),

p.value	= round(p.value, 3)

) %>%

select(term, RR, CI, p.value)

doc <- read_docx() %>%

body_add_par("Meta-regression results (Risk Ratios, 95% CI)", style = "heading 1") %>%

body_add_flextable(autofit(flextable(rr_table)))

print(doc, target = "MetaRegression_Results_RR.docx")

\# =============================================================================

\# END OF SCRIPT

\# =============================================================================

\# Session information for reproducibility

sessionInfo()
